# Values and preferences in COVID-19 public health guidelines: A systematic review

**DOI:** 10.1101/2024.03.25.24304859

**Authors:** Sarah Kirsh, Michael Ling, Tanvir Jassal, Tyler Pitre, Thomas Pigott, Dena Zeraatkar

## Abstract

**Background:** Internationally accepted standards for trustworthy guidelines include the necessity to ground recommendations in values and preferences. Considering values and preferences respects the rights of citizens to participate in health decision-making and ensures that guidelines align with the needs and priorities of the communities they are intended to serve. Early anecdotal reports suggest that COVID-19 public health guidelines did not consider values and preferences.

**Objective:** To capture and characterize whether and how COVID-19 public health guidelines considered values and preferences.

**Methods:** We performed a systematic review of COVID-19 public health guidelines. We searched the eCOVID19 RecMap platform—a comprehensive international catalog of COVID-19 guidelines—up to July 2023. We included guidelines that made recommendations addressing vaccination, masking, isolation, lockdowns, travel restrictions, contact tracing, infection surveillance, and school closures. Reviewers worked independently and in duplicate to review guidelines for consideration of values and preferences.

**Results:** Our search yielded 129 eligible guidelines, of which 43 (33.3%) were published by national organizations, 73 (56.6%) by international organizations, and 14 (10.9%) by professional societies and associations. Twenty-six (20.2%) guidelines considered values and preferences. Among guidelines that considered values and preferences, most did so to assess the acceptability of recommendations (23; 88.5%) and by referencing published research (24; 92.3%). Guidelines only occasionally engaged laypersons as part of the guideline development group (6; 23.1%). None of the guidelines performed systematic reviews of the literature addressing values and preferences.

**Conclusion:** Most COVID-19 public health guidelines did not consider values and preferences. When values and preferences were considered, it was suboptimal. Disregard for values and preferences in guidelines might have partly contributed to divisive and unpopular COVID-19 policies. Given the possibility of future health emergencies, we recommend guideline developers identify efficient methods for considering values and preferences in crisis situations.

## Background

Public health guidelines provide recommendations intended to optimize population health (1). They are typically published by professional associations and authoritative organizations, examples of which include the World Health Organization (WHO) and the US Centers for Disease Control and Prevention (CDC). The impact of public health guidelines is wide-reaching and diverse—they inform national policies, rules and regulations at schools and workplaces, and messaging to which the public is exposed. Given their significant impact, it is essential for these guidelines to be credible and effective.

For guidelines to be considered credible, they must adhere to certain standards (2–12). Considering public values and preferences—defined as the public’s perspectives, beliefs, expectations, and goals for health and life—is internationally recognized as a critical component of credible guideline development (2, 3, 10). Considering values and preferences respects the rights of citizens to participate in health decision-making, aligns guidelines with the needs and priorities of the communities they are intended to serve, ensures recommendations are logistically feasible and acceptable, and improves support for the recommendations (13, 14). Values and preferences may vary by country or setting, and therefore may be an important lens for contextualizing or adapting guidance between settings (15).

While some public health professionals and decision-makers may argue that scientific evidence alone should inform recommendations, trustworthy guideline development necessitates the synthesis of both scientific evidence and values and preferences to arrive at recommendations (2, 3, 10). Guideline developers face decisions that invariably involve trade-offs between benefits and harms. For example, a new lung cancer screening program might improve survival rates through early detection but could also result in overdiagnosis, false positives, radiation, and divert resources from other healthcare services, potentially increasing morbidity and mortality from other causes. Ideally, the public’s values and preferences should guide the assessment of whether the benefits of a given action outweigh its harms.

For these reasons, nearly all organizations that establish standards for guideline development advise that guidelines also consider values and preferences (2, 3, 10, 16). For example, the AGREE II tool, the gold standard tool for evaluating the quality of guidelines, includes a domain addressing the consideration of values and preferences (3). Likewise, the GRADE Evidence-to-Decision frameworks, the most commonly used frameworks for formulating recommendations, include values and preferences as a key consideration (10).

Guideline developers may incorporate public values and preferences in several ways, such as involving laypersons as part of the guideline development group, reviewing published research on public values and preferences, conducting de novo studies, and providing the public the opportunity to comment on draft recommendations (2, 3, 10, 14, 17–22). Ideally, guideline developers should perform systematic reviews of the literature addressing values and preferences (19, 23, 24). In situations in which guideline developers are unable to find sufficient high-quality evidence, they can perform de novo surveys and focus groups to generate this information. While insights and beliefs of the guideline development group, especially if it includes laypersons, are valuable, they may not fully capture the broader public’s values and preferences, leaving some uncertainty.

The COVID-19 pandemic caused significant threats to health globally. In response, governments implemented policies, informed by public health guidelines, that ranged from stringent measures to curb transmission, including “lockdowns”, to attempts that focused on minimizing disruptions to daily life (25–28). Public health professionals, and ultimately politicians, were confronted with the delicate, ethically complex task of balancing COVID-19 with other public health problems and social and economic well-being. Ideally, public values and preferences should have informed this trade-off.

Early anecdotal reports, however, suggest that public engagement was limited and suboptimal in COVID- 19 public health guidelines (29). In France, for example, the public was not consulted about lockdowns (30). In Germany, early decisions were made only by a handpicked group of scientific experts (31, 32). While rigorous guidelines that considered values and preferences may have been possible, the unprecedented circumstances posed by the COVID-19 pandemic may have adversely impacted guideline development methods and thus guideline quality. Nevertheless, consideration of values and preferences remains important even in crisis situations (33, 34). Even guidance for producing guidelines during emergency situations requires consideration of values and preferences (33). Suboptimal consideration of values and preferences might have contributed to divisive and unpopular policies, the erosion of trust in public health, and limited compliance (35–40).

As the pandemic evolves from an acute threat to a long-term public health challenge, now is a critical time to evaluate and learn from our response. While the COVID-19 pandemic was unprecedented, we anticipate other health emergencies that will also benefit from credible guidelines that are aligned with societal values. We conducted a systematic review of national and international public health COVID-19 guidelines to capture and characterize whether and how they considered public values and preferences.

## Methods

We report our systematic review of COVID-19 public health guidelines following the Preferred Reporting Items for Systematic Reviews and Meta-Analyses (PRISMA) reporting guidelines for systematic reviews (41). Figure 1 presents an overview of our methods.

**Figure 1:**
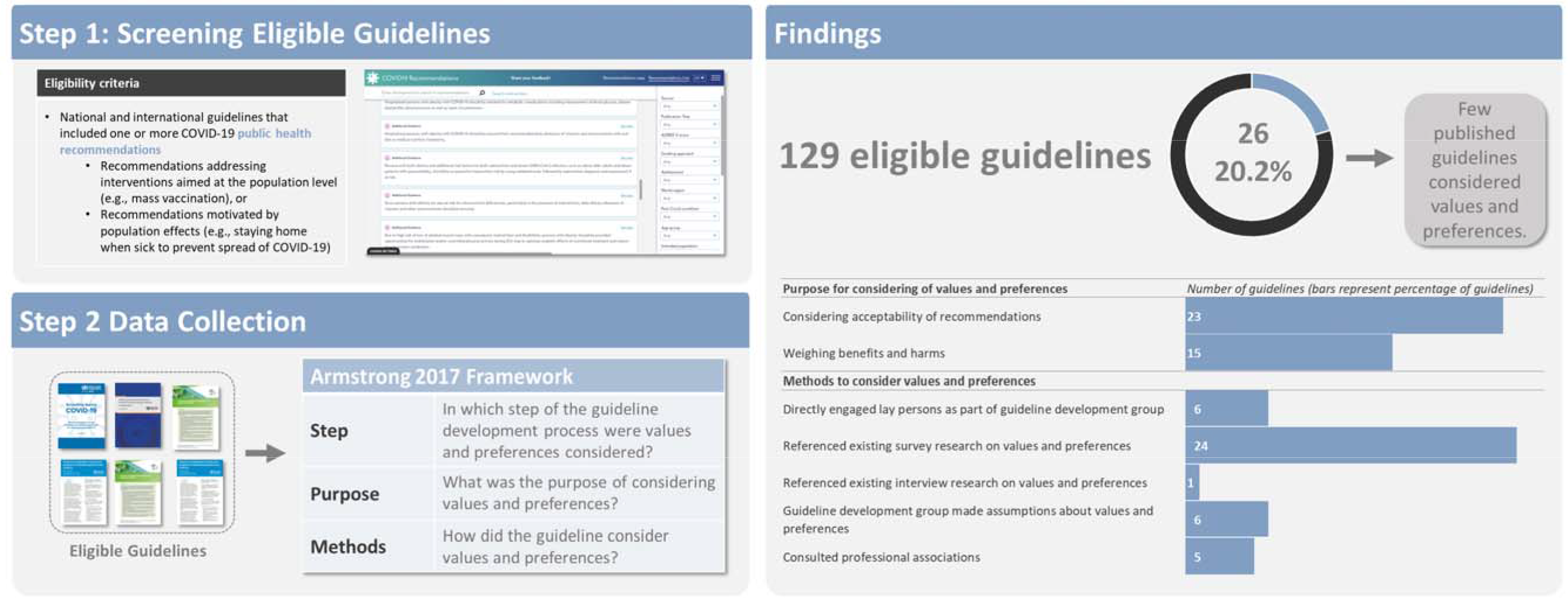
Overview of methods and findings We searched the eCOVID19 RecMap platform—a comprehensive international catalog of COVID-19 guidelines—for COVID-19 public health guidelines containing one or more recommendations addressing vaccination, masking, isolation, lockdowns, travel restrictions, contact tracing, infection surveillance, and school closures. Reviewers worked independently and in duplicate to assess guidelines for eligibility. From eligible guidelines, reviewers again worked independently and duplicate to collect information on whether and how guidelines considered public values and preferences. Reviewers used a previously established framework by Armstrong and colleagues classify the steps in guideline development in which values and preferences were considered, the purpose for which values and preferences were considered, and the methods used to consider values and preferences.

### Search strategy and screening

We intended to include a representative sample of national and international COVID-19 public health guidelines. We anticipated that a search of research databases would identify few eligible guidelines, since most guidelines are typically published on webpages or reports from national and professional organizations instead of journals. Thus, to identify eligible guidelines, we searched the eCOVID19-19 RecMap—a comprehensive open-access digital collection of COVID-19 guidelines and recommendations, developed in collaboration between Cochrane Canada, the WHO Collaborating Center for Infectious Diseases, and the Guidelines International Network, up to July 2023 (42, 43).

The eCOVID19-19 RecMap presents thematically organized lists of recommendations contained in COVID-19 guidelines. Following training and calibration to ensure sufficient agreement, two reviewers worked independently and in duplicate to screen all recommendations catalogued in the eCOVID19 RecMap for eligibility. When reviewers identified an eligible recommendation, they included the guideline containing that recommendation in the present systematic review. Reviewers resolved discrepancies by discussion, or when necessary, by adjudication with a third reviewer.

We supplemented our search by reviewing webpages of national and international guideline producing organizations (i.e., WHO, US CDC, and European Centers for Disease Control) and reviewing the references of the included guidelines.

### Eligibility criteria

We included national and international guidelines that contained one or more COVID-19 public health recommendations. We defined guidelines as documents that described themselves as ‘guidelines’ or as providing recommendations. Distinguishing between clinical practice and public health guidelines is challenging and guidelines may include a combination of clinical practice and public health recommendations. To address this issue, we pragmatically defined public health recommendations as recommendations addressing interventions aimed at the population level instead of at the individual patient level or recommendations that are motivated by population effects (e.g., staying home when sick to prevent spread of COVID-19, lockdowns, closure of educational institutions).

We recognized that not all recommendations will require consideration of values and preferences. For example, the benefits of a particular course of action may obviously and overwhelmingly outweigh potential harms, or vice versa, such that the consideration of values and preferences is unimportant (44, 45). For example, the potential benefits of hand washing overwhelmingly outweigh any potential harms such that a recommendation to practice frequent handwashing does not require consideration of values and preferences. We restricted eligibility to guidelines that made recommendations addressing topics that we considered to be sensitive to values and preferences: vaccination, masking, isolation, lockdowns, travel restrictions, contact tracing, infection surveillance, and school closures. Our selection of these topics was informed by scientific discourse and discussions in social and traditional media. Box 1 describes how recommendations addressing these topics are sensitive to values and preferences.

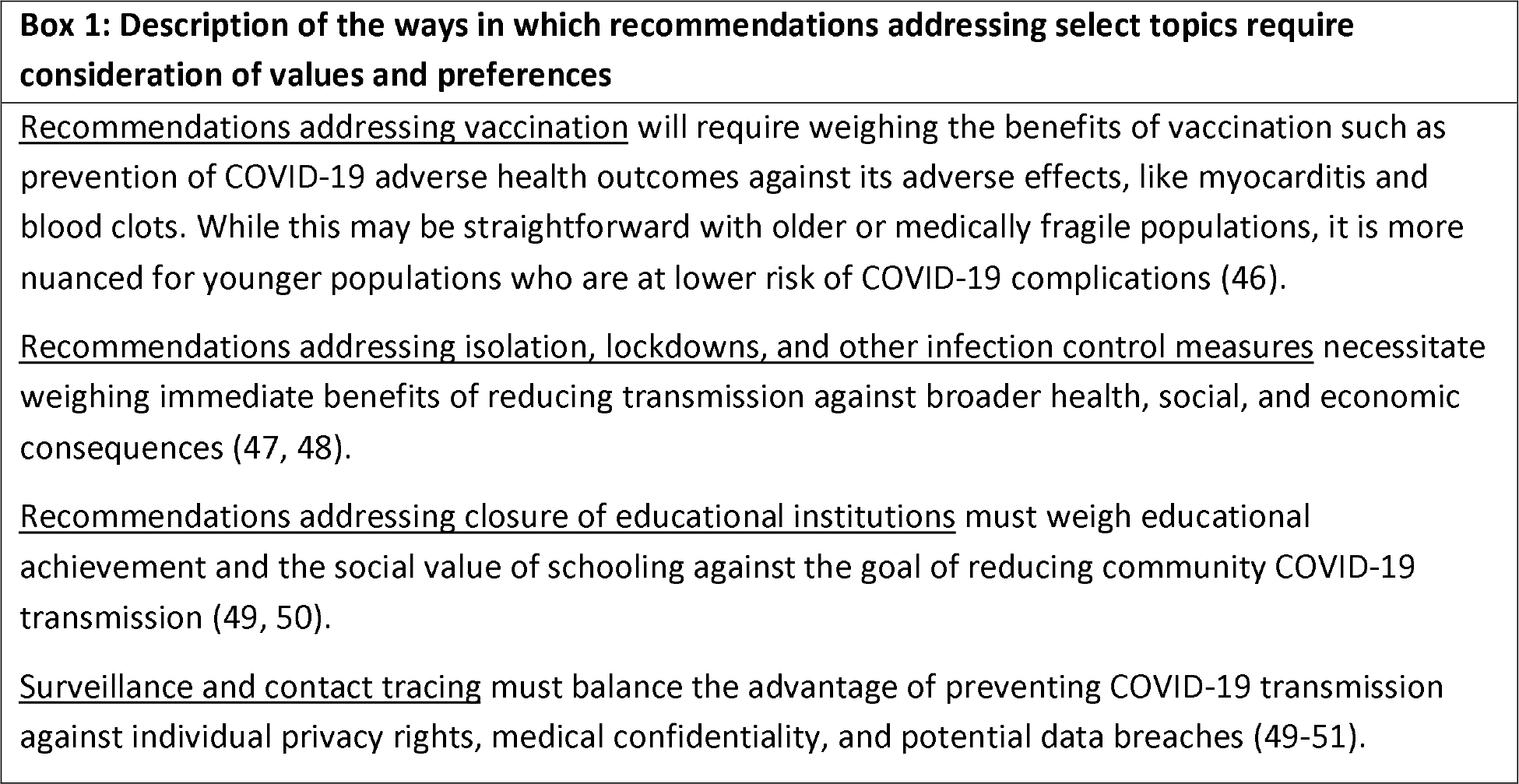

For feasibility, we excluded regional guidelines. We also excluded guidelines addressing the care of patients with COVID-19, the operation of healthcare facilities, performance of diagnostic tests for COVID-19, care of patients with comorbidities during the pandemic, management of long covid, and niche professional activities (e.g., care of heritage collections during COVID-19).

When guidelines were updated, resulting in multiple versions, we included all versions in our study. This approach allowed us to study how consideration of values and preferences may have evolved during the course of the pandemic.

### Data extraction

Following training and calibration to ensure sufficient agreement, two reviewers worked independently and in duplicate and used a pilot-tested structured form to collect information on guideline characteristics, methods, and values and preferences.

We intended to capture whether and how guidelines considered public values and preferences. We used a broad operationalization of values and preferences, defined as public’s perspectives, beliefs, expectations, and goals for health and life, including but not limited to soliciting insights from laypersons as part of the guideline development process or as part of the dissemination process, assessing acceptability and tolerability of potential courses of action, and weighing benefits and harms (2, 3, 10).

The Armstrong framework for patient engagement in guideline development informed the type of data that we collected (18). This framework contains 10 categories describing the steps of the guideline development process in which values and preferences may be considered, 37 categories describing the purpose of considering values and preferences, and 36 categories describing methods by which values and preferences may be considered (18). Box 2 presents an overview of the framework. While this framework was originally conceived for patient engagement in clinical practice guidelines, our examination suggests it is also equally relevant for public health guidelines. We created new categories when we encountered scenarios not included in the framework.

| **Box 2: Overview of the Armstrong framework for patient engagement in guidelines with example categories (18)** |  |
| --- | --- |
| **Values & preferences** | **Example categories** |
| **Step in guideline development process in which values and preferences may be considered** | - Nominating guideline topics<br>- Developing recommendations<br>- Disseminating and implementing recommendations |
| **Purpose of considering values and preferences** | - Identify topics that are important to patients, caregivers, and the community<br>- Assist in translating evidence-based conclusions into meaningful, clear, and respectful recommendations<br>- Endorse guidelines from patient perspective |
| **Methods for considering values and preferences** | - Solicit topic nominations from public<br>- Review existing research on patients' |
|  | preferences<br>- Consult patients, caregivers, and advocacy groups regarding barriers to dissemination and implementation |

For guidelines published in a language other than English, we intended to use Google Translate to translate the guideline document to facilitate extraction and subsequently ask a native language speaker to confirm our extractions. We did not, however, identify any eligible guidelines published in a language other than English. Reviewers resolved discrepancies by discussion, and, if necessary, by adjudication with a third reviewer.

### Data synthesis and analysis

Descriptive statistics summarize guideline characteristics, methods, and consideration of values & preferences. We used content analysis to map the steps in which values and preferences were considered in guideline development, the purpose for considering values and preferences, and methods for considering values and preferences to the categories in the Armstrong framework (18).

We anticipated that consideration of values and preferences across guidelines may differ according to the time at which they were published, the topics they address, the jurisdiction of the guideline (high income versus low/middle income countries), the publishing organization, and whether the guideline used GRADE methods (52). Hence, we also interpret results considering these characteristics.

## Results

### Search results

We reviewed 617 guidelines for eligibility, of which 129 proved eligible. Figure 2 presents the selection of guidelines. Supplement 1 lists included guidelines and Supplement 2 lists the excluded guidelines and reasons for their exclusion.

**Figure 2:**
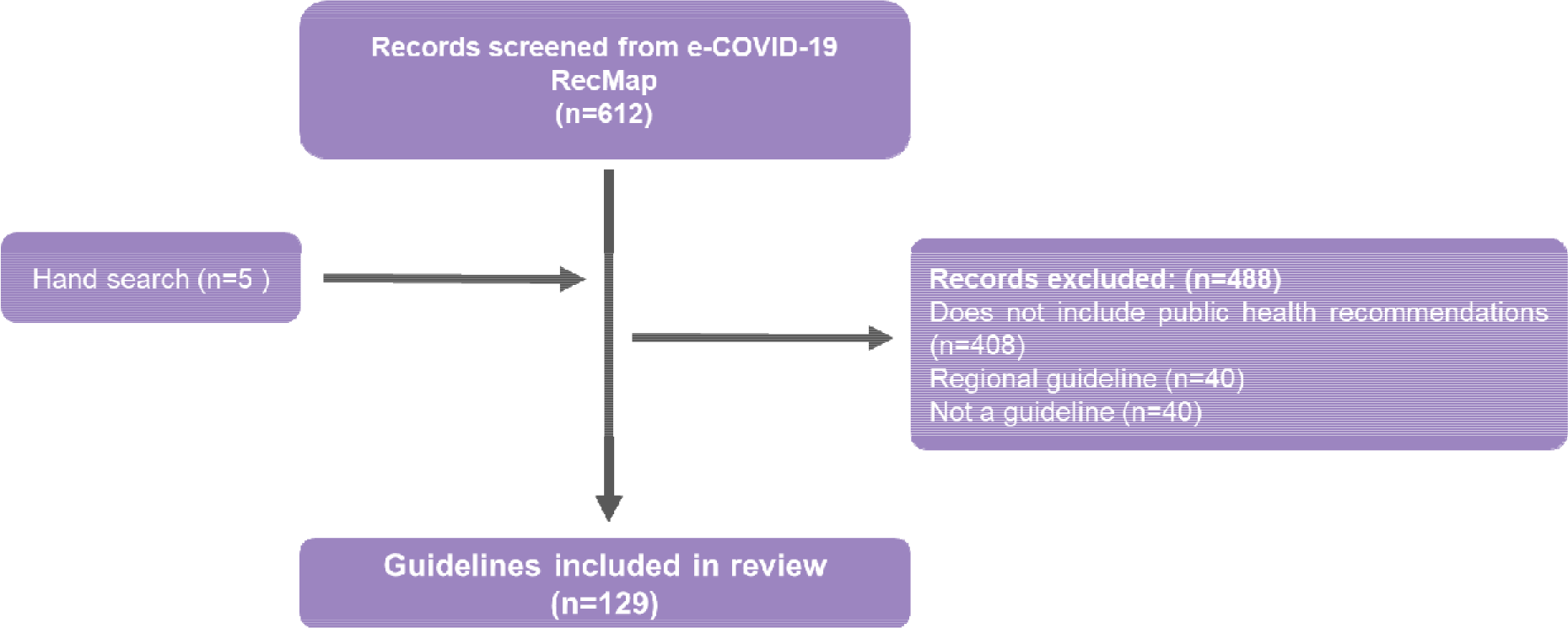
Selection of guidelines We searched the eCOVID19 RecMap platform—a comprehensive international catalog of COVID-19 guidelines—for COVID-19 public health guidelines containing one or more recommendations addressing vaccination, masking, isolation, lockdowns, travel restrictions, contact tracing, infection surveillance, and school closures. We supplemented our search by reviewing webpages of national and international guideline producing organizations (i.e., WHO, US CDC, and European Centers for Disease Control) and reviewing the references of the included guidelines, by which we identified five additional guidelines. Reviewers worked independently and in duplicate to assess guidelines for eligibility. Ultimately, 129 guidelines met eligibility and were included in this study.

### Guideline characteristics

Of 129 included guidelines, 73 (56.6%) were published by international organizations, 43 (33.3%) by national organizations, and 14 (10.9%) by professional societies and associations. We included guidelines from a total of 25 unique government and non-government organizations. Forty (31%) guidelines were published in 2020, 48 (37.2%) in 2021, 33 (25.6%) in 2022 and 8 (6.2%) in 2023. Nearly all guidelines targeted high-income countries or regions (96.9%), typically in North America or Europe. Guidelines seldom disclosed funding (24; 18.6%) or conflicts of interest (35; 27.1%). Five (3.9%) guidelines were updates of previously published guidelines.

Less than one in 10 guidelines used GRADE to evaluate the certainty of evidence (52). An almost equal proportion used GRADE evidence-to-decision frameworks to formulate recommendations (10).

Guidelines addressed a range of topics, with the most common topics being vaccination and vaccine prioritization (36; 27.9%), general public health measures (34; 26.4%), and infection control & prevention (18; 14%). Most guidelines included recommendations that targeted community-dwelling people (42; 32.6%) or were intended to be used by public health practitioners and other decision-makers (59; 45.7%).

### Values and preferences

Of 129 eligible guidelines, 26 (20.2%) considered values and preferences. Seventeen of these guidelines (65.4%) were published by the WHO, 7 (26.9%) by the CDC, and 1 (3.8%) by the American College of Obstetricians and Gynecologists. Six (23.1%) of the guidelines were iterations of the WHO living guideline on infection prevention and control (53–58). None of the guidelines were published in 2020, 8 (30.8%) were published in 2021, 13 (50%) in 2022, and four (15.4%) in 2023. Nearly all guidelines (22; 84.6%) that considered values and preferences also used GRADE methods to evaluate the certainty of evidence and to move from evidence to recommendations. Guidelines addressed either vaccination and vaccine prioritization (16; 61.5%), masking in the community (6; 23.1%), or school & education (2; 7.7%).

Table 2 describes the consideration of values and preferences in guidelines and table 3 describes the guidelines that considered values and preferences. Among guidelines that considered values and preferences, all did so to develop recommendations. Half of these guidelines also recommended for consideration of values and preferences in the implementation of recommendations (11; 42.3%). None of the guidelines considered values in preferences in other steps of the guideline development process, such as nominating or prioritizing topic, drafting recommendations, or disseminating or endorsing the guideline.

**Table 1:**
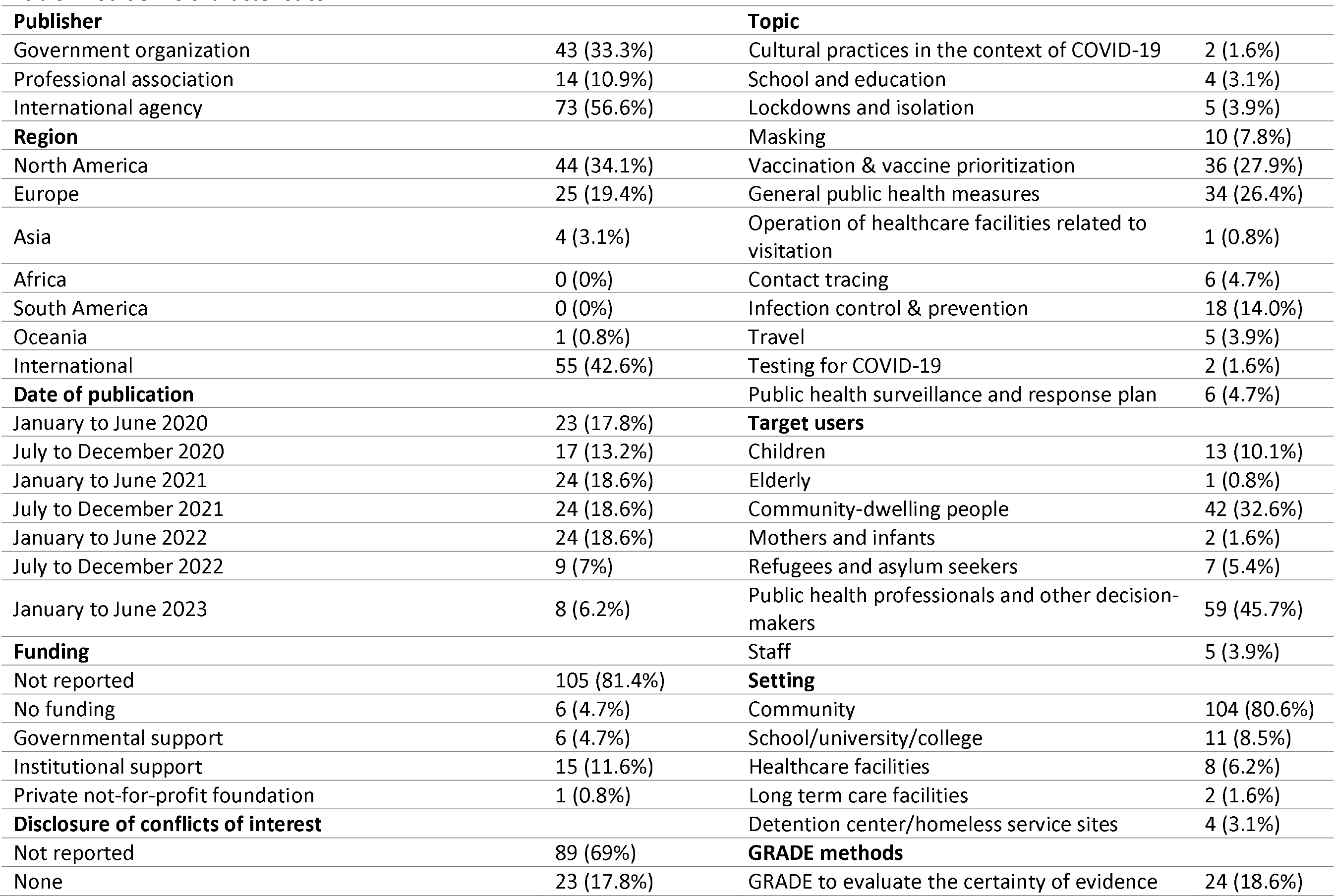
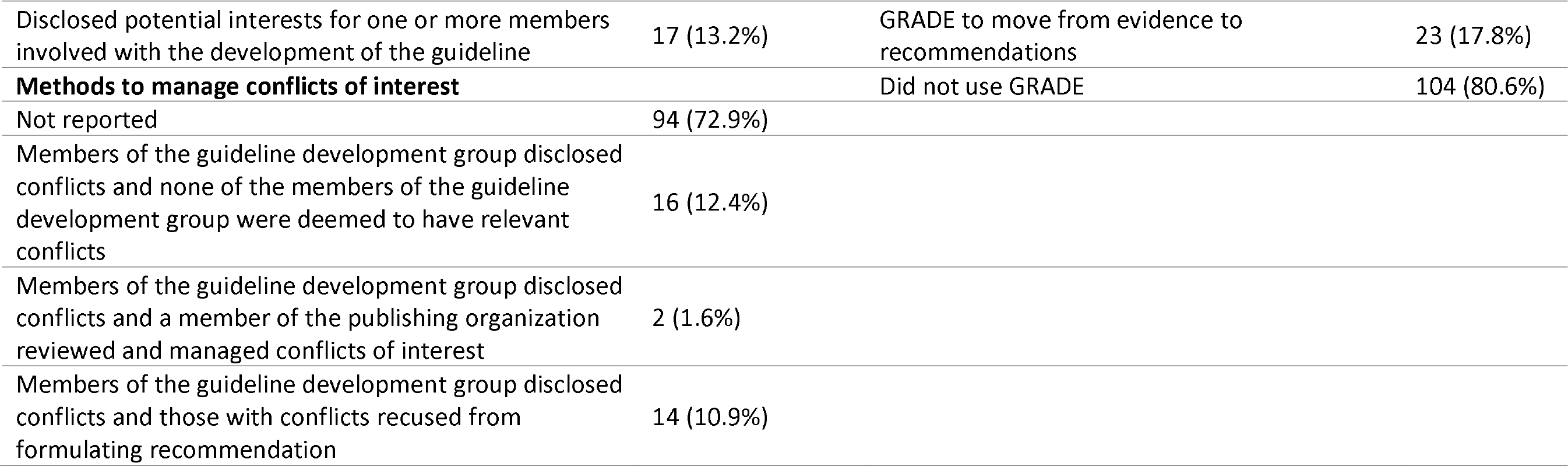
Guideline characteristics.

**Table 2:**
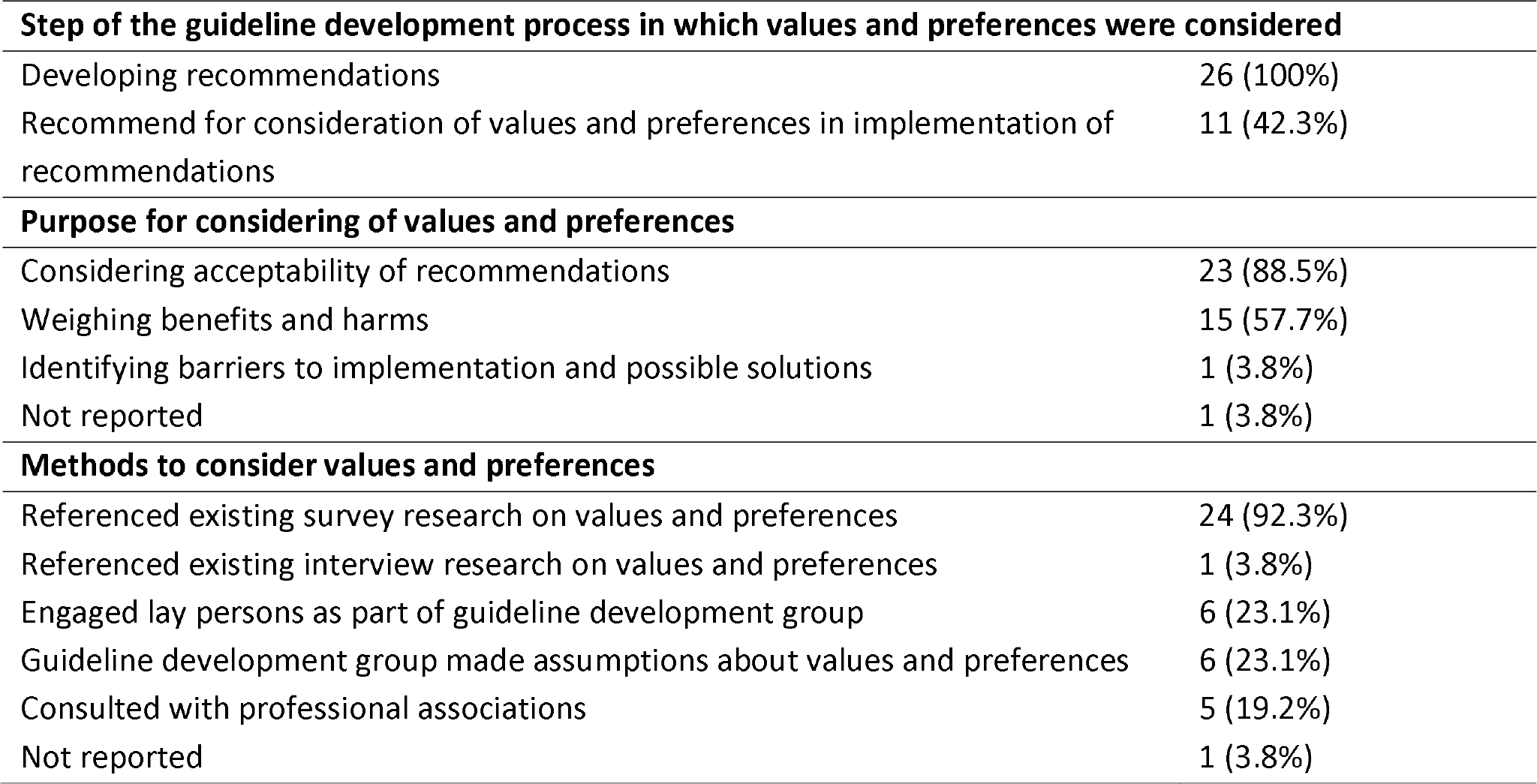
Description of the steps, purpose, and methods by which guidelines considered values and preferences.

| **Step of the guideline development process in which values and preferences were considered** |  |
| --- | --- |
| Developing recommendations | 26 (100%) |
| Recommend for consideration of values and preferences in implementation of recommendations | 11 (42.3%) |
| **Purpose for considering of values and preferences** |  |
| Considering acceptability of recommendations | 23 (88.5%) |
| Weighing benefits and harms | 15 (57.7%) |
| Identifying barriers to implementation and possible solutions | 1 (3.8%) |
| Not reported | 1 (3.8%) |
| **Methods to consider values and preferences** |  |
| Referenced existing survey research on values and preferences | 24 (92.3%) |
| Referenced existing interview research on values and preferences | 1 (3.8%) |
| Engaged lay persons as part of guideline development group | 6 (23.1%) |
| Guideline development group made assumptions about values and preferences | 6 (23.1%) |
| Consulted with professional associations | 5 (19.2%) |
| Not reported | 1 (3.8%) |

**Table 3:**
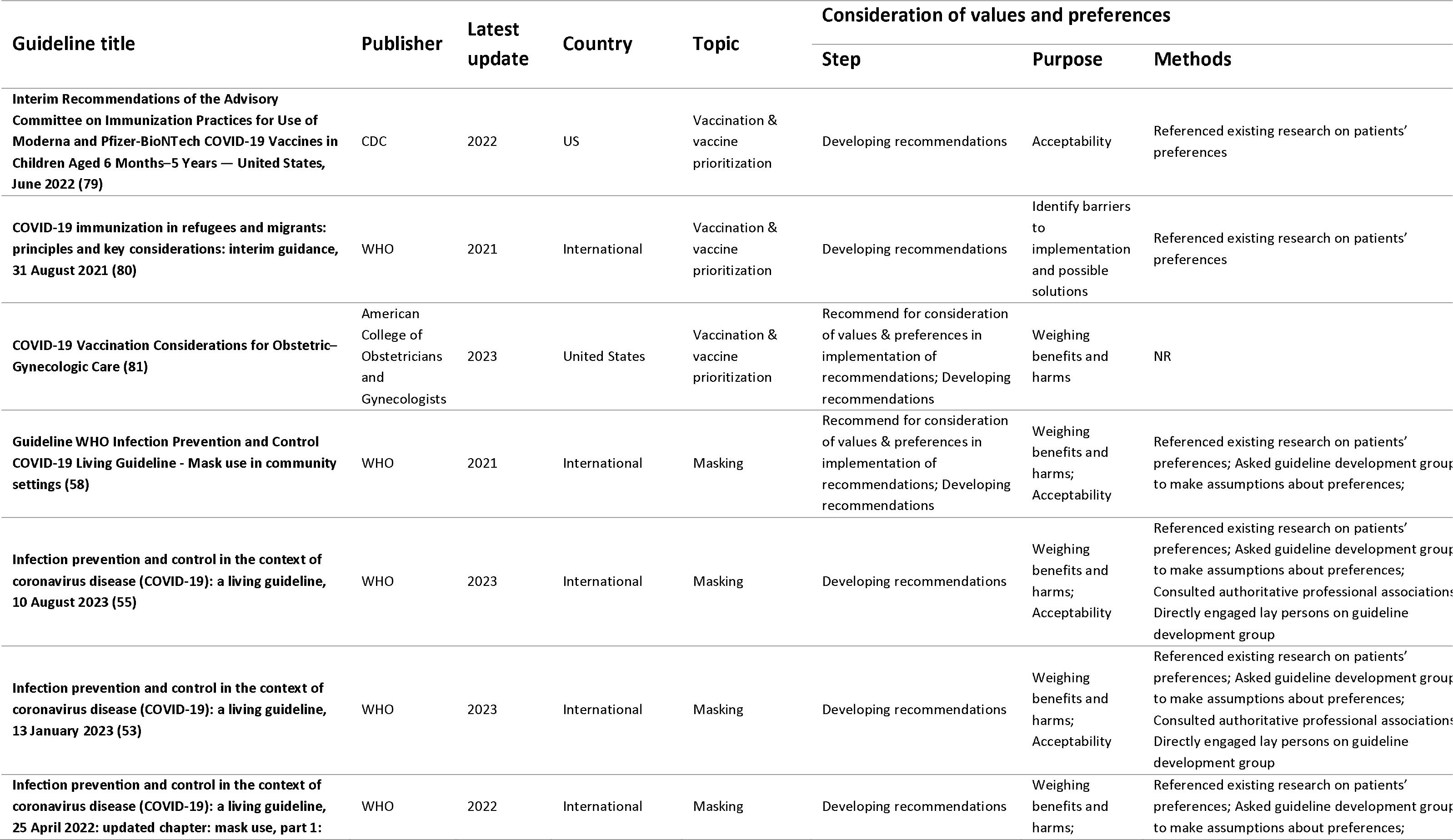
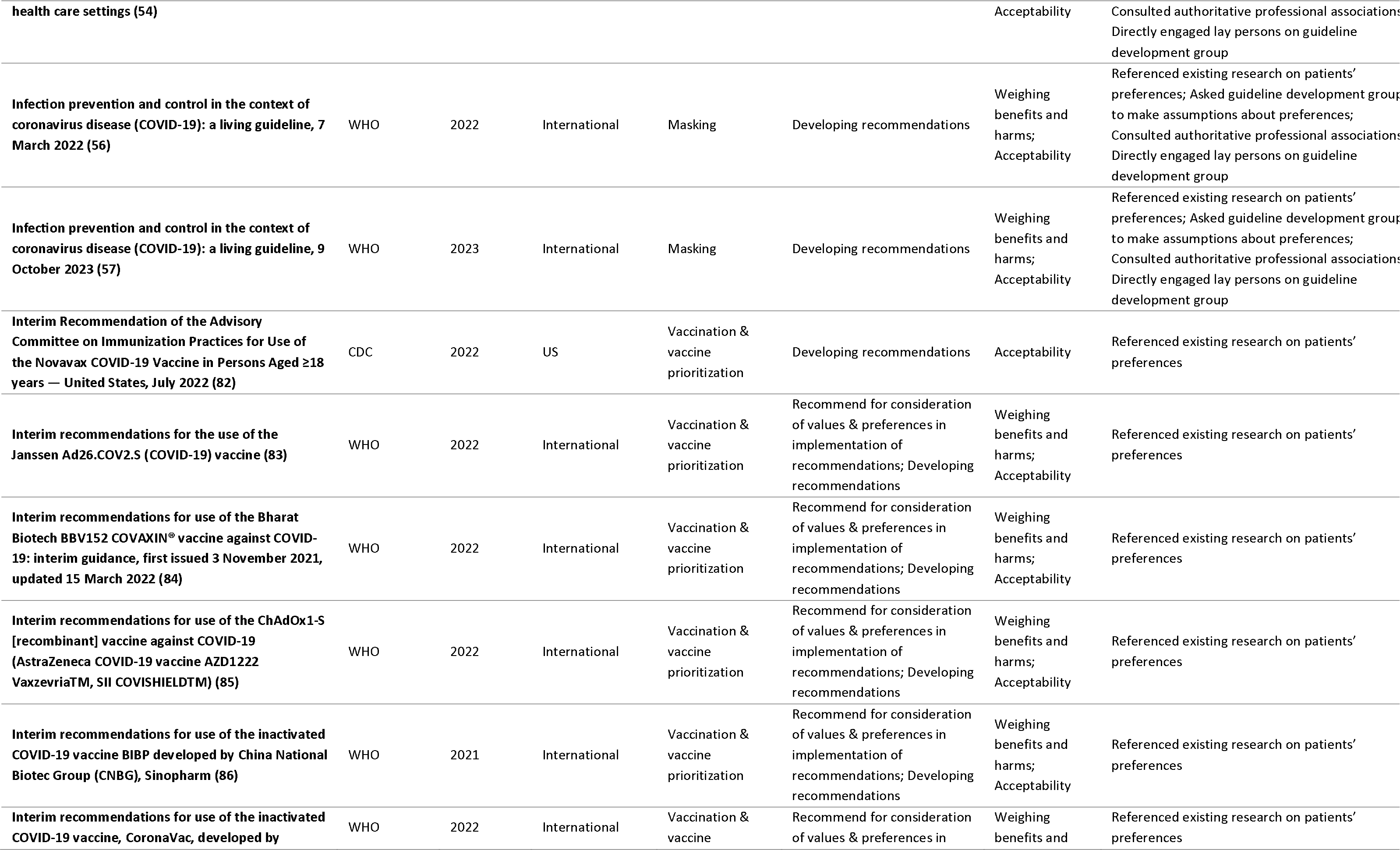
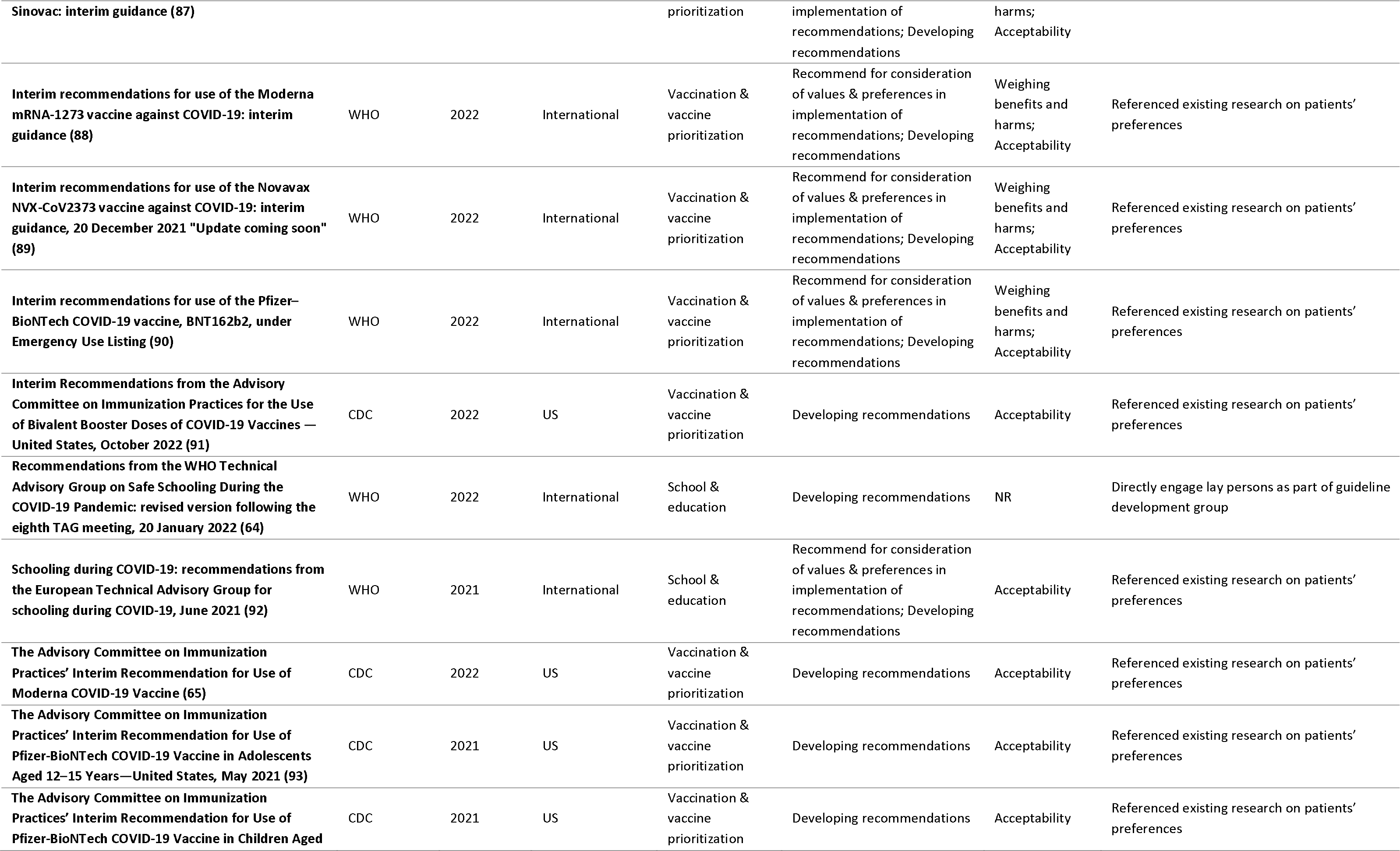
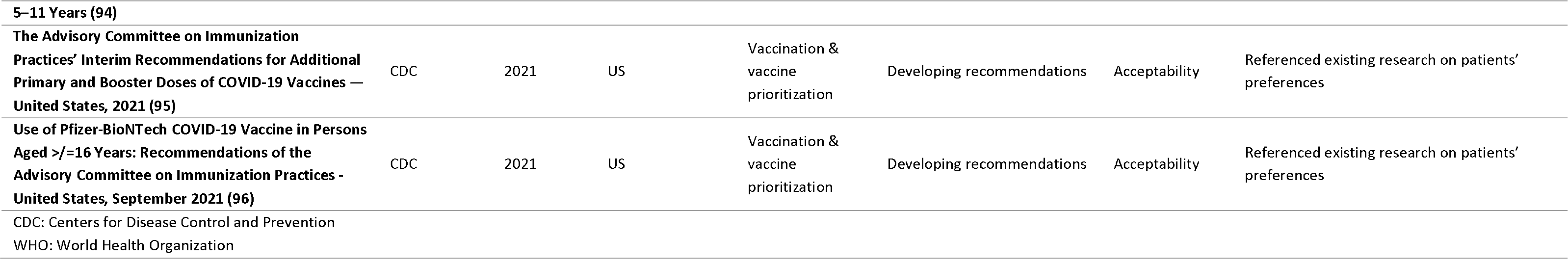
Description of guidelines that considered values and preferences.

| Guideline title | Publisher | Latest update | Country | Topic | Consideration of values and preferences |  |  |
| --- | --- | --- | --- | --- | --- | --- | --- |
|  |  |  |  |  | Step | Purpose | Methods |
| Interim Recommendations of the Advisory Committee on Immunization Practices for Use of Moderna and Pfizer-BioNTech COVID-19 Vaccines in Children Aged 6 Months–5 Years — United States, June 2022 (79) | CDC | 2022 | US | Vaccination & vaccine prioritization | Developing recommendations | Acceptability | Referenced existing research on patients’ preferences |
| COVID-19 immunization in refugees and migrants: principles and key considerations: interim guidance, 31 August 2021 (80) | WHO | 2021 | International | Vaccination & vaccine prioritization | Developing recommendations | Identify barriers to implementation and possible solutions | Referenced existing research on patients’ preferences |
| COVID-19 Vaccination Considerations for Obstetric–Gynecologic Care (81) | American College of Obstetricians and Gynecologists | 2023 | United States | Vaccination & vaccine prioritization | Recommend for consideration of values & preferences in implementation of recommendations; Developing recommendations | Weighing benefits and harms | NR |
| Guideline WHO Infection Prevention and Control COVID-19 Living Guideline - Mask use in community settings (58) | WHO | 2021 | International | Masking | Recommend for consideration of values & preferences in implementation of recommendations; Developing recommendations | Weighing benefits and harms; Acceptability | Referenced existing research on patients’ preferences; Asked guideline development group to make assumptions about preferences; |
| Infection prevention and control in the context of coronavirus disease (COVID-19): a living guideline, 10 August 2023 (55) | WHO | 2023 | International | Masking | Developing recommendations | Weighing benefits and harms; Acceptability | Referenced existing research on patients’ preferences; Asked guideline development group to make assumptions about preferences; Consulted authoritative professional associations; Directly engaged lay persons on guideline development group |
| Infection prevention and control in the context of coronavirus disease (COVID-19): a living guideline, 13 January 2023 (53) | WHO | 2023 | International | Masking | Developing recommendations | Weighing benefits and harms; Acceptability | Referenced existing research on patients’ preferences; Asked guideline development group to make assumptions about preferences; Consulted authoritative professional associations; Directly engaged lay persons on guideline development group |
| Infection prevention and control in the context of coronavirus disease (COVID-19): a living guideline, 25 April 2022: updated chapter: mask use, part 1: | WHO | 2022 | International | Masking | Developing recommendations | Weighing benefits and harms; | Referenced existing research on patients’ preferences; Asked guideline development group to make assumptions about preferences; |
| **health care settings (54)** |  |  |  |  |  | Acceptability | Consulted authoritative professional associations;<br>Directly engaged lay persons on guideline development group |
| **Infection prevention and control in the context of coronavirus disease (COVID-19): a living guideline, 7 March 2022 (56)** | WHO | 2022 | International | Masking | Developing recommendations | Weighing benefits and harms;<br>Acceptability | Referenced existing research on patients' preferences; Asked guideline development group to make assumptions about preferences; Consulted authoritative professional associations; Directly engaged lay persons on guideline development group |
| **Infection prevention and control in the context of coronavirus disease (COVID-19): a living guideline, 9 October 2023 (57)** | WHO | 2023 | International | Masking | Developing recommendations | Weighing benefits and harms;<br>Acceptability | Referenced existing research on patients' preferences; Asked guideline development group to make assumptions about preferences; Consulted authoritative professional associations; Directly engaged lay persons on guideline development group |
| **Interim Recommendation of the Advisory Committee on Immunization Practices for Use of the Novavax COVID-19 Vaccine in Persons Aged ≥18 years — United States, July 2022 (82)** | CDC | 2022 | US | Vaccination & vaccine prioritization | Developing recommendations | Acceptability | Referenced existing research on patients' preferences |
| **Interim recommendations for the use of the Janssen Ad26.COV2.S (COVID-19) vaccine (83)** | WHO | 2022 | International | Vaccination & vaccine prioritization | Recommend for consideration of values & preferences in implementation of recommendations; Developing recommendations | Weighing benefits and harms;<br>Acceptability | Referenced existing research on patients' preferences |
| **Interim recommendations for use of the Bharat Biotech BBV152 COVAXIN® vaccine against COVID-19: interim guidance, first issued 3 November 2021, updated 15 March 2022 (84)** | WHO | 2022 | International | Vaccination & vaccine prioritization | Recommend for consideration of values & preferences in implementation of recommendations; Developing recommendations | Weighing benefits and harms;<br>Acceptability | Referenced existing research on patients' preferences |
| **Interim recommendations for use of the ChAdOx1-S [recombinant] vaccine against COVID-19 (AstraZeneca COVID-19 vaccine AZD1222 Vaxzevria™, SII COVISHIELD™) (85)** | WHO | 2022 | International | Vaccination & vaccine prioritization | Recommend for consideration of values & preferences in implementation of recommendations; Developing recommendations | Weighing benefits and harms;<br>Acceptability | Referenced existing research on patients' preferences |
| **Interim recommendations for use of the inactivated COVID-19 vaccine BIBP developed by China National Biotech Group (CNBG), Sinopharm (86)** | WHO | 2021 | International | Vaccination & vaccine prioritization | Recommend for consideration of values & preferences in implementation of recommendations; Developing recommendations | Weighing benefits and harms;<br>Acceptability | Referenced existing research on patients' preferences |
| **Interim recommendations for use of the inactivated COVID-19 vaccine, CoronaVac, developed by** | WHO | 2022 | International | Vaccination & vaccine | Recommend for consideration of values & preferences in | Weighing benefits and | Referenced existing research on patients' preferences |
| **Sinovac: interim guidance (87)** |  |  |  | prioritization | implementation of recommendations; Developing recommendations | harms; Acceptability |  |
| **Interim recommendations for use of the Moderna mRNA-1273 vaccine against COVID-19: interim guidance (88)** | WHO | 2022 | International | Vaccination & vaccine prioritization | Recommend for consideration of values & preferences in implementation of recommendations; Developing recommendations | Weighing benefits and harms; Acceptability | Referenced existing research on patients' preferences |
| **Interim recommendations for use of the Novavax NVX-CoV2373 vaccine against COVID-19: interim guidance, 20 December 2021 "Update coming soon" (89)** | WHO | 2022 | International | Vaccination & vaccine prioritization | Recommend for consideration of values & preferences in implementation of recommendations; Developing recommendations | Weighing benefits and harms; Acceptability | Referenced existing research on patients' preferences |
| **Interim recommendations for use of the Pfizer–BioNTech COVID-19 vaccine, BNT162b2, under Emergency Use Listing (90)** | WHO | 2022 | International | Vaccination & vaccine prioritization | Recommend for consideration of values & preferences in implementation of recommendations; Developing recommendations | Weighing benefits and harms; Acceptability | Referenced existing research on patients' preferences |
| **Interim Recommendations from the Advisory Committee on Immunization Practices for the Use of Bivalent Booster Doses of COVID-19 Vaccines — United States, October 2022 (91)** | CDC | 2022 | US | Vaccination & vaccine prioritization | Developing recommendations | Acceptability | Referenced existing research on patients' preferences |
| **Recommendations from the WHO Technical Advisory Group on Safe Schooling During the COVID-19 Pandemic: revised version following the eighth TAG meeting, 20 January 2022 (64)** | WHO | 2022 | International | School & education | Developing recommendations | NR | Directly engage lay persons as part of guideline development group |
| **Schooling during COVID-19: recommendations from the European Technical Advisory Group for schooling during COVID-19, June 2021 (92)** | WHO | 2021 | International | School & education | Recommend for consideration of values & preferences in implementation of recommendations; Developing recommendations | Acceptability | Referenced existing research on patients' preferences |
| **The Advisory Committee on Immunization Practices' Interim Recommendation for Use of Moderna COVID-19 Vaccine (65)** | CDC | 2022 | US | Vaccination & vaccine prioritization | Developing recommendations | Acceptability | Referenced existing research on patients' preferences |
| **The Advisory Committee on Immunization Practices' Interim Recommendation for Use of Pfizer-BioNTech COVID-19 Vaccine in Adolescents Aged 12–15 Years—United States, May 2021 (93)** | CDC | 2021 | US | Vaccination & vaccine prioritization | Developing recommendations | Acceptability | Referenced existing research on patients' preferences |
| **The Advisory Committee on Immunization Practices' Interim Recommendation for Use of Pfizer-BioNTech COVID-19 Vaccine in Children Aged** | CDC | 2021 | US | Vaccination & vaccine prioritization | Developing recommendations | Acceptability | Referenced existing research on patients' preferences |
| 5–11 Years (94) |  |  |  |  |  |  |  |
| The Advisory Committee on Immunization Practices' Interim Recommendations for Additional Primary and Booster Doses of COVID-19 Vaccines — United States, 2021 (95) | CDC | 2021 | US | Vaccination & vaccine prioritization | Developing recommendations | Acceptability | Referenced existing research on patients' preferences |
| Use of Pfizer-BioNTech COVID-19 Vaccine in Persons Aged ≥/16 Years: Recommendations of the Advisory Committee on Immunization Practices - United States, September 2021 (96) | CDC | 2021 | US | Vaccination & vaccine prioritization | Developing recommendations | Acceptability | Referenced existing research on patients' preferences |
| CDC: Centers for Disease Control and Prevention<br>WHO: World Health Organization |  |  |  |  |  |  |  |

The purpose for considering values and preferences was most often to assess the acceptability of recommendations (23; 88.5%) or to weigh anticipated benefits and harms of alternative courses of action (15; 57.7%). None of the guidelines reported considering values and preferences for other purposes, like developing simple language summaries of the recommendations.

Most guidelines considered values and preferences by consulting published survey research (24; 92.3%). These guidelines did not report identifying this published research using systematic review methods. None of the guidelines performed de novo research or provided opportunity for public comment. Supplement 3 lists the studies referenced by guidelines as evidence of values and preferences.

Less than a quarter of guidelines (6; 23.1%) reported engaging lay persons as part of the guideline development group. Among guidelines that reported engaging lay persons as part of the guideline development group, none described the number of lay people consulted or their demographic characteristics (e.g., age, sex, equity deserving characteristics).

A minority of guideline development groups (6; 23.1%) reported making assumptions about public values and preferences. A minority of guidelines (5; 19.2%) reported consulting with professional associations about values and preferences.

Among guidelines that were updated throughout the course of the pandemic, we did not identify any guidelines in which values and preferences were first not considered but were later incorporated in subsequent updates.

None of the guidelines reflected on lessons learned or the advantages or disadvantages of considering public values and preferences or evaluated the quality or impact of public engagement.

Box 3 describes examples of ways in which guidelines considered values and preferences.

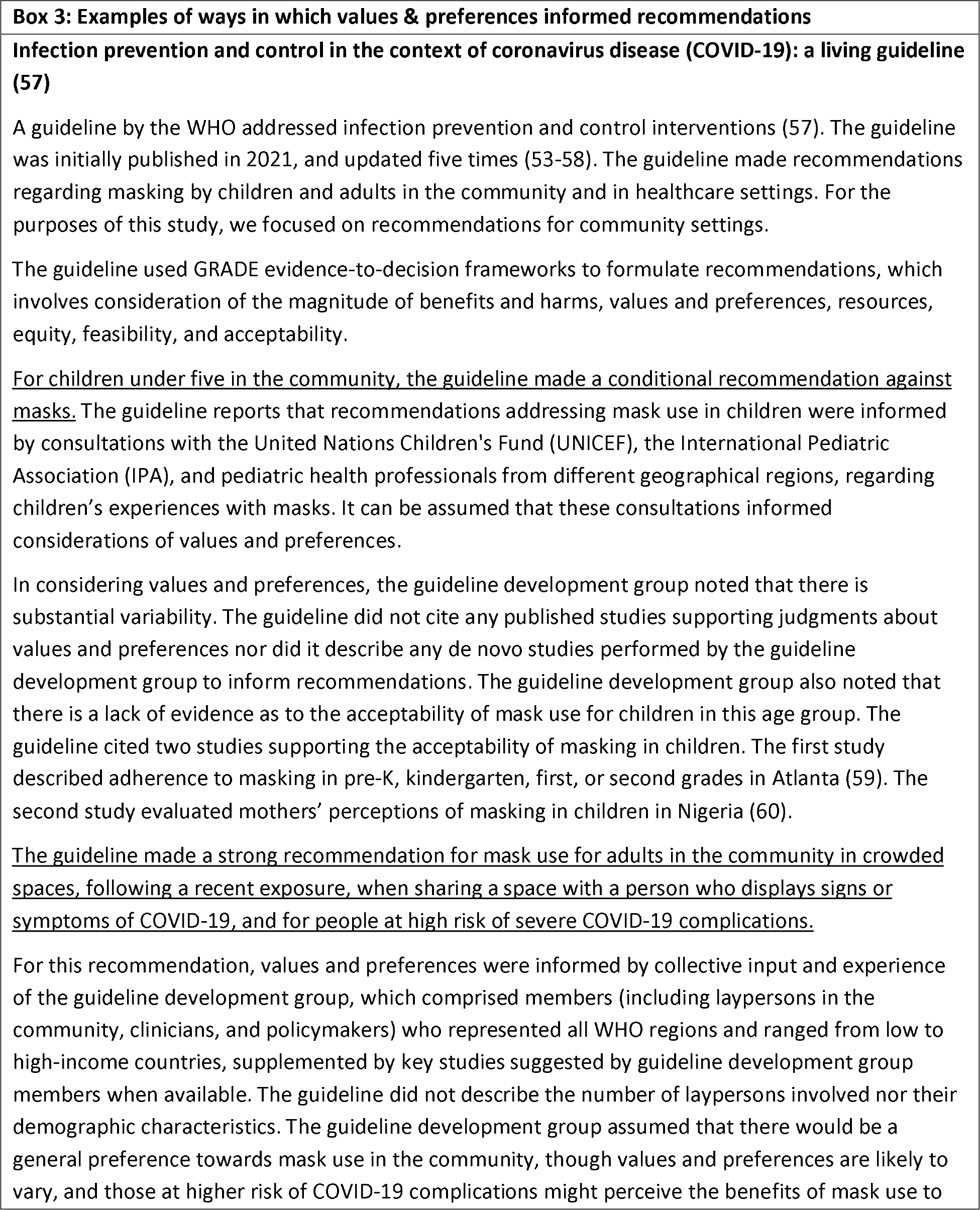
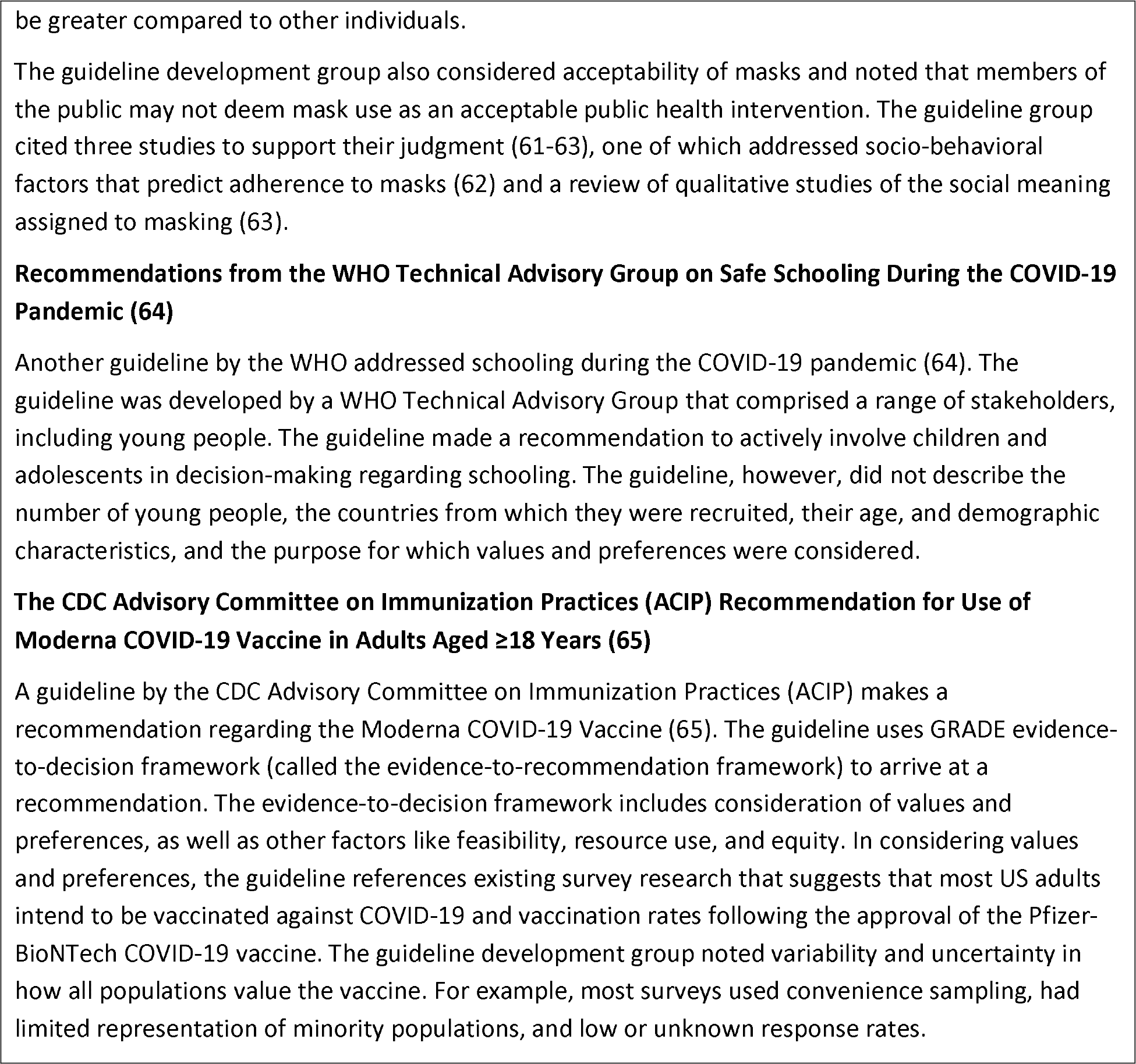

## Discussion

### Main findings

Our systematic review of 129 COVID-19 public health guidelines found that they seldom considered values and preferences. Among the few guidelines that considered values and preferences, we found limitations in how they were considered. For example, none of the included guidelines performed or referenced systematic reviews of values and preferences. Instead, they referenced select research publications without a description of how the studies were identified. Failure to perform systematic reviews risks cherry-picking studies based on their findings. Some guideline development groups made assumptions about the public’s values and preferences. This approach, though practical, may be suboptimal because the general public likely weighs pandemic policy decisions differently than professionals (31).

The pandemic forced guideline developers to produce guidelines quickly under circumstances that did not allow for typical guideline development procedures such as in-person meetings or travel. While the pressures of the pandemic may account for the suboptimal consideration of values and preferences, we show that even guidelines produced three years into the pandemic, when most of these challenges were mitigated and circumstances had normalized, did not consider values and preferences. Further, none of the guidelines explicitly acknowledged lack of or inadequate consideration of values and preferences, engagement of the public, or the use of pragmatic approaches as limitations.

Thus, other factors are more likely to explain the suboptimal consideration of values and preferences. Until recently, there has been limited consensus on the ideal approaches to involve laypersons in guideline development (11, 22, 66). For example, while methodological handbooks strongly encourage consideration of values and preferences, they contain little practical guidance (12, 21, 22). Hence, even if guideline developers may have wished to incorporate values and preferences, they may have been uncertain about how to do so effectively. Adding to this already complex process is evidence that values and preferences related to COVID-19 recommendations varied widely across regions and age groups and changed during the pandemic (67–70). Such challenges might have discouraged guideline developers from giving priority to this issue.

The omission of values and preferences could also be attributed to guideline developers’ limited familiarity with established standards for developing trustworthy guidelines. This is further evidenced by additional methodological oversights in guidelines, such as the failure to disclose the conflicts of interests of the guideline development group. Moreover, a recent study also showed that few COVID-19 recommendations considered issues related to health equity—an aspect increasingly recognized as an important component of guideline development (71). Such patterns suggest that guideline developers that did not consider values and preferences might not be fully aware of established standards for trustworthy guideline development.

### Relation to previous research

This review is the first to systematically examine how COVID-19 public health guidelines considered public values and preferences. Prior research on COVID-19 guideline quality has mainly focused on clinical practice guidelines (e.g., guidelines that advise healthcare providers on the treatment of patients with COVID-19) (29, 31, 72). Previous evidence on public engagement during COVID-19 has typically been in the form of anecdotal evidence and case studies (29, 31, 72). For example, the Scottish government hosted an online public discussion about people’s ideas and concerns around lockdown, which generated 18,000 comments from citizens (73). In the Netherlands, 30,000 citizens advised the government on relaxing lockdown measures (31). Our work systematically evaluates how and to what extent COVID-19 guidelines accounted for public values and preferences.

### Strengths and limitation

The strengths of this review include our systematic search for COVID-19 public health guidelines and rigorous collection of data on values and preferences by two reviewers independently using an established framework for consideration of values and preferences.

Although we attempted to provide a comprehensive overview of all COVID-19 public health guidelines, there remains a possibility that some eligible guidelines were excluded. We used the eCOVID-19 RecMap platform to identify guidelines—originally designed to reduce the time and resources organizations would need to invest to develop de novo guidelines and to facilitate guideline adaptation (42, 44, 45). Moreover, the eCOVID19 RecMap appears to favor guidelines from the most influential organizations that likely have the highest standards for developing guidelines (e.g., WHO, CDC). Consequently, our sample likely includes an overrepresentation of guidelines that consider values and preferences.

Likewise, although we used a broad definition and operationalization of values and preferences, it is possible that our definition and framework may not have captured guideline developers’ considerations of values and preferences (18).

To identify eligible guidelines, reviewers worked independently and in duplicate to screen all recommendations catalogued on the eCOVID-19 RecMap. Any guideline that contained at minimum one eligible recommendation was included. Given the length and complexity of guideline documents, it is possible that we may have missed guidelines that included eligible recommendations.

For feasibility, we restricted eligibility to national and international guidelines and excluded regional guidelines. It is possible for national and international guidelines to be adapted into regional settings during which local values and preferences may be considered.

Guidelines are sometimes updated as new evidence emerges and a single guideline may include several versions. We intended to include all versions of guidelines to study how consideration of values and preferences may have evolved during the pandemic. It is possible that some organizations removed access to outdated guidelines or replaced outdated guidelines on webpages, precluding their inclusion in our study.

It is possible that guidelines considered values and preferences but did not report on such considerations. For example, a survey of guideline developers showed that organizations often implicitly rather than explicitly consider values and preferences (66). Nevertheless, failure to explicitly report consideration of values and preferences reduces the guideline’s transparency and leaves readers unable to evaluate its credibility.

For select recommendations, such as good practice statements that summarize recommendations where the alternative would be absurd (e.g., a recommendation for surgeons to forego handwashing before a surgical procedure), values and preferences are unlikely to be consequential (44, 45). We, however, restricted our study to guidelines that addressed recommendations that require consideration of values and preferences. For example, recommendations addressing isolation, lockdowns, and other infection control measures necessitate weighing immediate benefits of controlling the virus against broader health and socioeconomic consequences (47, 48).

We did not review documents from guideline producing organizations outlining their typical standards and methods for developing guidelines. We anticipated that these organizations may have diverged from typical practices due to the unique context presented by the COVID-19 pandemic. Likewise, inferences on how recommendations might have differed if values and preferences were considered were beyond the scope of this study.

### Implications

As the pandemic evolves from an acute threat to a long-term public health issue, now is a critical time to reflect and learn from our response to inform future emergency preparedness efforts. While the COVID- 19 pandemic was unprecedented, we anticipate other health emergencies that will also benefit from guidelines that are aligned with societal values.

Our investigation revealed that few COVID-19 public health guidelines considered values and preferences, which may have contributed to unpopular pandemic policies and led to an erosion of trust in public health (35–38, 74–77).This erosion of trust may present a challenge for managing future crises, which may again require cohesive and coordinated efforts from the public.

We acknowledge that guideline developers and decision-makers may be reluctant to consider values and preferences in favor of relying only on scientific evidence. Considering values and preferences respects the rights of citizens to participate in health decision-making, aligns guidelines with the needs and priorities of the communities they are intended to serve, ensures recommendations are logistically feasible and acceptable, and improves support for the recommendations (13, 14). Formulating recommendations will invariably involve inherent value judgements—alternative courses of action are almost always accompanied by both benefits and harms and value judgments are necessary to determine whether benefits of a particular course of action outweigh harms or vice versa. For these reasons, all guideline producing organizations and organizations that establish standards for developing guidelines require consideration of values and preferences (2, 3, 10, 16, 78).

Our results suggest that guideline developers may not be aware of standards that necessitate consideration of values and preferences. Therefore, we recommend future efforts focus on improving guideline developers’ understanding of the importance of values and preferences in formulating recommendations.

Future research could also address guideline developers’ experiences considering values and preferences in COVID-19 guidelines. Surveys and interviews with guideline developers can provide insights on facilitators and barriers to public engagement during the pandemic and identify efficient methods for considering values and preferences in health emergencies and crisis situations. Such research will ensure that future health crises are met with public health guidelines that are aligned with core societal values.

## Conclusion

We reviewed a sample of COVID-19 public health guidelines to capture and characterize whether and how they considered public values and preferences. We found few COVID-19 public health guidelines to consider public values and preferences. Where values and preferences were considered, it was using suboptimal methods. Disregard for values and preferences might have contributed to divisive COVID-19 policies. While the COVID-19 pandemic was unprecedented, we anticipate other health emergencies that will also benefit from guidelines that are aligned with societal values. We recommend future research identify efficient methods for considering values and preferences in health emergencies and crisis situations.

## Disclaimers

None.

## Data

We will make all data publicly accessible upon the publication of this manuscript on Open Science Framework (https://osf.io/aup36/).

## Acknowledgements

None.

## Authors’ Contributions

DZ conceived the idea. SK, ML, and TJ collected data. SK and ML synthesized data and produced descriptive characteristics. SK, TP, and DZ wrote the first draft of the manuscript. All authors reviewed, provided critical feedback on the manuscript, and approved the final version of the manuscript.

