## Supplement for "Values and preferences in COVID-19 public health guidelines: A systematic review"

Dena Zeraatkar, PhD

Department of Anesthesia and Department of Health Research Methods, Evidence, and Impact

McMaster University, Hamilton, ON, Canada

### Supplement 1: Included guidelines

1.	American Academy of Pediatrics (AAP). COVID-19 Testing Guidance. December 2022.

2.	American Academy of Pediatrics (AAP). FAQs: Management of Infants Born to Mothers with Suspected or Confirmed COVID-19. June 2021.

3.	American College of Obstetricians and Gynecologists (ACOG). COVID-19 Vaccination Considerations for Obstetric–Gynecologic Care. May 2023.

4.	Australian and New Zealand Intensive Care Society COVID-19 Guidelines. October 2020.

5.	Centers for Disease Control and Prevention (CDC). Care for breastfeeding people : interim guidance on breastfeeding and breast milk feeds in the context of COVID-19. February 2022.

6.	Centers for Disease Control and Prevention (CDC). Cleaning and Disinfecting Your Facility - Every Day and When Someone is Sick. Centers for Disease Control and Prevention. November 2022.

7.	Centers for Disease Control and Prevention (CDC). Contact Tracing for COVID-19. February 2022.

8.	Centers for Disease Control and Prevention (CDC). COVID-19 and Cooling Centers Interim guidance to reduce the risk of introducing and transmitting SARS COV-2 (the agent responsible for causing COVID-19 disease) in cooling centers. April 2021.

9.	Centers for Disease Control and Prevention (CDC). Guidance and tips for tribal community living during COVID-19. August 2022.

10.	Centers for Disease Control and Prevention (CDC). Guidance for Adult Day Services Centers. July 2021.

11.	Centers for Disease Control and Prevention (CDC). Guidance for General Population Disaster Shelters During the COVID-19 Pandemic. September 2022.

12.	Centers for Disease Control and Prevention (CDC). Guidance for institutions of higher education (IHEs). July 2021.

13.	Centers for Disease Control and Prevention (CDC). Guidance on Management of COVID-19 in Homeless Service Sites and in Correctional and Detention Facilities. May 2023.

14.	Centers for Disease Control and Prevention (CDC). Interim Guidance for Homeless Service Providers to Plan and Respond to Coronavirus Disease 2019 (COVID-19). May 2023.

15.	Centers for Disease Control and Prevention (CDC). Investigating and responding to COVID-19 cases at homeless service provider sites. May 2023.

16.	Centers for Disease Control and Prevention (CDC). Isolation and Precautions for People with COVID-19. May 2023.

17.	Centers for Disease Control and Prevention (CDC). Operational Guidance for K-12 Schools and Early Care and Education Programs to Support Safe In-Person Learning. May 2023.

18.	Centers for Disease Control and Prevention (CDC). Prioritizing Case Investigations and Contact Tracing for COVID-19. February 2022.

19.	Centers for Disease Control and Prevention (CDC). Use and Care of Masks. May 2023.

25.	European Centre for Disease Prevention and Control (ECDC). Considerations for the use of face masks in the community in the context of the SARS-CoV-2 Omicron variant of concern. February 2022.

26.	European Centre for Disease Prevention and Control (ECDC). Contact tracing in the European Union: public health management of persons, including healthcare workers, who have had contact with COVID-19 cases – fourth update. October 2021.

27.	European Centre for Disease Prevention and Control (ECDC). COVID-19 Aviation Health Safety Protocol: Operational guidelines for the management of air passengers and aviation personnel in relation to the COVID-19 pandemic. May 2022.

28.	European Centre for Disease Prevention and Control (ECDC). COVID-19: EU guidance for cruise ship operations. May 2021.

29.	European Centre for Disease Prevention and Control (ECDC). Disinfection of environments in healthcare and non-healthcare settings potentially contaminated with SARS-CoV-2. March 2020.

30.	European Centre for Disease Prevention and Control (ECDC). Guidance for COVID-19 quarantine and testing of travellers. March 2021.

31.	European Centre for Disease Prevention and Control (ECDC). Guidance for COVID-19 testing and quarantine of air travellers. December 2020.

32.	European Centre for Disease Prevention and Control (ECDC). Guidance for the prevention and control of COVID-19 in temporary reception centres in the context of the large numbers of people fleeing Ukraine. March 2022.

33.	European Centre for Disease Prevention and Control (ECDC). Guidance on ending the isolation period for people with COVID-19, third update. January 2022.

34.	European Centre for Disease Prevention and Control (ECDC). Guidelines for the implementation of non-pharmaceutical interventions against COVID-19. September 2020.

35.	European Centre for Disease Prevention and Control (ECDC). Guidelines in response to the worsening of the epidemiological situation - Addendum to the Aviation Health Safety Protocol. January 2023.

36.	European Centre for Disease Prevention and Control (ECDC). Heating, ventilation and air-conditioning systems in the context of COVID-19: first update. November 2020.

37.	European Centre for Disease Prevention and Control (ECDC). Interim guidance on the benefits of full vaccination against COVID-19 for transmission and implications for non-pharmaceutical interventions. April 2021.

38.	European Centre for Disease Prevention and Control (ECDC). Operational public health considerations for the prevention and control of infectious diseases in the context of Russia’s aggression towards Ukraine. March 2022.

39.	European Centre for Disease Prevention and Control (ECDC). Preliminary public health considerations for COVID-19 vaccination strategies in the second half of 2022. July 2022.

40.	Fleming-Dutra KE, Wallace M, Moulia DL, Twentyman E, Roper LE, Hall E, et al. Interim Recommendations of the Advisory Committee on Immunization Practices for Use of Moderna and Pfizer-BioNTech COVID-19 Vaccines in Children Aged 6 Months–5 Years — United States, June 2022. Centers for Disease Control and Prevention (CDC). June 2022;71(26):859-68.

41.	Ghate S, Zadey S, Thapar RK, Shah D, Basavaraja GV, Kamath SS, et al. Indian Academy of Pediatrics Revised Guidelines on School Reopening: First Revision, September 2021. Indian Academy of Pediatrics (IAP). September 2021;58(10):959-61.

42.	Government of Ireland. COVID-19 Response Plan for the safe and sustainable reopening of Post Primary Schools. August 2021.

43.	Government of Ireland. COVID-19 Response Plan for the safe and sustainable reopening of Primary and Special Schools. August 2021.

44.	Government of Ireland. Transitional Protocol: Good Practice Guidance for Continuing to Prevent the Spread of COVID-19. January 2022.

45.	Harvard’s Edmond J. Safra Center for Ethics HGHI. Schools and the Path to Zero: Strategies for Pandemic Resilience in the Face of High Community Spread. December 2020.

46.	Johansen TB, Astrup E, Jore S, Nilssen H, Dahlberg BB, Klingenberg C, et al. Infection prevention guidelines and considerations for paediatric risk groups when reopening primary schools during COVID-19 pandemic, Norway, April 2020. Norwegian Institute of Public Health, European Public Health Microbiology Training Programme (EUPHEM), European Centre for Disease Prevention and Control (ECDC), Norwegian Directorate for Education and Training, Paediatric Research Group, Faculty of Health Sciences, University of Tromsø-Arctic University of Norway, Department of Paediatrics and Adolescence Medicine, University Hospital of North Norway. April 2020;25(22).

47.	Kalinka J, Wielgos M, Leszczynska-Gorzelak B, Piekarska A, Huras H, Sieroszewski P, et al. COVID-19 impact on perinatal care: risk factors, clinical manifestation and prophylaxis. Polish experts’ opinion for December 2020. Ginekol Pol. January 2021;92(1):57-63.

48.	Kasi SG, Dhir SK, Shah A, Shivananda S, Verma S, Marathe S, et al. Coronavirus Disease 2019 (COVID-19) Vaccination for Children: Position Statement of Indian Academy of Pediatrics Advisory Committee on Vaccination and Immunization Practices. Indian Academy of Pediatrics (IAP). December 2021;59(1):51-7.

49.	Kasi SG, Dhir SK, Shivananda S, Marathe S, Chatterjee K, Agarwalla S, et al. Breastfeeding and Coronavirus Disease 2019 (COVID-19) Vaccination: Position Statement of Indian Academy of Pediatrics Advisory Committee on Vaccination and Immunization Practices. Indian Academy of Pediatrics (IAP), Indian Academy of Pediatrics Advisory Committee on Vaccination and Immunization Practices (IAP/ACVIP). July 2021;58(7):647-9.

50.	Mbaeyi S, Oliver SE, Collins JP, Godfrey M, Goswami ND, Hadler SC, Jones J, Moline H, Moulia D, Reddy S, Schmit K, Wallace M, Chamberland M, Campos-Outcalt D, Morgan RL, Bell BP, Brooks O, Kotton C, Talbot HK, Lee G, Daley MF, Dooling K. The Advisory Committee on Immunization Practices’ Interim Recommendations for Additional Primary and Booster Doses of COVID-19 Vaccines — United States, 2021. Centers for Disease Control and Prevention (CDC). November 2021;70.

51.	Oliver SE, Wallace M, See I, Mbaeyi S, Godfrey M, Hadler SC, Jatlaoui T, Twentyman E, Hughes MM, Rao A, Fiore A, Su JR, Broder K, Shimabukuro T, Lale A, Shay DK, Markowitz LE, Wharton M, Bell BP, Brooks O, McNally V, Lee GM, Talbot HK, Daley MF. Use of the Janssen (Johnson & Johnson) COVID-19 Vaccine: Updated Interim Recommendations from the Advisory Committee on Immunization Practices — United States, December 2021. Centers for Disease Control and Prevention (CDC). January 2022;71:90–5.

52.	Pilato TC, Taki FA, Kaur G. Safeguarding pregnant asylum-seekers and refugees during the era of COVID-19. Weill Cornell's Department of Anesthesiology. January 2021;11.

53.	Public Health Agency of Canada (PHAC). Adjusting public health measures in the context of COVID-19 vaccination. October 2021.

54.	Public Health Agency of Canada (PHAC). At home: Using ventilation and filtration to reduce the risk of aerosol transmission of COVID-19. January 2023.

55.	Public Health Agency of Canada (PHAC). COVID-19 and people with disabilities in Canada. January 2023.

56.	Public Health Agency of Canada (PHAC). COVID-19 mask use: Advice for community settings. January 2023.

57.	Public Health Agency of Canada (PHAC). COVID-19 vaccines: Canadian Immunization Guide. June 2023.

58.	Public Health Agency of Canada (PHAC). COVID-19: Guidance on indoor ventilation during the pandemic. July 2022.

59.	Public Health Agency of Canada (PHAC). Guidance for a strategic approach to lifting restrictive public health measures. June 2022.

60.	Public Health Agency of Canada (PHAC). Infection prevention and control for COVID-19: Interim guidance for home care setting. January 2022.

61.	Public Health Agency of Canada (PHAC). National surveillance for Coronavirus disease (COVID-19). March 2021.

62.	Public Health Agency of Canada (PHAC). Planning guidance for administration of COVID-19 vaccine. December 2020.

63.	Public Health Agency of Canada (PHAC). Public health management of cases and contacts associated with COVID-19. December 2021.

64.	Public Health Agency of Canada (PHAC). Rapid response: Guidance on the use of booster COVID-19 vaccine doses in adolescents 12-17 years of age. January 2022.

65.	Public Health Agency of Canada (PHAC). Update with consideration of Omicron – Interim COVID-19 infection prevention and control in the health care setting when COVID-19 is suspected or confirmed– December 23, 2021. December 2021.

66.	Rosenblum HG, Wallace M, Godfrey M, Roper LE, Hall E, Fleming-Dutra KE, et al. Interim Recommendations from the Advisory Committee on Immunization Practices for the Use of Bivalent Booster Doses of COVID-19 Vaccines — United States, October 2022. Centers for Disease Control and Prevention (CDC). November 2022;71(45):1436-41.

67.	Royal College of Physicians of Ireland. 2023 COVID-19 Vaccination Strategy Recommendations. April 2023.

68.	Twentyman E, Wallace M, Roper LE, Anderson TC, Rubis AB, Fleming-Dutra KE, et al. Interim Recommendation of the Advisory Committee on Immunization Practices for Use of the Novavax COVID-19 Vaccine in Persons Aged ≥18 years — United States, July 2022. Centers for Disease Control and Prevention (CDC). August 2022;71(31):988-92.

69.	UK Health Security Agency. People with symptoms of a respiratory infection including COVID-19. June 2022.

70.	United Nations International Children's Emergency Fund (UNICEF) WHOW, International Federation of Red Cross and Red Crescent Societies (IFRC). Interim Guidance for COVID-19 Prevention and Control in Schools. March 2020.

71.	United Nations International Children's Emergency Fund WHO, International Federation of Red Cross and Red Crescent Societies. Updated Interim Guidance for COVID-19 Prevention and Control in Schools V2. August 2022.

72.	Wallace M, Moulia D, Blain AE, Ricketts EK, Minhaj FS, Link-Gelles R, et al. The Advisory Committee on Immunization Practices’ Recommendation for Use of Moderna COVID-19 Vaccine in Adults Aged ≥18 Years and Considerations for Extended Intervals for Administration of Primary Series Doses of mRNA COVID-19 Vaccines — United States, February 2022. Centers for Disease Control and Prevention (CDC). February 2022;71(11):416-21.

73.	Wallace M, Woodworth KR, Gargano JW, Scobie HM, Blain AE, Moulia D, et al. The Advisory Committee on Immunization Practices’ Interim Recommendation for Use of Pfizer-BioNTech COVID-19 Vaccine in Adolescents Aged 12–15 Years — United States, May 2021. Centers for Disease Control and Prevention (CDC). May 2021;70(20):749-52.

74.	Woodworth KR, Moulia D, Collins JP, Hadler SC, Jones JM, Reddy SC, et al. The Advisory Committee on Immunization Practices’ Interim Recommendation for Use of Pfizer-BioNTech COVID-19 Vaccine in Children Aged 5–11 Years — United States, November 2021. Centers for Disease Control and Prevention (CDC). November 2021;70(45):1579-83.

75.	World Health Organization (WHO). Actions for consideration in the care and protection of vulnerable population groups for COVID-19. October 2021.

76.	World Health Organization (WHO). Cleaning and disinfection of environmental surfaces in the context of COVID-19. May 2020.

77.	World Health Organization (WHO). Conducting community engagement for COVID-19 vaccines: interim guidance, 31 January 2021. January 2021.

78.	World Health Organization (WHO). Considerations for COVID-19 surveillance for vulnerable populations. September 2021.

79.	World Health Organization (WHO). Considerations for implementing and adjusting public health and social measures in the context of COVID-19. March 2023.

80.	World Health Organization (WHO). Considerations in the investigation of cases and clusters of COVID-19. October 2020.

81.	World Health Organization (WHO). Considerations to relax border restrictions in the Western Pacific Region. June 2020.

82.	World Health Organization (WHO). Contact tracing and quarantine in the context of COVID-19: interim guidance, 6 July 2022. July 2022.

83.	World Health Organization (WHO). COVID-19 immunization in refugees and migrants: principles and key considerations: interim guidance, 31 August 2021. August 2021.

84.	World Health Organization (WHO). COVID-19 management in hotels and other entities of the accommodation sector interim guidance, 25 August 2020. August 2020.

85.	World Health Organization (WHO). The COVID-19 pandemic in the Eastern Mediterranean Region. October 2020.

86.	World Health Organization (WHO). COVID-19 Strategic Preparedness and Response Plan Operational Planning Guideline. January 2022.

87.	World Health Organization (WHO). Critical preparedness, readiness and response actions for COVID-19. November 2020.

88.	World Health Organization (WHO). Disaster evacuation shelters in the context of COVID-19. July 2022.

89.	World Health Organization (WHO). European Technical Advisory Group of Experts on Immunization (ETAGE) interim recommendations, June 2021. June 2021.

90.	World Health Organization (WHO). Framework for decision-making: implementation of mass vaccination campaigns in the context of COVID-19. May 2020.

91.	World Health Organization (WHO). Guidance for surveillance of SARS-CoV-2 variants: Interim guidance, 9 August 2021. August 2021.

92.	World Health Organization (WHO). Guidance on COVID-19 for the care of older people and people living in long-term care facilities, other non-acute care facilities and home care. March 2022.

93.	World Health Organization (WHO). Guidance on operational microplanning for COVID-19 vaccination. November 2021.

94.	World Health Organization (WHO). Guideline WHO Infection Prevention and Control COVID-19 Living Guideline - Mask use in community settings. December 2021.

95.	World Health Organization (WHO). Guiding principles for immunization activities during the COVID-19 pandemic interim guidance, 26 March 2020. March 2020.

96.	World Health Organization (WHO). Infection prevention and control guidance for long-term care facilities in the context of COVID-19 interim guidance, 8 January 2021. January 2021.

97.	World Health Organization (WHO). Infection prevention and control in the context of coronavirus disease (‎COVID-19)‎: a living guideline, 7 March 2022. March 2022.

98.	World Health Organization (WHO). Infection prevention and control in the context of coronavirus disease (‎COVID-19)‎: a living guideline, 9 October 2023. October 2023

99.	World Health Organization (WHO). Infection prevention and control in the context of coronavirus disease (‎COVID-19)‎: a living guideline, 10 August 2023. August 2023.

100.	World Health Organization (WHO). Infection prevention and control in the context of coronavirus disease (COVID-19): a living guideline, 13 January 2023. January 2023.

101.	World Health Organization (WHO). Infection prevention and control in the context of coronavirus disease (‎COVID-19)‎: a living guideline, 25 April 2022: updated chapter: mask use, part 1: health care settings. April 2022.

102.	World Health Organization (WHO). Infection prevention and control in the household management of people with suspected or confirmed coronavirus disease (COVID-19). March 2020.

103.	World Health Organization (WHO). Interim recommendations for the use of the Janssen Ad26.COV2.S (COVID-19) vaccine. June 2022.

104.	World Health Organization (WHO). Interim recommendations for use of the Bharat Biotech BBV152 COVAXIN® vaccine against COVID-19: interim guidance, first issued 3 November 2021, updated 15 March 2022. March 2022.

105.	World Health Organization (WHO). Interim recommendations for use of the ChAdOx1-S [recombinant] vaccine against COVID-19 (AstraZeneca COVID-19 vaccine AZD1222 Vaxzevria™, SII COVISHIELD™). March 2022.

106.	World Health Organization (WHO). Interim recommendations for use of the inactivated COVID-19 vaccine BIBP developed by China National Biotec Group (‎CNBG)‎, Sinopharm. May 2021.

107.	World Health Organization (WHO). Interim recommendations for use of the inactivated COVID-19 vaccine, CoronaVac, developed by Sinovac. March 2022.

108.	World Health Organization (WHO). Interim recommendations for use of the Moderna mRNA-1273 vaccine against COVID-19. August 2022.

109.	World Health Organization (WHO). Interim recommendations for use of the Novavax NVXCoV2373 vaccine against COVID-19. September 2022.

110.	World Health Organization (WHO). Interim recommendations for use of the Pfizer–BioNTech COVID-19 vaccine, BNT162b2, under Emergency Use Listing. January 2022.

111.	World Health Organization (WHO). Operational considerations for case management of COVID-19 in health facility and community. March 2020.

112.	World Health Organization (WHO). Operational guidance: evidence-based decision-making process for developing national COVID-19 vaccination strategies, January 2021. January 2021.

113.	World Health Organization (WHO). Operational guide for engaging communities in contact tracing. May 2021.

114.	World Health Organization (WHO). Preparedness, prevention and control of coronavirus disease (COVID-19) for refugees and migrants in non-camp settings. April 2020.

115.	World Health Organization (WHO). Preparedness, prevention and control of COVID-19 in prisons and other places of detention. February 2021.

116.	World Health Organization (WHO). Public health considerations for elections and related activities in the context of the COVID-19 pandemic. December 2020.

117.	World Health Organization (WHO). Public health surveillance for COVID-19: interim guidance. February 2022.

118.	World Health Organization (WHO). Recommendations for national SARS-CoV-2 testing strategies and diagnostic capacities. June 2021.

119.	World Health Organization (WHO). Recommendations from the WHO Technical Advisory Group on Safe Schooling During the COVID-19 Pandemic: revised version following the eighth TAG meeting, 20 January 2022. January 2022.

120.	World Health Organization (WHO). Recommendations to Member States to improve hand hygiene practices to help prevent the transmission of the COVID-19 virus. April 2020.

121.	World Health Organization (WHO). Responding to community spread of COVID-19. March 2020.

122.	World Health Organization (WHO). Safe Eid al Adha practices in the context of COVID-19: Interim guidance. July 2021.

123.	World Health Organization (WHO). Safe Ramadan practices in the context of COVID-19, interim guidance. April 2021.

124.	World Health Organization (WHO). Schooling during COVID-19: recommendations from the European Technical Advisory Group for schooling during COVID-19, June 2021. June 2021.

125.	World Health Organization (WHO). Strengthening the health systems response to COVID-19: technical guidance #6: preventing and managing the COVID-19. May 2020.

126.	World Health Organization (WHO). Technical considerations for implementing a risk-based approach to international travel in the context of COVID-19: Interim guidance, 2 July 2021. July 2021.

127.	World Health Organization (WHO). Technical Note to WHO AFRO Member States on the Shortening of the Quarantine Period for Contacts of COVID-19 Cases. March 2022.

128.	World Health Organization (WHO). WHO high-level mission to North Macedonia on coronavirus disease 2019 (COVID-19) 23–25 June 2020. June 2020.

129.	World Health Organization (WHO). WHO SAGE Roadmap for prioritizing uses of COVID-19 vaccines: An approach to optimize the global impact of COVID-19 vaccines, based on public health goals, global and national equity, and vaccine access and coverage scenarios. January 2022.

### Supplement 2: Excluded guidelines and reasons for exclusion

| **Guideline Title** | **Reason for Exclusion** | **Year** | **Publisher** | **Link** |
| --- | --- | --- | --- | --- |
| COVID-19-associated acute kidney injury: consensus report of the 25th Acute Disease Quality Initiative (ADQI) Workgroup. | Does not include public health recommendations | 2020 | Acute Disease Quality Initiative (ADQI) Workgroup | https://www.nature.com/articles/s41581-020-00356-5.pdf |
| A multidisciplinary approach to triage patients with breast disease during the COVID-19 pandemic: Experience from a tertiary care center in the developing world | Does not include public health recommendations | 2021 | Aga Khan University | https://onlinelibrary.wiley.com/doi/epdf/10.1002/cnr2.1309 |
| Expert recommendation for novel severe acute respiratory syndrome coronavirus 2 (SARS-CoV-2) vaccination in patients with HIV infection | Does not include public health recommendations | 2021 | AIDS and Hepatitis C Professional Group, Chinese Society of Infectious Diseases | https://rs.yiigle.com/CN112138202107/1327551.htm |
| Alberta Health Services - COVID-19 Scientific Advisory Group Rapid Evidence Report | Regional guideline | 2021 | Alberta Health Services | https://www.albertahealthservices.ca/assets/info/ppih/if-ppih-covid-19-sag-rapid-evidence-report-masking-guidance-healthcare-workers.pdf |
| COVID-19 Scientific Advisory Group Rapid Evidence Report | Regional guideline | 2021 | Alberta Health Services | https://www.albertahealthservices.ca/assets/info/ppih/if-ppih-covid-19-sag-rapid-review-management-post-covid-conditions.pdf |
| Rehabilitation & Allied Health Practice Considerations Post - COVID-19 | Regional guideline | 2022 | Alberta Health Services | https://www.albertahealthservices.ca/topics/Page17540.aspx |
| Practical recommendations for the allergological risk assessment of the COVID-19 vaccination – a harmonized statement of allergy centers in Germany | Does not include public health recommendations | 2021 | Allergologie select | https://www.ncbi.nlm.nih.gov/pmc/articles/PMC7841415/pdf/allergologieselect-5-072.pdf |
| The Risk of Allergic Reaction to SARS-CoV-2 Vaccines and Recommended Evaluation and Management: A Systematic Review, Meta-Analysis, GRADE Assessment, and International Consensus Approach. | Does not include public health recommendations | 2021 | American Academy of Allergy, Asthma & Immunology | https://www.jaci-inpractice.org/article/S2213-2198(21)00671-1/fulltext |
| Post-COVID-19 Conditions in Children and Adolescents | Does not include public health recommendations | 2022 | American Academy of Pediatrics | https://www.aap.org/en/pages/2019-novel-coronavirus-covid-19-infections/clinical-guidance/post-covid-19-conditions-in-children-and-adolescents/ |
| American Association for the Study of Liver Diseases Expert Panel Consensus Statement: Vaccines to Prevent Coronavirus Disease 2019 Infection in Patients With Liver Disease | Does not include public health recommendations | 2021 | American Association for the Study of Liver Diseases (ASLD) | https://aasldpubs.onlinelibrary.wiley.com/doi/10.1002/hep.31751 |
| Interpreting COVID-19 Test Results in Clinical Settings: It Depends! | Does not include public health recommendations | 2021 | American Board of Family Medicine | https://www.jabfm.org/content/jabfp/34/Supplement/S233.full-text.pdf |
| 2022 ACC Expert Consensus Decision Pathway on Cardiovascular Sequelae of COVID-19 in Adults: Myocarditis and Other Myocardial Involvement, Post-Acute Sequelae of SARS-CoV-2 Infection, and Return to Play | Does not include public health recommendations | 2022 | American College of Cardiology (ACC) | https://www.sciencedirect.com/science/article/pii/S0735109722003060?via%3Dihub |
| Prevention, diagnosis, and treatment of VTE in patients with coronavirus disease 2019: CHEST guideline and Expert Panel report | Does not include public health recommendations | 2020 | American College of Chest Physicians (CHEST) | https://journal.chestnet.org/action/showPdf?pii=S0012-3692%2820%2931625-1 |
| Thromboprophylaxis in Patients with COVID-19. A Brief Update to the CHEST Guideline and Expert Panel Report | Does not include public health recommendations | 2022 | American College of Chest Physicians (CHEST) | https://journal.chestnet.org/article/S0012-3692(22)00250-1/pdf |
| From the trenches: inpatient management of coronavirus disease 2019 in pregnancy. | Does not include public health recommendations | 2020 | American College of Obstetricians and Gynecologists (ACOG) | https://www.ncbi.nlm.nih.gov/pmc/articles/PMC7294275/ |
| Update Alert: Should Remdesivir Be Used for the Treatment of Patients With COVID-19? Rapid, Living Practice Points From the American College of Physicians (Version 2) | Does not include public health recommendations | 2021 | American College of Physicians (ACP) | https://www.ncbi.nlm.nih.gov/pmc/articles/PMC8297419/ |
| What Is the Antibody Response and Role in Conferring Natural Immunity After SARS-CoV-2 Infection? Rapid, Living Practice Points From the American College of Physicians (Version 2) | Does not include public health recommendations | 2022 | American College of Physicians (ACP) | https://www.ncbi.nlm.nih.gov/pmc/articles/PMC8803138/ |
| American College of Rheumatology Clinical Guidance for Multisystem Inflammatory Syndrome in Children Associated With SARS-CoV-2 and Hyperinflammation in Pediatric COVID-19: Version 3 | Does not include public health recommendations | 2022 | American College of Rheumatology | https://onlinelibrary.wiley.com/doi/10.1002/art.42062 |
| American College of Rheumatology Guidance for COVID-19 Vaccination in Patients With Rheumatic and Musculoskeletal Diseases: Version 4 | Does not include public health recommendations | 2022 | American College of Rheumatology | https://pubmed.ncbi.nlm.nih.gov/35474640/ |
| AGA Rapid Review and Guideline for SARS-CoV2 Testing and Endoscopy Post-Vaccination: 2021 Update | Does not include public health recommendations | 2021 | American Gastroenterological Association (AGA) | https://linkinghub.elsevier.com/retrieve/pii/S0016-5085(21)03029-8 |
| Patient management and clinical recommendations during the coronavirus | Does not include public health recommendations | 2022 | American Society for Reproductive Medicine (ASRM) | https://www.asrm.org/news-and-publications/news-and-research/press-releases-and-bulletins/asrm-covd-19-task-force-issues-update-no.-20/ |
| Guidelines for COVID-19 management in hematopoietic cell transplant and cellular therapy patients | Does not include public health recommendations | 2020 | American Society for Transplantation and Cellular Therapy | https://www.ncbi.nlm.nih.gov/pmc/articles/PMC7386267/ |
| Advanced Pulmonary and Cardiac Support of COVID-19 Patients: Emerging Recommendations From ASAIO—A “Living Working Document” | Does not include public health recommendations | 2020 | American Society of Artificial Internal Organs | https://journals.lww.com/asaiojournal/FullText/2020/06000/Advanced_Pulmonary_and_Cardiac_Support_of_COVID_19.2.aspx |
| ASE statement on adapting pediatric, fetal, and congenital heart disease echocardiographic services to the evolving COVID-19 pandemic | Does not include public health recommendations | 2021 | American Society of Echocardiography | https://www.onlinejase.com/article/S0894-7317(21)00027-4/fulltext?dgcid=raven_jbs_aip_email |
| American Society of Hematology living guidelines on the use of anticoagulation for thromboprophylaxis in patients with COVID-19: January 2022 update on the use of therapeutic- intensity anticoagulation in acutely ill patients | Does not include public health recommendations | 2022 | American Society of Hematology (ASH) | https://ashpublications.org/bloodadvances/article/doi/10.1182/bloodadvances.2022007561/485124/American-Society-of-Hematology-living-guidelines |
| American Society of Hematology living guidelines on the use of anticoagulation for thromboprophylaxis in patients with COVID-19: July 2021 update on post-discharge thromboprophylaxis | Does not include public health recommendations | 2021 | American Society of Hematology (ASH) | https://ashpublications.org/bloodadvances/article/doi/10.1182/bloodadvances.2021005945/477753/American-Society-of-Hematology-living-guidelines |
| American Society of Hematology living guidelines on the use of anticoagulation for thromboprophylaxis in patients with COVID-19: March 2022 update on the use of anticoagulation in critically ill patients | Does not include public health recommendations | 2022 | American Society of Hematology (ASH) | https://ashpublications.org/bloodadvances/article/doi/10.1182/bloodadvances.2022007940/485690/American-Society-of-Hematology-living-guidelines |
| American Society of Hematology living guidelines on the use of anticoagulation for thromboprophylaxis in patients with COVID-19: May 2021 update on the use of intermediate intensity anticoagulation in critically ill patients | Does not include public health recommendations | 2021 | American Society of Hematology (ASH) | https://ashpublications.org/bloodadvances/article-split/doi/10.1182/bloodadvances.2021005493/476770/American-Society-of-Hematology-living-guidelines |
| Use of Anticoagulation in Patients with COVID-19 | Does not include public health recommendations | 2020 | American Society of Hematology (ASH) | https://www.hematology.org/education/clinicians/guidelines-and-quality-care/clinical-practice-guidelines/venous-thromboembolism-guidelines/ash-guidelines-on-use-of-anticoagulation-in-patients-with-covid-19 |
| Pragmatic Recommendations for Safety while Caring for Hospitalized Patients with Coronavirus Disease 2019 (COVID-19) in Low- and Middle-Income Countries | Does not include public health recommendations | 2020 | American Society of Tropical Medicine and Hygiene | https://www.ajtmh.org/view/journals/tpmd/104/3_Suppl/article-p12.xml |
| Updated guidance on the management of COVID-19: from an American Thoracic Society/European Respiratory Society coordinated International Task Force | Does not include public health recommendations | 2020 | American Thoracic Society, European Respiratory Society | https://bit.ly/32B96uI |
| Thromboembolism and anticoagulant therapy during the COVID‐19 pandemic: interim clinical guidance from the anticoagulation forum | Does not include public health recommendations | 2020 | Anticoagulation Forum | https://pubmed.ncbi.nlm.nih.gov/32440883/ |
| Consensus on treatment of multisystemic inflammatory syndrome associated with COVID-19 | Does not include public health recommendations | 2021 | Argentina Society of Pediatrics Subcommittees, Committees and Working Groups | https://www.sap.org.ar/docs/publicaciones/archivosarg/2021/v119n4a39s.pdf |
| Updated APLAR consensus statements on care for patients with rheumatic diseases during the COVID-19 pandemic | Does not include public health recommendations | 2021 | Asia Pacific League of Associations for Rheumatology (APLAR) | https://onlinelibrary.wiley.com/doi/10.1111/1756-185X.14124 |
| Spanish consensus document on diagnosis, stabilisation and treatment of pediatric multisystem inflammatory syndrome related to SARS-CoV-2 (SIM-PedS) | Does not include public health recommendations | 2021 | Asociación Española de Pediatría | https://www.sciencedirect.com/science/article/pii/S234128792100003X |
| COVID-19 convalescent plasma: Interim recommendations from the AABB | Does not include public health recommendations | 2021 | Association for the Advancement of Blood & Biotherapies (AABB) | https://onlinelibrary.wiley.com/doi/10.1111/trf.16328 |
| Clinical Practice Guidelines From the Association for the Advancement of Blood and Biotherapies (AABB): COVID-19 Convalescent Plasma | Does not include public health recommendations | 2022 | Association for the Advancement of Blood and Biotherapies (AABB) | https://www.ncbi.nlm.nih.gov/pmc/articles/PMC9450870/pdf/aim-olf-M221079.pdf |
| Timing of elective surgery and risk assessment after SARS-CoV-2 infection: an update | Does not include public health recommendations | 2022 | Association of Anaesthetists, Centre for Perioperative Care, Federation of Surgical Specialty Associations, Royal College of Anaesthetists, Royal College of Surgeons of England | <https://pubmed.ncbi.nlm.nih.gov/35194788/> |
| SARS-CoV-2 infection, COVID-19 and timing of elective surgery: A multidisciplinary consensus statement on behalf of the Association of Anaesthetists, the Centre for Peri-operative Care, the Federation of Surgical Specialty Associations, the Royal College of Anaesthetists and the Royal College of Surgeons of England. | Does not include public health recommendations | 2021 | Association Of Anesthesia | https://associationofanaesthetists-publications.onlinelibrary.wiley.com/doi/epdf/10.1111/anae.15464 |
| Managing haematology and oncology patients during the COVID‐19 pandemic: interim consensus guidance | Does not include public health recommendations | 2020 | Australasian Leukaemia and Lymphoma Group, National Centre for Infections in Cancer | https://www.ncbi.nlm.nih.gov/pmc/articles/PMC7273031/ |
| SARS-CoV-2 (COVID-19) vaccination in dermatology patients on immunomodulatory and biologic agents: Recommendations from the Australasian Medical Dermatology Group. | Does not include public health recommendations | 2020 | Australasian Medical Dermatology Group | https://www.ncbi.nlm.nih.gov/pmc/articles/PMC8250550/pdf/AJD-62-151.pdf |
| ANZICS guiding principles for complex decision making during the COVID-19 pandemic | Does not include public health recommendations | 2020 | Australian and New Zealand Intensive Care Society (ANZICS) | https://www.anzics.com.au/wp-content/uploads/2020/04/ANZI_3367_Guiding-Principles.pdf |
| Australian guidelines for the clinical care of people with COVID-19 "Update coming soon" | Does not include public health recommendations | 2022 | Australian National COVID-19 Clinical Evidence Taskforce | https://app.magicapp.org/#/guideline/L4Q5An |
| Brazilian guidelines for the treatment of outpatients with suspected or confirmed COVID-19. A joint guideline of the Brazilian Association of Emergency Medicine (ABRAMEDE), Brazilian Medical Association (AMB), Brazilian Society of Angiology and Vascular Surgery (SBACV), Brazilian Society of Geriatrics and Gerontology (SBGG), Brazilian Society of Infectious Diseases (SBI), Brazilian Society of Family and Community Medicine (SBFMC), and Brazilian Thoracic Society (SBPT) | Does not include public health recommendations | 2022 | Brazilian Association of Emergency Medicine (ABRAMEDE), Brazilian Medical Association (AMB), Brazilian Society of Angiology and Vascular Surgery (SBACV), Brazilian Society of Geriatrics and Gerontology (SBGG), Brazilian Society of Infectious Diseases (SBI), Brazilian Society of Family and Community Medicine (SBFMC), Brazilian Thoracic Society (SBPT) | https://www.sciencedirect.com/science/article/pii/S1413867022000356 |
| Brazilian Guidelines for the pharmacological treatment of patients hospitalized with COVID-19 | Does not include public health recommendations | 2022 | Brazilian Association of Emergency Medicine, Brazilian Association of Intensive Care Medicine, Brazilian Medical Association, Brazilian Society of Angiology and Vascular Surgery, Brazilian Society of Infectious Diseases, Brazilian Society of Pulmonology and Phthisiology, Brazilian Society of Rheumatology | https://www.scielo.br/j/rbti/a/hbNqqXwv3L4csjb5HjmyqFv/abstract/?lang=en |
| Guidelines for the pharmacological treatment of COVID-19 | Does not include public health recommendations | 2020 | Brazilian Association of Intensive Care Medicine (AMIB), Brazilian Society of Infectious Diseases (SBI), Brazilian Society of Pulmonology and Tisiology (SBPT) | https://www.scielo.br/scielo.php?script=sci_arttext&pid=S0103-507X2020000200166&lng=en&nrm=iso&tlng=en |
| Pan-American Guidelines for the Treatment of SARS-CoV-2/COVID-19: a joint evidence-based guideline of the Brazilian Society of Infectious Diseases (SBI) and the Pan-American Association of Infectious Diseases | Does not include public health recommendations | 2023 | Brazilian Society of Infectious Diseases (SBI), the Pan-American Association of Infectious Diseases (API) | https://docs.bvsalud.org/biblioref/2023/02/1416163/33abd3ae-f8fc-4475-b7df-759cbe801632.pdf |
| End-of-life care during the COVID-19 pandemic-What makes the difference? | Does not include public health recommendations | 2021 | British Association of Critical Care Nurses | https://onlinelibrary.wiley.com/doi/epdf/10.1111/nicc.12593 |
| Approach to assessment and management of long-term COVID-19 symptoms in primary care | Regional guideline | 2021 | British Columbia Ministry of Health | http://www.bccdc.ca/Health-Professionals-Site/Documents/Long-term_COVID-19_symptoms_primary_care.pdf |
| Critical Care Specialist Group Covid-19 Best Practice Guidance: Feeding Patients on Critical Care Units in the Prone Position (awake and sedated). Second edition | Does not include public health recommendations | 2020 | British Dietetic Association | https://www.bda.uk.com/uploads/assets/3f487dea-81e4-4277-bf1def44abc075bd/e319c889-23a3-4c7c-ab49e5efc9d82f91/201209-CCSG-BP-Guidance-for-Prone-Enteral-Feeding-Formatted-v2.pdf |
| British HIV Association Guidelines on Immunisation for Adults with HIV: SARS-CoV-2 (COVID-19) | Does not include public health recommendations | 2021 | British HIV Association (BHIVA) | https://www.bhiva.org/COVID-19-immunisation-guidelines-consultation |
| British Thoracic Society Guidance on Respiratory Follow Up of Patients with a Clinico-Radiological Diagnosis of COVID-19 Pneumonia | Does not include public health recommendations | 2020 | British Thoracic Society | https://www.brit-thoracic.org.uk/document-library/quality-improvement/covid-19/resp-follow-up-guidance-post-covid-pneumonia/ |
| Guidance for inpatient infection control management of patients receiving Acute Non Invasive Ventilation and Long-Term Ventilation during and beyond COVID-19 | Does not include public health recommendations | 2020 | British Thoracic Society | https://www.brit-thoracic.org.uk/document-library/quality-improvement/covid-19/guidance-for-inpatient-infection-control-management-receiving-acute-niv-and-ltb/ |
| BTS/ICS guidance: respiratory care in patients with acute hypoxaemic respiratory failure associated with COVID-19 | Does not include public health recommendations | 2021 | British Thoracic Society, Intensive Care Society | https://www.brit-thoracic.org.uk/document-library/quality-improvement/covid-19/btsics-guidance-respiratory-care-in-patients-with-acute-hypoxaemic-respiratory-failure-associated-with-covid-19/ |
| Canadian Association of Paediatric Nephrologists COVID-19 Rapid Response: Home and In-Center Dialysis Guidance. | Does not include public health recommendations | 2021 | Canadian Association of Paediatric Nephrologists | https://journals.sagepub.com/doi/10.1177/20543581211053458 |
| Clinical Practice Guideline for Solid Organ Donation and Transplantation During the COVID-19 Pandemic | Does not include public health recommendations | 2021 | Canadian Blood Services, Canadian Donation and Transplantation Research Program, Canadian Society of Transplantation, Peter Morris Centre for Evidence in Transplantation | <https://journals.lww.com/transplantationdirect/Fulltext/2021/10000/Clinical_Practice_Guideline_for_Solid_Organ.10.aspx> |
| Long COVID-19: A Primer for Cardiovascular Health Professionals, on Behalf of the CCS Rapid Response Team. | Does not include public health recommendations | 2021 | Canadian Cardiovascular Society | https://www.onlinecjc.ca/article/S0828-282X(21)00287-7/fulltext#relatedArticles |
| CRA Recommendation on Covid-19 Vaccination in Persons with Autoimmune Rheumatic Disease | Does not include public health recommendations | 2021 | Canadian Rheumatology Association (CRA) | https://rheum.ca/wp-content/uploads/2021/11/V3_Nov_23_2021_EN.pdf |
| Canadian Rheumatology Association Position Statement on COVID-19 Vaccination: Version 6.0 | Does not include public health recommendations | 2022 | Canadian Rheumatology Association (CRA) | https://rheum.ca/wp-content/uploads/2022/03/FINAL-Updated-PS-on-COVID-19-Vacc-March-25_2022_for-publication.pdf |
| Canadian Association of Paediatric Nephrologists COVID-19 Rapid Response: Guidelines for Management of Acute Kidney Injury in Children | Does not include public health recommendations | 2021 | Canadian Society of Nephrology | https://journals.sagepub.com/doi/pdf/10.1177/2054358121990135 |
| Revised recommendations from the CSO-HNS taskforce on performance of tracheotomy during the COVID-19 pandemic - what a difference a year makes | Does not include public health recommendations | 2021 | Canadian Society of Otolaryngology - Head & Neck Surgery (CSO-HNS) | https://journalotohns.biomedcentral.com/articles/10.1186/s40463-021-00531-z#citeas |
| Algorithms for testing COVID-19 focused on use of RT-PCR and high-affinity serological testing: a consensus statement from a panel of Latin American experts | Does not include public health recommendations | 2020 | Carlos Eduardo Ferreira, Pablo E. Bonvehic, Juan Carlos Gómez de la Torre, Klever Vinicio Sáenz-Flor, Antonio Condino-Neto | <https://www.ijidonline.com/article/S1201-9712(20)32484-X/fulltext> |
| Long Covid-19: Proposed Primary Care Clinical Guidelines for Diagnosis and Disease Management | Does not include public health recommendations | 2021 | Catalan Society of Family and Community Medicine (CAMFiC) | https://www.mdpi.com/1660-4601/18/8/4350 |
| Guidance for Reporting SARS-CoV-2 Sequencing Results | Does not include public health recommendations | 2022 | Centers for Disease Control and Prevention (CDC) | https://www.cdc.gov/coronavirus/2019-ncov/lab/resources/reporting-sequencing-guidance.html#print |
| Guidance for SARS-CoV-2 Rapid Testing Performed in Point-of-Care Settings | Does not include public health recommendations | 2022 | Centers for Disease Control and Prevention (CDC) | https://www.cdc.gov/coronavirus/2019-ncov/lab/point-of-care-testing.html |
| Interim Guidance for Managing Healthcare Personnel with SARS-CoV-2 Infection or Exposure to SARS-CoV-2 | Does not include public health recommendations | 2023 | Centers for Disease Control and Prevention (CDC) | https://www.cdc.gov/coronavirus/2019-ncov/hcp/guidance-risk-assesment-hcp.html |
| Interim guidance for SARS-CoV-2 testing in non-healthcare workplace | Does not include public health recommendations | 2021 | Centers for Disease Control and Prevention (CDC) | https://www.cdc.gov/coronavirus/2019-ncov/community/organizations/testing-non-healthcare-workplaces.html |
| Interim Guidance for Use of Pooling Procedures in SARS- CoV-2 Diagnostic and Screening Testing | Does not include public health recommendations | 2021 | Centers for Disease Control and Prevention (CDC) | https://www.cdc.gov/coronavirus/2019-ncov/lab/pooling-procedures.html |
| Interim Guidelines for Collecting and Handling of Clinical Specimens for COVID-19 Testing | Does not include public health recommendations | 2022 | Centers for Disease Control and Prevention (CDC) | https://www.cdc.gov/coronavirus/2019-ncov/lab/guidelines-clinical-specimens.html |
| Nucleic Acid Amplification Tests (NAATs) | Does not include public health recommendations | 2021 | Centers for Disease Control and Prevention (CDC) | https://www.cdc.gov/coronavirus/2019-ncov/lab/naats.html?s_cid=qr2021 |
| Strategies to Mitigate Healthcare Personnel Staffing Shortages | Does not include public health recommendations | 2022 | Centers for Disease Control and Prevention (CDC) | https://www.cdc.gov/coronavirus/2019-ncov/hcp/mitigating-staff-shortages.html |
| Testing Strategies for SARS-CoV-2 | Does not include public health recommendations | 2022 | Centers for Disease Control and Prevention (CDC) | https://www.cdc.gov/coronavirus/2019-ncov/hcp/testing-overview.html?CDC_AA_refVal=https%3A%2F%2Fwww.cdc.gov%2Fcoronavirus%2F2019-ncov%2Flab%2Fresources%2Fsars-cov2-testing-strategies.html |
| Considerations for Inpatient Obstetric Healthcare Settings | Does not include public health recommendations | 2021 | Centers for Disease Control and Prevention (CDC) | https://www.cdc.gov/coronavirus/2019-ncov/hcp/inpatient-obstetric-healthcare-guidance.html |
| Guidance for General Laboratory Safety Practices during the COVID-19 Pandemic | Does not include public health recommendations | 2022 | Centers for Disease Control and Prevention (CDC) | https://www.cdc.gov/coronavirus/2019-ncov/lab/lab-safety-practices.html |
| Information for Pediatric Healthcare Providers | Does not include public health recommendations | 2023 | Centers for Disease Control and Prevention (CDC) | https://www.cdc.gov/coronavirus/2019-ncov/hcp/pediatric-hcp.html |
| Interim Infection Prevention and Control Recommendations for Healthcare Personnel During the Coronavirus Disease 2019 (COVID-19) Pandemic | Does not include public health recommendations | 2023 | Centers for Disease Control and Prevention (CDC) | https://www.cdc.gov/coronavirus/2019-ncov/hcp/infection-control-recommendations.html |
| Interim Laboratory Biosafety Guidelines for Handling and processing Specimens Associated with CoronavirusDisease 2019 (COVID-19) | Does not include public health recommendations | 2021 | Centers for Disease Control and Prevention (CDC) | https://www.cdc.gov/coronavirus/2019-ncov/lab/lab-biosafety-guidelines.html |
| Operational Considerations for Infection Prevention and Control in Outpatient Facilities: non-U.S. Healthcare Settings | Does not include public health recommendations | 2021 | Centers for Disease Control and Prevention (CDC) | https://www.cdc.gov/coronavirus/2019-ncov/hcp/non-us-settings/outpatient.html |
| Operational Considerations for Routine Immunization Services during the COVID-19 pandemic in non-US Settings Focusing on Low- and Middle-Income Countries | Does not include public health recommendations | 2022 | Centers for Disease Control and Prevention (CDC) | https://www.cdc.gov/coronavirus/2019-ncov/global-covid-19/maintaining-immunization-services.html |
| Operational Considerations for the Identification of Healthcare Workers and Inpatients with Suspected COVID-19 in non-U.S. Healthcare Settings | Does not include public health recommendations | 2022 | Centers for Disease Control and Prevention (CDC) | https://www.cdc.gov/coronavirus/2019-ncov/hcp/non-us-settings/guidance-identify-hcw-patients.html |
| Standard Operating Procedure (SOP) for Triage of Suspected COVID-19 Patients in non-US Healthcare Settings | Does not include public health recommendations | 2021 | Centers for Disease Control and Prevention (CDC) | https://www.cdc.gov/coronavirus/2019-ncov/hcp/non-us-settings/sop-triage-prevent-transmission.html |
| ACIP Evidence to Recommendations for Use of Modern COVID-19 Vaccine in Children Ages 6 – 11 years and Adolescents Ages 12 – 17 years under an emergency Use Authorization | Not a guideline | 2022 | Centers for Disease Control and Prevention (CDC) | https://www.cdc.gov/vaccines/acip/recs/grade/covid-19-moderna-vaccine-6-17-years-etr.html |
| ACIP Update to the Evidence to Recommendations for a 2nd COVID-19 Booster Dose in Adults Ages 50 Years and Older and Immunocompromised Individuals | Not a guideline | 2022 | Centers for Disease Control and Prevention (CDC) | https://www.cdc.gov/vaccines/acip/recs/grade/covid-19-second-booster-dose-etr.html |
| ACIP Update to the Evidence to Recommendations for a Pfizer-BioNTech COVID-19 Booster in Children | Not a guideline | 2022 | Centers for Disease Control and Prevention (CDC) | https://www.cdc.gov/vaccines/acip/recs/grade/pfizer-biontech-covid19-booster-children-etr.html#print |
| Considerations for Non-emergency Vehicle Transportation for Tribal Communities During COVID-19 | Not a guideline | 2021 | Centers for Disease Control and Prevention (CDC) | https://www.cdc.gov/coronavirus/2019-ncov/community/tribal/non-ems-transportation.html#print |
| Use of COVID-19 Vaccines After Reports of Adverse Events Among Adult Recipients of Janssen (Johnson & Johnson) and mRNA COVID-19 Vaccines (Pfizer-BioNTech and Moderna): Update from the Advisory Committee on Immunization Practices — United States, July 2021 | Not a guideline | 2021 | Centers for Disease Control and Prevention (CDC) | https://www.cdc.gov/mmwr/volumes/70/wr/mm7032e4.htm?s_cid=mm7032e4_w |
| Use of mRNA COVID-19 Vaccine After Reports of Myocarditis Among Vaccine Recipients: Update from the Advisory Committee on Immunization Practices | Not a guideline | 2021 | Centers for Disease Control and Prevention (CDC) | https://www.cdc.gov/mmwr/volumes/70/wr/mm7027e2.htm |
| Guidance for Antigen Testing for SARS-CoV-2 for Healthcare Providers Testing Individuals in the Community | Does not include public health recommendations | 2023 | Centers for Disease Control and Prevention (CDC) | https://www.cdc.gov/coronavirus/2019-ncov/lab/resources/antigen-tests-guidelines.html |
| Interim Considerations: Preparing for the Potential Management of Anaphylaxis after COVID-19 Vaccination | Does not include public health recommendations | 2022 | Centers for Disease Control and Prevention (CDC) | https://www.cdc.gov/vaccines/covid-19/clinical-considerations/managing-anaphylaxis.html |
| Interim Guidelines for COVID-19 Antibody Testing | Does not include public health recommendations | 2022 | Centers for Disease Control and Prevention (CDC) | https://www.cdc.gov/coronavirus/2019-ncov/lab/resources/antibody-tests-guidelines.html |
| Investigating a COVID-19 case | Does not include public health recommendations | 2022 | Centers for Disease Control and Prevention (CDC) | https://www.cdc.gov/coronavirus/2019-ncov/php/contact-tracing/contact-tracing-plan/investigating-covid-19-case.html |
| Overview of Testing for SARS-CoV-2 (COVID-19) | Does not include public health recommendations | 2022 | Centers for Disease Control and Prevention (CDC) | https://www.cdc.gov/coronavirus/2019-ncov/hcp/testing-overview.html |
| Performing Broad-Based Testing for SARS-CoV-2 in Congregate Correctional, Detention, and Homeless Service Settings | Does not include public health recommendations | 2021 | Centers for Disease Control and Prevention (CDC) | https://www.cdc.gov/coronavirus/2019-ncov/hcp/broad-based-testing.html |
| Safe Watering Points During COVID-19 | Not a guideline | 2021 | Centers for Disease Control and Prevention (CDC) | https://www.cdc.gov/coronavirus/2019-ncov/community/safe-watering.html |
| Strategies for Optimizing the Supply of N95 Respirators | Does not include public health recommendations | 2021 | Centers for Disease Control and Prevention (CDC) | https://www.cdc.gov/coronavirus/2019-ncov/hcp/respirators-strategy/index.html#previous |
| Post-COVID Conditions: Information for Healthcare Providers | Does not include public health recommendations | 2022 | Centers for Disease Control and Prevention (CDC) | https://www.cdc.gov/coronavirus/2019-ncov/hcp/clinical-care/post-covid-conditions.html |
| Providing Care and Treatment for People Living with HIV in Low-Resource Non-US Settings During COVID-19 Pandemic | Does not include public health recommendations | 2021 | Centers for Disease Control and Prevention (CDC) | https://www.cdc.gov/coronavirus/2019-ncov/global-covid-19/maintaining-essential-HIV-services.html |
| COVID-19 Overview and Infection Prevention and Control Priorities in non-US Healthcare Settings | Not a guideline | 2021 | Centers for Disease Control and Prevention (CDC) | https://www.cdc.gov/coronavirus/2019-ncov/hcp/non-us-settings/overview/index.html |
| Ending Isolation and Precautions for People with COVID-19 | Does not include public health recommendations | 2022 | Centers for Disease Control and Prevention (CDC) | https://www.cdc.gov/coronavirus/2019-ncov/hcp/duration-isolation.html |
| Travel | Not a guideline | 2022 | Centers for Disease Control and Prevention (CDC) | https://www.cdc.gov/coronavirus/2019-ncov/travelers/index.html |
| Additional Information for Community Congregate Living Settings (e.g., Group Homes, Assisted Living) | Does not include public health recommendations | 2023 | Centre for Disease Control and Prevention (CDC) | https://www.cdc.gov/coronavirus/2019-ncov/community/community-congregate-living-settings.html |
| International Travel to and from the United States | Not a guideline | 2022 | Centres for Disease Control and Prevention (CDC) | https://www.cdc.gov/coronavirus/2019-ncov/travelers/international-travel-during-covid19.html |
| Working with longCOVID: Research evidence to inform support | Does not include public health recommendations | 2022 | Chartered Institute of Personnel and Development (CIPD) | https://www.cipd.co.uk/Images/long-covid-report-feb-22_tcm18-106089.pdf |
| Guidance of hemodialysis management and prevention during 2019 novel coronavirus Omicron variant | Does not include public health recommendations | 2022 | China National Knowledge Infrastructure (CNKI) | https://oversea.cnki.net/KCMS/detail/detail.aspx?dbcode=CAPJ&dbname=CAPJLAST&filename=SHYX20220423000&uniplatform=OVERSEAS_EN&v=1CWYO_FaZ1f642tIOQ2_rWfCx4F8-NhWywloNt_kyehIZ6Suc11Ow-7qXBFUn9uy |
| Rehabilitation guidance for integrated traditional Chinese and Western medicine in patients discharged from COVID-19 | Does not include public health recommendations | 2020 | China-based, name not specified | https://global.cnki.net/KCMS/detail/detail.aspx?dbcode=CJFD&dbname=CJFDLAST2020&filename=FYXB202004001&uniplatform=OVERSEA&v=aOrLFH6UAqtEXOAryvWdKvqzqAsQQQEypUaxrr1F3nvSAaKxqqJ8R_3ZGGgBj2mg |
| Recommendation of respiratory rehabilitation for PICS in critically ill patients with COVID-19 | Does not include public health recommendations | 2020 | Chinese Association of Chest Physician, Respiratory Rehabilitation Committee of Chinese Association of Rehabilitation Medicine | https://pubmed.ncbi.nlm.nih.gov/32894909/ |
| Recommendations for respiratory rehabilitation in adults with COVID-19 | Does not include public health recommendations | 2020 | Chinese Association of Rehabilitation Medicine, Respiratory Rehabilitation Committee of Chinese Association of Rehabilitation Medicine, Cardiopulmonary Rehabilitation Group of Chinese Society of Physical Medicine and Rehabilitation | https://www.ncbi.nlm.nih.gov/pmc/articles/PMC7470013/ |
| Pulmonary rehabilitation guidelines in the principle of 4S for patients infected with 2019 novel coronavirus (2019⁃nCoV) | Does not include public health recommendations | 2020 | Chinese Medical Association (CMA) | http://www.yiigle.com/LinkIn.do?linkin_type=pubmed&DOI=10.3760%2Fcma.j.issn.1001-0939.2020.03.007 |
| Holistic care for patients with severe coronavirus disease 2019: An expert consensus | Does not include public health recommendations | 2020 | Chinese Nursing Association | 10.3761/j.issn.0254-1769.2020.03.003 |
| Expert Recommendation on Severe Acute Respiratory Syndrome Coronavirus 2 Vaccination in Patients with Chronic Liver Diseases, TuberculosisExpert Recommendation on Severe Acute Respiratory Syndrome Coronavirus 2 Vaccination in Patients with Chronic Liver Diseases, Tuberculosis or Rheumatoid Diseases or Rheumatoid Diseases | Does not include public health recommendations | 2021 | Chinese Society of Infectious Diseases, Chinese Medical Association, Chinese Society of Rheumatology, Chinese Medical Association | https://mednexus.org/doi/epdf/10.1097/ID9.0000000000000021 |
| Humanitarian Surgical Missions in Times of COVID-19: Recommendations to Safely Return to a Sub-Saharan Africa Low-Resource Setting | Does not include public health recommendations | 2021 | Cirugía Solidaria | https://link.springer.com/article/10.1007/s00268-021-06001-x |
| Management of COVID‐19‐associated coagulopathy in persons with haemophilia | Does not include public health recommendations | 2020 | Coagulation Products Safety, Supply and Access (CPSSA) Committee of the World Federation of Hemophilia | https://onlinelibrary.wiley.com/doi/full/10.1111/hae.14191 |
| Key summary of German national treatment guidance for hospitalized COVID‑19 patients Key pharmacologic recommendations from a national German living guideline using an Evidence to Decision Framework | Does not include public health recommendations | 2021 | COVID-19 Evidence Ecosystem Project (CEOsys) | https://link.springer.com/article/10.1007/s15010-021-01645-2 |
| Pragmatic Recommendations for the Management of Acute Respiratory Failure and Mechanical Ventilation in Patients with COVID-19 in Low- and Middle-Income Countries | Does not include public health recommendations | 2021 | COVID-LMIC Task Force, Mahidol-Oxford Research Unit (MORU) | https://www.ajtmh.org/view/journals/tpmd/104/3_Suppl/article-p60.xml |
| Pragmatic Recommendations for Identification and Triage of Patients with COVID-19 Disease in Low- and Middle-Income Countries. | Does not include public health recommendations | 2021 | COVID–LMIC Task Force and the Mahidol–Oxford Research Unit(MORU) | https://www.ncbi.nlm.nih.gov/pmc/articles/PMC7957239/pdf/tpmd201064.pdf |
| Pragmatic Recommendations for the Prevention and Treatment of Acute Kidney Injury in Patients with COVID-19 in Low- and Middle-Income Countries. | Does not include public health recommendations | 2021 | COVID–LMIC Task Force, Mahidol–Oxford Research Unit(MORU) | https://www.ncbi.nlm.nih.gov/pmc/articles/PMC7957240/pdf/tpmd201242.pdf |
| COVID-19 vaccine guidance for patients with cancer participating in oncology clinical trials | Does not include public health recommendations | 2021 | COVID19 and Cancer Clinical Trials Working Group | https://www.nature.com/articles/s41571-021-00487-z |
| PICO Questions and DELPHI Methodology for the Management of Venous Thromboembolism Associated with COVID-19 | Does not include public health recommendations | 2021 | COVILAX Project | https://www.mdpi.com/1999-4915/13/11/2128 |
| Prevence a léčba COVID-19, první aktualizace [Prevention and treatment of COVID-19, the first update] | Does not include public health recommendations | 2023 | Czech Health Research Council (AZV ČR) | <https://kdp.uzis.cz/index.php?pg=kdp&id=52> |
| The Stanford Hall consensus statement for post-COVID- 19 rehabilitation | Does not include public health recommendations | 2021 | Defence Medical Rehabilitation Centre | https://bjsm.bmj.com/content/bjsports/54/16/949.full.pdf |
| Position Statement on COVID-19 and Pregnancy in Women with Heart Disease Department of Women Cardiology of the Brazilian Society of Cardiology - 2020. | Does not include public health recommendations | 2020 | Department of Women Cardiology of the Brazilian Society of Cardiology. | https://www.scielo.br/j/abc/a/8SVNP3tDnZSPh9pkbwbNrFw/?lang=en |
| Return to Activity After SARS-CoV-2 Infection: Cardiac Clearance for Children and Adolescents | Does not include public health recommendations | 2021 | Devyani Chowdhury, Michael A. Fremed, Peter Dean, Julie S. Glickstein, Jeff Robinson, Neil Rellosa, Deepika Thacker, David Soma, Susannah M. Briskin, Chad Asplund, Jonathan Johnson, Christopher Snyder | <https://journals.sagepub.com/doi/10.1177/19417381211039746> |
| Recommendation for standardized medical care for children and adolescents with long COVID [Einheitliche Basisversorgung von Kindern und Jugendlichen mit Long COVID] | Does not include public health recommendations | 2022 | DGKJ convention societies | https://pubmed.ncbi.nlm.nih.gov/35637934/ |
| How to Best Protect People With Diabetes From the Impact of SARS-CoV-2: Report of the International COVID-19 and Diabetes Summit | Does not include public health recommendations | 2021 | Diabetes Technology Society | https://journals.sagepub.com/doi/full/10.1177/1932296820978399?rfr_dat=cr_pub++0pubmed&url_ver=Z39.88-2003&rfr_id=ori%3Arid%3Acrossref.org |
| NHG guidance on COVID-19 | Does not include public health recommendations | 2022 | Dutch General Practitioners Association | https://richtlijnen.nhg.org/standaarden/covid-19 |
| Recommendations for antibacterial therapy in adults with COVID-19 e an evidence based guideline | Does not include public health recommendations | 2020 | Dutch Working Party on Antibiotic Policy | https://www.sciencedirect.com/science/article/pii/S1198743X20305942 |
| Guía de práctica clínica basada en la evidencia para el abordaje del paciente adulto crítico con COVID-19: versión completa/ Evidence-based clinical practice guideline for the management of critically ill adult patients with COVID-19: full version | Does not include public health recommendations | 2021 | El Gobierno de El Salvador - MINISTERIO DE SALUD | https://docs.bvsalud.org/biblioref/2022/04/1363436/gpc-vr_covid_tc_033022_1333.pdf |
| Expert consensus on large-scale multi-scene mobile laboratory emergency testing for novel coronavirus nucleic acids | Does not include public health recommendations | 2021 | Emergency Nucleic Acid Testing Expert Group of Guangzhou Laboratory | https://rs.yiigle.com/CN112137202140/1335665.htm |
| Diagnosis and Management Considerations in Steroid-Related Hyperglycemia in COVID-19: A Position Statement from the Endocrine Society of India. | Does not include public health recommendations | 2021 | Endocrine Society of India | https://www.ncbi.nlm.nih.gov/pmc/articles/PMC8323636/ |
| EAACI statement on the diagnosis, management and prevention of severe allergic reactions to COVID‐19 vaccines | Does not include public health recommendations | 2021 | European Academy of Allergy and Clinical Immunology (EAACI) | https://onlinelibrary.wiley.com/doi/abs/10.1111/all.14739 |
| Thoracic Anesthesia of Patients With Suspected or Confirmed 2019 Novel Coronavirus Infection: Preliminary Recommendations for Airway Management by the European Association of Cardiothoracic Anaesthesiology Thoracic Subspecialty Committee | Does not include public health recommendations | 2020 | European Association of Cardiothoracic Anaesthesiology | https://www.ncbi.nlm.nih.gov/pmc/articles/PMC7151284/pdf/main.pdf |
| Considerations on the use of self-tests for COVID-19 in the EU/EEA | Not a guideline | 2021 | European Center for Disease Prevention and Control (ECDC) | https://www.ecdc.europa.eu/en/publications-data/considerations-use-self-tests-covid-19-eueea |
| Public health considerations and evidence to support decisions on the implementation of a second mRNA COVID-19 vaccine booster dose | Not a guideline | 2022 | European Center for Disease Prevention and Control (ECDC) | https://www.ecdc.europa.eu/en/publications-data/public-health-considerations-and-evidence-support-decisions-implementation-second |
| Considerations for the use of antibody tests for SARS-COV-2– first update | Does not include public health recommendations | 2022 | European Centre for Disease Prevention and Control (ECDC) | https://www.ecdc.europa.eu/sites/default/files/documents/Considerations-for-the-use-of-antibody-tests-for-SARS-CoV2-first-update.pdf |
| Introducing a coherent European framework for tuning COVID-19 response measures | Does not include public health recommendations | 2021 | European Centre for Disease Prevention and Control (ECDC) | https://www.ecdc.europa.eu/sites/default/files/documents/Framework-for-tuning-COVID-19-response-measures.pdf |
| Operational considerations for influenza surveillance in the WHO European Region during COVID-19: interim guidance. October 2020 | Does not include public health recommendations | 2021 | European Centre for Disease Prevention and Control (ECDC) | https://apps.who.int/iris/handle/10665/336079 |
| Options for the use of rapid antigen tests for COVID-19 in the EU/EEA - first update | Does not include public health recommendations | 2021 | European Centre for Disease Prevention and Control (ECDC) | https://www.ecdc.europa.eu/en/publications-data/options-use-rapid-antigen-tests-covid-19-eueea-first-update |
| Sequencing of SARS-CoV-2: first update | Does not include public health recommendations | 2021 | European Centre for Disease Prevention and Control (ECDC) | https://www.ecdc.europa.eu/en/publications-data/sequencing-sars-cov-2#no-link |
| COVID 19 vaccine effectiveness in adolescents aged 12 17 years and interim public health considerations for administration of a booster dose | Not a guideline | 2022 | European Centre for Disease Prevention and Control (ECDC) | https://www.ecdc.europa.eu/en/publications-data/covid-19-vaccine-effectiveness-adolescents-and-interim-considerations-for-booster-dose |
| Data collection on COVID-19 outbreaks in closed settings: long-term care facilities, version 2.1 | Not a guideline | 2022 | European Centre for Disease Prevention and Control (ECDC) | https://www.ecdc.europa.eu/sites/default/files/documents/Data-collection-on-COVID-19-outbreaks-in-closed-settings-Version%202.1-18-Feb-2022.pdf |
| Coronavirus disease 2019 (COVID-19) and supply of substances of human origin in the EU/EEA - second update | Not a guideline | 2020 | European Centre for Disease Prevention and Control (ECDC) | https://www.ecdc.europa.eu/sites/default/files/documents/covid-19-supply-substances-human-origin-second-update.pdf |
| COVID-19 Rail Protocol: Recommendations for safe resumption of railway services in Europe | Does not include public health recommendations | 2021 | European Centre for Disease Prevention and Control (ECDC) | https://www.era.europa.eu/content/covid-19-rail-protocol_en |
| Risk related to the spread of new SARS-CoV-2 variants of concern in the EU/EEA – first update | Not a guideline | 2021 | European Centre for Disease Prevention and Control (ECDC) | https://www.ecdc.europa.eu/en/publications-data/covid-19-risk-assessment-spread-new-variants-concern-eueea-first-update |
| Rollout of COVID-19 vaccines in the EU/EEA: challenges and good practice | Not a guideline | 2021 | European Centre for Disease Prevention and Control (ECDC) | https://www.ecdc.europa.eu/en/publications-data/rollout-covid-19-vaccines-eueea-challenges-and-good-practice |
| Infection prevention and control and preparedness for COVID-19 in healthcare settings | Does not include public health recommendations | 2021 | European Centre for Disease Prevention and Control (ECDC) | https://www.ecdc.europa.eu/en/publications-data/infection-prevention-and-control-and-preparedness-covid-19-healthcare-settings |
| Rapid Risk Assessment: Assessing SARS-CoV-2 circulation, variants of concern, non-pharmaceutical interventions and vaccine rollout in the EU/EEA, 16th update | Not a guideline | 2021 | European Centre for Disease Prevention and Control (ECDC) | https://www.ecdc.europa.eu/sites/default/files/documents/covid-19-rapid-risk-assessment-16th-update-september-2021.pdf |
| Technical guidance for antigenic SARS-CoV-2 monitoring | Does not include public health recommendations | 2022 | European Centre for Disease Prevention and Control (ECDC), World Health Organization (WHO) | https://www.ecdc.europa.eu/en/publications-data/technical-guidance-antigenic-sars-cov-2-monitoring |
| Methods for the detection and characterization of SARS-CoV-2 variants – second update | Does not include public health recommendations | 2022 | European Centre for Disease Prevention and Control (ECDC), World Health Organization (WHO) | https://apps.who.int/iris/handle/10665/360875 |
| Management of post-acute COVID-19 patients in geriatric rehabilitation: EuGMS guidance | Does not include public health recommendations | 2021 | European Geriatric Medicine (EuGMS) | https://www.ncbi.nlm.nih.gov/pmc/articles/PMC8605452/ |
| 2021 update of the EULAR points to consider on the use of immunomodulatory therapies in COVID-19 | Does not include public health recommendations | 2021 | European League Against Rheumatism (EULAR) | https://ard.bmj.com/content/81/1/34 |
| Fabry disease and COVID-19: international expert recommendations for management based on real-world experience | Does not include public health recommendations | 2020 | European Renal Association (ERA-EDTA) | https://academic.oup.com/ckj/article/13/6/913/6054307 |
| Update March 2022: management of hospitalised adults with coronavirus disease-19 (COVID-19): a European Respiratory Society living guideline | Does not include public health recommendations | 2022 | European Respiratory Society | https://erj.ersjournals.com/content/early/2022/06/09/13993003.00803-2022 |
| Nutritional management of individuals with obesity and COVID-19: ESPEN expert statements and practical guidance | Does not include public health recommendations | 2021 | European Society for Clinical Nutrition and Metabolism ESPEN | https://www.clinicalnutritionjournal.com/action/showPdf?pii=S0261-5614%2821%2900248-X |
| Coronavirus disease 2019 in adults with congenital heart disease: a position paper from the ESC working group of adult congenital heart disease | Does not include public health recommendations | 2020 | European Society of Cardiology | https://pubmed.ncbi.nlm.nih.gov/33313664/ |
| ESCMID COVID-19 guidelines: diagnostic testing for SARS-CoV-2 | Does not include public health recommendations | 2022 | European Society of Clinical Microbiology and Infectious Diseases (ESCMID) | https://www.clinicalmicrobiologyandinfection.com/article/S1198-743X(22)00084-2/fulltext |
| ESCMID COVID-19 Living Guidelines: Drug Treatment and Clinical Management | Does not include public health recommendations | 2022 | European Society of Clinical Microbiology and Infectious Diseases (ESCMID) | https://www.clinicalmicrobiologyandinfection.com/article/S1198-743X(22)00429-3/fulltext#secsectitle0065 |
| ESCMID guidelines on testing for SARS-CoV-2 in asymptomatic individuals to prevent transmission in the healthcare setting | Does not include public health recommendations | 2022 | European Society of Clinical Microbiology and Infectious Diseases (ESCMID) | https://linkinghub.elsevier.com/retrieve/pii/S1198-743X(22)00030-1 |
| ESCMID rapid guidelines for assessment and management of long COVID. | Does not include public health recommendations | 2022 | European Society of Clinical Microbiology and Infectious Diseases (ESCMID) | <https://www.clinicalmicrobiologyandinfection.com/article/S1198-743X(22)00092-1/fulltext> |
| Practice recommendations for the management of children with suspected or proven COVID-19 infections from the Paediatric Mechanical Ventilation Consensus Conference (PEMVECC) and the section Respiratory Failure from the European Society for Paediatric and Neonatal Intensive Care (ESPNIC) | Does not include public health recommendations | 2020 | European Society of Paediatric and Neonatal Intensive Care (ESPNIC) | https://espnic-online.org/News/Latest-News/Practice-recommendations-for-managing-children-with-proven-or-suspected-COVID-19 |
| European consensus recommendations for neonatal and paediatric retrievals of positive or suspected COVID-19 patients. | Does not include public health recommendations | 2020 | European Society of Paediatric and Neonatal Intensive Care (ESPNIC), Transport section and the European Society for Paediatric Research (ESPR) | https://www.nature.com/articles/s41390-020-1050-z |
| European stroke organization interim expert opinion on cerebral venous thrombosis occurring after SARS-CoV-2 vaccination | Does not include public health recommendations | 2021 | European Stroke Organization | https://journals.sagepub.com/doi/full/10.1177/23969873211030842 |
| Extracorporeal Membrane Oxygenation for COVID-19: Updated 2021 Guidelines from the Extracorporeal Life Support Organization. | Does not include public health recommendations | 2021 | Extracorporeal Life Support Organization (ELSO) | https://journals.lww.com/asaiojournal/Fulltext/2021/05000/Extracorporeal_Membrane_Oxygenation_for_COVID_19_.3.aspx |
| Post Abortion Care and Management After Induced Abortion During the COVID-19 Pandemic: A Chinese Expert Consensus. | Does not include public health recommendations | 2021 | Family Planning Group of Minimally Invasive Gynecological branch of the Liaoning Medical Association | https://www.ncbi.nlm.nih.gov/pmc/articles/PMC7812565/ |
| Aerosolization of COVID-19 and Contamination Risks During Respiratory Treatments. | Does not include public health recommendations | 2020 | Federal Practitioner | https://www.ncbi.nlm.nih.gov/pmc/articles/PMC7173638/pdf/fp-37-04-160.pdf |
| Infection with SARS-CoV-2 in pregnancy. Update of Information and proposed care. CNGOF | Does not include public health recommendations | 2020 | French National College of French Obstetrician Gynaecologists | https://www.ncbi.nlm.nih.gov/pmc/articles/PMC7534662/ |
| COVID19 and acute lymphoblastic leukemias of children and adolescents: Updated recommendations (Version 2) of the Leukemia Committee of the French Society for the fight against Cancers and leukemias in children and adolescents (SFCE) | Does not include public health recommendations | 2021 | French Society for the fight against Cancers and leukemias in children and adolescents (SFCE) | https://www.sciencedirect.com/science/article/pii/S0007455121000825?via%3Dihub |
| Management of drug-drug interactions with nirmatrelvir/ritonavir in patients treated for COVID-19: Guidelines from the French Society of Pharmacology and Therapeutics (SFPT) | Does not include public health recommendations | 2022 | French Society of Pharmacology and Therapeutics | https://www.ncbi.nlm.nih.gov/pmc/articles/PMC9020499/ |
| French Vasculitis Study Group recommendations for the management of COVID-19 vaccination and prophylaxis in patients with systemic vasculitis | Does not include public health recommendations | 2021 | French Vasculitis Study Group | https://www.ncbi.nlm.nih.gov/pmc/articles/PMC8704893/ |
| SARS-CoV-2/ Covid-19- Information & Practice Aids for Family Physicians in Private Practice | Does not include public health recommendations | 2022 | German Society for General and Family Medicine | https://www.awmf.org/uploads/tx_szleitlinien/053-054l_S2e_SARS-CoV-2-Covid-19-Informationen-Praxishilfen-Hausaerztinnen-Hausaerzte_2022-02_2.pdf |
| 2021 update of the AGIHO guideline on evidence-based management of COVID-19 in patients with cancer regarding diagnostics, viral shedding, vaccination and therapy | Does not include public health recommendations | 2021 | German Society for Haematology and Medical Oncology (DGHO) | https://pubmed.ncbi.nlm.nih.gov/33676266/ |
| Neurological manifestations of post-COVID-19 syndrome S1-guideline of the German society of neurology | Does not include public health recommendations | 2022 | German society of neurology | https://neurolrespract.biomedcentral.com/articles/10.1186/s42466-022-00191-y |
| Neurological manifestations in COVID-19 patients. S2k-LL (DGN) | Does not include public health recommendations | 2021 | German Society of Neurology (DGN), German Society of NeuroIntensive and Emergency Medicine (DGNI), German Society of Otolaryngology, Head and Neck Surgery (DGHNO-KHC), German Society of Neurorehabilitation (DGNR) | https://dgn.org/wp-content/uploads/2020/08/030144_LL_Neurologische_Manifestationen_bei_COVID-19_V3.1.pdf |
| Guideline S1: Long COVID: Diagnostics and treatment strategies | Does not include public health recommendations | 2021 | German Society of Pneumology, AMWF | https://link.springer.com/article/10.1007/s00508-021-01974-0 |
| Consensus Clinical Guidance for Diagnosis and Management of Adult COVID-19 Encephalopathy Patients | Does not include public health recommendations | 2022 | Global COVID-19 Neuro Research Coalition | https://neuro.psychiatryonline.org/doi/10.1176/appi.neuropsych.22010002?url_ver=Z39.88-2003&rfr_id=ori:rid:crossref.org&rfr_dat=cr_pub%20%200pubmed |
| COVID-19 and Thrombotic or Thromboembolic Disease: Implications for Prevention, Antithrombotic Therapy, and Follow-Up | Does not include public health recommendations | 2020 | Global COVID-19 Thrombosis Collaborative Group | https://www.sciencedirect.com/science/article/pii/S0735109720350087 |
| Updated Guidance on Continuity of Schooling: Supporting Pupils with Special Educational Needs For mainstream primary and special schools | Does not include public health recommendations | 2021 | Government of Ireland | https://www.gov.ie/en/collection/965639-continuity-of-schooling/#pupils-students-with-special-educational-needs |
| Framework to maintain Physical Distancing in the Classroom in Post Primary Schools with a Full Return of All Students for the 2020/21 School Year | Not a guideline | 2020 | Government of Ireland | https://webcache.googleusercontent.com/search?q=cache:my5v9N200P0J:https://assets.gov.ie/83472/ca0e3029-2d43-4e77-8181-bc3dc89455d2.pdf+&cd=1&hl=pt-BR&ct=clnk&gl=br&client=safari |
| Manitoba COVID-19 Vaccine: Clinical Practice Guidelines | Regional guideline | 2022 | Government of Manitoba | https://www.gov.mb.ca/asset_library/en/covidvaccine/clinical_practice_guidelines.pdf |
| New-Brunswick COVID-19 Vaccine Clinic Guide for Immunizers and Providers | Regional guideline | 2022 | Government of New-Brunswick | https://www2.gnb.ca/content/dam/gnb/Departments/eco-bce/Promo/covid-19/Vaccine-Clinic-Guide.pdf |
| COVID-19 vaccination in patients suffering from respiratory diseases. Update of 25th June 2021 | Does not include public health recommendations | 2021 | Group for Research and Teaching in Pneumo-Infectiology (GREPI) | https://www.ncbi.nlm.nih.gov/pmc/articles/PMC8330974/ |
| SARS-CoV-2 safety: Guidelines for shielding frontline nurses | Does not include public health recommendations | 2021 | Guidelines for Shielding frontline nurses | https://journals.lww.com/nursing/Abstract/2021/03000/SARS_CoV_2_safety__Guidelines_for_shielding.11.aspx |
| Palestinian strategies, guidelines, and challenges in the treatment and management of coronavirus disease-2019 (COVID-19). | Does not include public health recommendations | 2020 | Hatem A. Hejaz | <https://www.thieme-connect.com/products/ejournals/abstract/10.4103/ajm.ajm_171_20> |
| COVID-19 position statement: Maternal critical care provision | Does not include public health recommendations | 2020 | Healthcare Improvement Scotland, SIGN | https://www.sign.ac.uk/media/1787/sg-maternal-critical-care-provision_v33.pdf |
| Managing hyperlipidaemia in patients with COVID-19 and during its pandemic: An expert panel position statement from HEART UK | Does not include public health recommendations | 2020 | Heart UK's Medical Scientific and Research Committee | https://www.ncbi.nlm.nih.gov/pmc/articles/PMC7490256/ |
| Prostate cancer management in the era of COVID-19: Recommendations from the Hong Kong Urological Association and Hong Kong Society of Uro-oncology | Does not include public health recommendations | 2021 | Hong Kong Urological Association, Hong Kong Society of Uro-oncology | https://onlinelibrary.wiley.com/doi/full/10.1111/ajco.13579 |
| IAP Guideline on Practicing Safely During COVID-19 Era: Clinics and Small Establishments | Does not include public health recommendations | 2021 | Indian Academy of Pediatrics (IAP) | http://www.indianpediatrics.net/apr2021/383.pdf |
| Clinical Guidance for management of adult COVID-19 patients | Does not include public health recommendations | 2022 | Indian Council of Medical Research- COVID-19 National Task Force, Government of India | https://www.icmr.gov.in/pdf/covid/techdoc/COVID_Clinical_Management_14012022.pdf |
| Infectious Diseases Society of America guidelines on infection prevention in patients with suspected or known COVID-19 | Does not include public health recommendations | 2021 | Infectious Disease Society of America (IDSA) | https://www.idsociety.org/practice-guideline/covid-19-guideline-infection-prevention/ |
| Infectious Diseases Society of America Guidelines on Infection Prevention for Healthcare Personnel Caring for Patients with Suspected or Known COVID-19 | Does not include public health recommendations | 2021 | Infectious Diseases Society of America (IDSA) | https://www.idsociety.org/practice-guideline/covid-19-guideline-infection-prevention/ |
| Infectious Diseases Society of America Guidelines on the Diagnosis of COVID-19: Serologic Testing | Does not include public health recommendations | 2020 | Infectious Diseases Society of America (IDSA) | https://www.idsociety.org/practice-guideline/covid-19-guideline-serology/ |
| Infectious Diseases Society of America Guidelines on the Treatment and Management of Patients with COVID-19 | Does not include public health recommendations | 2023 | Infectious Diseases Society of America (IDSA) | https://www.idsociety.org/practice-guideline/covid-19-guideline-treatment-and-management/# |
| The Infectious Diseases Society of America Guidelines on the Diagnosis of COVID-19: Antigen Testing | Does not include public health recommendations | 2023 | Infectious Diseases Society of America (IDSA) | https://academic.oup.com/cid/advance-article/doi/10.1093/cid/ciad032/7005394 |
| Covid-19 patient management and care pathway in Tunisia | Does not include public health recommendations | 2021 | Instance Nationale de l'Evaluation et de l'Accreditation en Santé (INEAS) | https://www.ineas.tn/sites/default/files/gpc_covid_19_version_11_mai_2021.pdf |
| Clinical practice guideline for the management of covid-19 (adults) version 3 | Does not include public health recommendations | 2021 | Institute for Health Technology Assessment and Research (EsSalud) | http://www.essalud.gob.pe/ietsi/pdfs/guias/GPC_COVID_19_Version_corta.pdf |
| Clinical practice guideline for the management of COVID-19 in pediatrics | Does not include public health recommendations | 2022 | Institute for Health Technology Evaluation and Research | https://ietsi.essalud.gob.pe/wp-content/uploads/2022/01/GPC-COVID-19-en-Pediatria_Version-corta.pdf |
| A Practical Guide for Anesthesia Providers on the Management of Coronavirus Disease 2019 Patients in the Acute Care Hospital | Does not include public health recommendations | 2021 | International Anesthesia Research Society (IARS) | https://journals.lww.com/anesthesia-analgesia/Fulltext/2021/03000/A_Practical_Guide_for_Anesthesia_Providers_on_the.2.aspx/Document.pdf |
| Cardio‐oncology care in the era of the coronavirus disease 2019 (COVID‐19) pandemic: An International Cardio‐Oncology Society (ICOS) statement | Does not include public health recommendations | 2020 | International Cardio‐Oncology Society (ICOS) | https://acsjournals.onlinelibrary.wiley.com/doi/10.3322/caac.21635 |
| IFCC interim guidelines on rapid point-of-care antigen testing for SARS-CoV-2 detection in asymptomatic and symptomatic individuals | Does not include public health recommendations | 2021 | International Federation of Clinical Chemistry and Laboratory Medicine (IFCC) | https://www.degruyter.com/document/doi/10.1515/cclm-2021-0455/html |
| Joint position statement on management of patient with osteoporosis during COVID-19 contingency from the AMMOM, CONAMEGER, FELAEN, FEMECOG, FEMECOT, and ICAAFYD | Does not include public health recommendations | 2021 | International Osteoporosis Foundation, National Osteoporosis Foundation | https://link.springer.com/article/10.1007/s11657-020-00869-3 |
| Good practice statements for antithrombotic therapy in the management of COVID- 19: Guidance from the SSC of the ISTH | Does not include public health recommendations | 2022 | International Society of Thrombosis and Haemostasis (ISTH) | https://onlinelibrary.wiley.com/doi/epdf/10.1111/jth.15809 |
| Consensus-based clinical recommendations and research priorities for anticoagulant thromboprophylaxis in children hospitalized for COVID-19-related illness. | Does not include public health recommendations | 2020 | International Society on Thrombosis and Haemostasis | https://onlinelibrary.wiley.com/doi/epdf/10.1111/jth.15073 |
| ISTH interim guidance on recognition and management of coagulopathy in COVID-19 | Does not include public health recommendations | 2020 | International Society on Thrombosis and Haemostasis (ISTH) | https://onlinelibrary.wiley.com/doi/10.1111/jth.14810 |
| Scientific and Standardization Committee communication: clinical guidance on the diagnosis, prevention, and treatment of venous thromboembolism in hospitalized patients with COVID-19 | Does not include public health recommendations | 2020 | International Society on Thrombosis and Haemostasis (ISTH) | https://onlinelibrary.wiley.com/doi/epdf/10.1111/jth.14929 |
| Interim Guidance on Long-COVID Management Principles | Does not include public health recommendations | 2021 | Italian National Institute of Health | https://www.iss.it/documents/20126/0/Rapporto+ISS+COVID-19+n.15_2021_EN.pdf/dd962ad9-fa53-73dd-7759-55cb5c167675?t=1627575304593 |
| Clinical Management of Adult Patients with COVID-19 Outside Intensive Care Units: Guidelines from the Italian Society of Anti-Infective Therapy (SITA) and the Italian Society of Pulmonology (SIP) | Does not include public health recommendations | 2021 | Italian Society of Anti-Infective Therapy (SITA), Italian Society of Pulmonology (SIP) | https://doi.org/10.1007/s40121-021-00487-7. |
| Resilience and response of the congenital cardiac network in Italy during the COVID-19 pandemic | Does not include public health recommendations | 2020 | Italian Society of Pediatric Cardiology and Congenital Heart Disease | https://journals.lww.com/jcardiovascularmedicine/Fulltext/2021/01000/Resilience_and_response_of_the_congenital_cardiac.2.aspx |
| Italian intersociety consensus on management of long covid in children | Does not include public health recommendations | 2022 | Italian Society of Pediatrics (SIP) | https://ijponline.biomedcentral.com/articles/10.1186/s13052-022-01233-6 |
| Covid-19 Infection in Cancer Patients: The Management in a Diagnostic Unit | Does not include public health recommendations | 2021 | Italian Society of Radiology and Interventional Radiology (SIRM) | https://www.ncbi.nlm.nih.gov/pmc/articles/PMC8042821/ |
| Immune-related (IR)-pneumonitis during the COVID-19 pandemic: multidisciplinary recommendations for diagnosis and management. | Does not include public health recommendations | 2020 | Jarushka Naidoo, Joshua E. Reuss, Karthik Suresh, David Feller-Kopman, Patrick M. Forde, Seema Mehta Steinke, Clare Rock, Douglas B. Johnson, Mizuki Nishino, Julie R. Brahmer | <https://jitc.bmj.com/content/8/1/e000984.long> |
| Joint Committee on Vaccination and Immunisation: advice on priority groups for COVID-19 vaccination, 30 December 2020 | Not a guideline | 2020 | Joint Committee on Vaccination and Immunisation (JCVI) | https://www.gov.uk/government/publications/priority-groups-for-coronavirus-covid-19-vaccination-advice-from-the-jcvi-30-december-2020/joint-committee-on-vaccination-and-immunisation-advice-on-priority-groups-for-covid-19-vaccination-30-december-2020 |
| How to manage inflammatory bowel disease during the COVID-19 pandemic: A guide for the practicing clinician | Does not include public health recommendations | 2021 | Júlio Maria Fonseca Chebli, Natália Sousa Freitas Queiroz, Adérson Omar Mourão Cintra Damião, Liliana Andrade Chebli, Márcia Henriques de Magalhães Costa, Rogério Serafim Parra | <https://www.ncbi.nlm.nih.gov/pmc/articles/PMC7985732/> |
| Return to School for Pediatric Solid Organ Transplant Recipients in the United States During the Coronavirus Disease 2019 Pandemic: Expert Opinion on Key Considerations and Best Practices | Does not include public health recommendations | 2020 | Kevin J. Downes, Lara A. Danziger-Isakov, Melissa K. Cousino, Michael Green, Marian G. Michaels, William J. Muller, Rachel C. Orscheln, Tanvi S. Sharma, Victoria A. Statler, Rachel L. Wattier, Monica I. Ardura | <https://www.ncbi.nlm.nih.gov/pmc/articles/PMC7454776/> |
| COVID-19 vaccines: Global challenges and prospects forum recommendations | Not a guideline | 2021 | King Abdullah International Medical Research Centre (KAIMRC) Annual Research Forum | https://www.ijidonline.com/article/S1201-9712(21)00179-X/fulltext |
| Korean College of Rheumatology Task Force for COVID-19 Vaccine Guidance for Patients with Autoimmune Inflammatory Rheumatic Diseases | Does not include public health recommendations | 2021 | Korean College of Rheumatology | https://jkms.org/DOIx.php?id=10.3346/jkms.2021.36.e95 |
| COVID-19 Vaccination for Endocrine Patients: A Position Statement from the Korean Endocrine Society | Not a guideline | 2021 | Korean Endocrine Society | https://www.ncbi.nlm.nih.gov/pmc/articles/PMC8419616/ |
| Guidelines for Mobile Laboratories for Molecular Diagnostic Testing of COVID-19 | Does not include public health recommendations | 2022 | Korean Society for Laboratory Medicine (KSLM), Korea Disease Control and Prevention Agency (KDCA) | https://www.annlabmed.org/journal/view.html?doi=10.3343/alm.2022.42.5.507 |
| Update of Guidelines for Laboratory Diagnosis of COVID-19 in Korea | Does not include public health recommendations | 2022 | Korean Society for Laboratory Medicine (KSLM), Korea Disease Control and Prevention Agency (KDCA) | https://www.annlabmed.org/journal/view.html?doi=10.3343/alm.2022.42.4.391 |
| Preliminary Guidelines for the Clinical Evaluation and Management of Long COVID | Does not include public health recommendations | 2022 | Korean Society of Infectious Diseases | https://www.ncbi.nlm.nih.gov/pmc/articles/PMC9533168/ |
| Pragmatic Recommendations for the Use of Diagnostic Testing and Prognostic Models in Hospitalized Patients with Severe COVID-19 in Low- and Middle-Income Countries | Does not include public health recommendations | 2021 | Mahidol-Oxford Research Unit (MORU) | https://www.ncbi.nlm.nih.gov/pmc/articles/PMC7957242/ |
| Prioritizing surgery during the COVID-19 pandemic: the Quebec guidelines. | Does not include public health recommendations | 2021 | Marie-Eve Bouthillier, Michel Lorange, Serge Legault, Lucie Wade, Joseph Dahine, Jean Latreille, Isabelle Germain, Roger Grégoire, Patrick Montpetit, Catherine Prady, Elise Thibault, Vincent Dumez, Lucie Opatrny | <https://www.canjsurg.ca/content/64/1/E103.long> |
| Clinical practice guideline: tocilizumab for patients with severe and critical COVID-19 | Does not include public health recommendations | 2021 | Martín A. Ragusa, Fernando Tortosa, Gabriela Carrasco, Guadalupe Montero, Pedro Haluska, Laura Lamfre, Ariel Izcovich | <https://pesquisa.bvsalud.org/portal/resource/pt/biblio-1365097> |
| Recommendations for the recognition, diagnosis, and management of long COVID: a Delphi study | Does not include public health recommendations | 2021 | Martine Nurek, Clare Rayner, Anette Freyer, Sharon Taylor, Linn Järte, Nathalie MacDermott, Brendan C. Delaney | <https://bjgp.org/content/71/712/e815#page> |
| Current Recommendations for the Management of Stroke Patients in the Middle East in the Era of COVID-19 Pandemic; Statement from the MENA SINO | Does not include public health recommendations | 2020 | Middle East and North Africa Stroke and Interventional Neurotherapies Organization (MENA-SINO) | https://www.strokejournal.org/article/S1052-3057(20)30599-1/fulltext |
| Chemoprophylaxis, diagnosis, treatments, and discharge management of COVID-19: An evidence-based clinical practice guideline (updated version) | Does not include public health recommendations | 2020 | Military Medical Research | https://mmrjournal.biomedcentral.com/articles/10.1186/s40779-020-00270-8 |
| Egyptian Consensus on the Role of Lung Ultrasonography During the Coronavirus Disease 2019 Pandemic | Does not include public health recommendations | 2022 | Ministry of Health and Population COVID-19 board, Egyptian Society of fever (ESF), UCHID-COVID-19 special interest group | https://pubmed.ncbi.nlm.nih.gov/36176457/ |
| Guía de práctica clínica basada en la evidencia para el abordaje del paciente con COVID-19 leve y moderada: versión completa/ Evidence-based clinical practice guideline for the approach to patients with mild and moderate COVID-19: full version | Does not include public health recommendations | 2022 | Ministry of Health of El Salvador | https://docs.bvsalud.org/biblioref/2022/04/1363436/gpc-vr_covid_tc_033022_1333.pdf |
| Endocrinology in the time of COVID-19-2021 updates: The management of diabetes insipidus and hyponatraemia. | Does not include public health recommendations | 2021 | Mirjam Christ-Crain, Ewout J. Hoorn, Mark Sherlock, Chris J. Thompson, John Wass | <https://academic.oup.com/ejendo/article/185/4/G35/6654315?searchresult=1> |
| 20 questions about children's new coronavirus vaccine vaccination | Not a guideline | 2021 | National Clinical Research Center for Respiratory Diseases, National Children's Medical Center, Respiratory Group, Pediatrics Branch of Chinese Medical Association, Pediatric Respiratory Working Committee of Respiratory Physician Branch of Chinese Medical Doctor Association, Pediatric Professional Committee of China Medical Education Association, Pediatrics Committee of China Association of Research Hospitals, Pediatric Professional Committee of China Association of Non-public Medical Institutions, Child Health and Drug Research Committee of China Association of Chinese Materia Medica, Children's Safe Medication Branch of China Medical News and Information Association, "June 1" Health Express Project Expert Committee, Global Pediatric Respiratory Alliance | https://rs.yiigle.com/CN101070202118/1337655.htm |
| Development of Evidence-Based COVID-19 Management Guidelines for Local Context: The Methodological Challenges | Does not include public health recommendations | 2022 | National Command Operation Center (NCOC) | <https://www.ncbi.nlm.nih.gov/pmc/articles/PMC9020141/> |
| Clinical care of children and adolescents with COVID-19: recommendations from the National COVID-19 Clinical Evidence Taskforce | Does not include public health recommendations | 2021 | National COVID-19 Clinical Evidence Taskforce | <https://www.ncbi.nlm.nih.gov/pmc/articles/PMC8661691/> |
| Australian guidelines for SARS-CoV-2 infection prevention and control of COVID-19 in healthcare workers | Does not include public health recommendations | 2021 | National COVID-19 clinical evidence taskforce | <https://www.health.qld.gov.au/__data/assets/pdf_file/0038/939656/qh-covid-19-Infection-control-guidelines.pdf> |
| Interim Adapted Clinical Guidelines for Post-COVID Conditions (Long-COVID) | Does not include public health recommendations | 2021 | National Health Care for the Homeless Council | <https://nhchc.org/wp-content/uploads/2021/10/Adapted-Clinical-Guidelines-Long-COVID-FINAL-10.14.pdf> |
| National commissioning guidance for post COVID services | Does not include public health recommendations | 2022 | National Health Service | <https://www.england.nhs.uk/wp-content/uploads/2022/07/C1670_National-commissioning-guidance-for-post-COVID-services_V3_July-2022-1.pdf> |
| Interim Clinical Commissioning Policy: IL-6 inhibitors (tocilizumab or sarilumab) for hospitalised patients with COVID-19 (adults) | Does not include public health recommendations | 2022 | National Health Service (NHS) | <http://www.england.nhs.uk/coronavirus/documents/interim-clinical-commissioning-policy-il-6-inhibitors-tocilizumab-or-sarilumab-for-hospitalised-patients-with-covid-19-adults-2/> |
| Managing the long-term effects of COVID - guidance for primary care | Does not include public health recommendations | 2021 | National Health Service (NHS) | <https://www.leedsccg.nhs.uk/content/uploads/2022/03/Managing-long-term-effects-of-Covid-19-guidance-for-primary-care-updated-March-2022-Copy-1.pdf> |
| Guidelines for managing people with lymphoedema remotely: a post-COVID-19 response document | Does not include public health recommendations | 2021 | National Health Service (NHS) | <https://www.magonlinelibrary.com/doi/epub/10.12968/bjon.2021.30.4.218> |
| COVID-19 rapid guideline: haematopoietic stem cell transplantation | Does not include public health recommendations | 2022 | National Institute for Health and Care Excellence | <https://www.nice.org.uk/guidance/ng164/evidence> |
| COVID-19 rapid guideline: managing COVID-19 | Does not include public health recommendations | 2023 | National Institute for Health and Care Excellence | <https://www.nice.org.uk/guidance/ng191> |
| COVID-19 rapid guideline: vaccine-induced immune thrombocytopenia and thrombosis (VITT) | Does not include public health recommendations | 2022 | National Institute for Health and Care Excellence (NICE) | <https://app.magicapp.org/#/guideline/nYP2ZL/section/Lrbr8L> |
| COVID-19 rapid guideline: delivery of systemic anticancer treatments | Does not include public health recommendations | 2022 | National Institute for Health and Care Excellence (NICE) | <https://www.nice.org.uk/guidance/ng161> |
| COVID-19 Rapid Guideline: Vitamin D | Does not include public health recommendations | 2022 | National Institute for Health and Care Excellence (NICE) | <https://www.nice.org.uk/guidance/ng187> |
| COVID-19 rapid guideline: Managing the long-term effects of COVID-19 | Does not include public health recommendations | 2022 | National Institute for Health and Care Excellence (NICE), Scottish Intercollegiate Guidelines Network (SIGN), Royal College of General Practitioners (RCGP) | <https://www.nice.org.uk/guidance/ng188> |
| Coronavirus Disease 2019 (COVID-19) Treatment Guidelines | Does not include public health recommendations | 2023 | National Institutes of Health | <https://www.covid19treatmentguidelines.nih.gov/about-the-guidelines/whats-new/> |
| National Psoriasis Foundation COVID-19 Task Force Guidance for Management of Psoriatic Disease During the Pandemic: Version 2 - Advances in Psoriatic Disease Management, COVID-19 Vaccines, and COVID-19 Treatments | Does not include public health recommendations | 2021 | National Psoriasis Foundation COVID-19 Task Force | <https://www.ncbi.nlm.nih.gov/pmc/articles/PMC7788316/> |
| Management of infants born to mothers with suspected or confirmed SARS-CoV-2 infection in the delivery room: A tentative proposal 2020 | Does not include public health recommendations | 2021 | Neonatal Resuscitation Committee, Japan Society of Perinatal, Neonatal Medicine | https://onlinelibrary.wiley.com/doi/10.1111/ped.14571 |
| An Italian Guidance Model for the Management of Suspected or Confirmed COVID-19 Patients in the Primary Care Setting | Does not include public health recommendations | 2020 | Noemi Lopes, Federica Vernuccio, Claudio Costantino, Claudia Imburgia, Cesare Gregoretti, Salvatore Salomone, Filippo Drago, Giuliano Lo Bianco | <https://www.frontiersin.org/articles/10.3389/fpubh.2020.572042/full> |
| Caribbean COVID-19 Recommendations | Does not include public health recommendations | 2022 | NR | NR |
| COVID-19 Diagnosis and Treatment Protocol (Revised trial eighth edition) | Does not include public health recommendations | 2021 | NR | <http://www.nhc.gov.cn/cms-search/downFiles/a449a3e2e2c94d9a856d5faea2ff0f94.pdf> |
| Best Practices for Managing COVID-19 Outbreaks in Acute Care Settings | Regional guideline | 2022 | Ontario Agency for Health Protection and Promotion (Public Health Ontario), Provincial Infectious Diseases Advisory Committee | <https://www.publichealthontario.ca/-/media/documents/ncov/ipac/2021/03/covid-19-pidac-outbreaks-acute-care.pdf?sc_lang=en> |
| Therapeutic Management of Adult Patients with COVID-19 | Regional guideline | 2022 | Ontario COVID-19 Drugs and Biologics Clinical Practice Guidelines Working Group | <https://covid19-sciencetable.ca/sciencebrief/clinical-practice-guideline-summary-recommended-drugs-and-biologics-in-adult-patients-with-covid-19-version-11-0/> |
| Rapid Antigen Tests for Voluntary Screen Testing | Regional guideline | 2021 | Ontario COVID-19 Science Advisory Table | <https://covid19-sciencetable.ca/wp-content/uploads/2021/12/Rapid-Antigen-Tests-for-Voluntary-Screen-Testing_published_20211210.pdf> |
| Evidence-Based Recommendations on the Use of Anti-SARS-CoV-2 Monoclonal Antibodies (Casirivimab + Imdevimab, and Sotrovimab) for Adults in Ontario | Regional guideline | 2021 | Ontario COVID-19 Science Advisory Table | <https://covid19-sciencetable.ca/sciencebrief/evidence-based-recommendations-on-the-use-of-anti-sars-cov-2-monoclonal-antibodies-casirivimab-imdevimab-and-sotrovimab-for-adults-in-ontario/> |
| Vaccine-Induced Immune Thrombotic Thrombocytopenia (VITT) Following Adenovirus Vector COVID-19 Vaccination: Interim Guidance for Healthcare Professionals in Emergency Department and Inpatient Settings | Regional guideline | 2021 | Ontario COVID-19 Science Advisory Table | <https://covid19-sciencetable.ca/wp-content/uploads/2021/05/Science-Brief_AstraZeneca_Hospital-Guide_version-2.0_20210510_published.pdf> |
| COVID-19 Guidance: Acute Care | Regional guideline | 2023 | Ontario Ministry of Health | <https://www.health.gov.on.ca/en/pro/programs/publichealth/coronavirus/docs/2019_acute_care_guidance.pdf> |
| Management of Cases and Contacts of COVID-19 in Ontario | Regional guideline | 2022 | Ontario Ministry of Health | <https://www.health.gov.on.ca/en/pro/programs/publichealth/coronavirus/docs/contact_mngmt/management_cases_contacts.pdf> |
| COVID-19 Vaccine Guidance | Regional guideline | 2022 | Ontario Ministry of Health | <https://www.health.gov.on.ca/en/pro/programs/publichealth/coronavirus/docs/vaccine/COVID-19_vaccine_administration.pdf> |
| COVID-19 guidance document for long-term care homes in Ontario | Regional guideline | 2022 | Ontario Ministry of Health | <https://www.ontario.ca/page/covid-19-guidance-document-long-term-care-homes-ontario> |
| COVID-19 Provincial Testing Guidance | Regional guideline | 2022 | Ontario Ministry of Health | <https://www.health.gov.on.ca/en/pro/programs/publichealth/coronavirus/docs/COVID-19_provincial_testing_guidance.pdf> |
| COVID-19 Guidance: Long-Term Care Homes and Retirement Homes for Public Health Units | Regional guideline | 2022 | Ontario Ministry of Health | <https://www.health.gov.on.ca/en/pro/programs/publichealth/coronavirus/docs/2019_LTC_homes_retirement_homes_for_PHUs_guidance.pdf> |
| COVID-19 Vaccine Booster Recommendations | Regional guideline | 2022 | Ontario Ministry of Health | <https://www.health.gov.on.ca/en/pro/programs/publichealth/coronavirus/docs/vaccine/COVID-19_vaccine_third_dose_recommendations.pdf> |
| COVID-19 Screening Tool for Long-Term Care Homes and Retirement Homes | Regional guideline | 2022 | Ontario Ministry of Health | <https://www.health.gov.on.ca/en/pro/programs/publichealth/coronavirus/docs/2019_screening_guidance.pdf> |
| COVID-19 Guidance: School Case, Contact, and Outbreak Management | Regional guideline | 2022 | Ontario Ministry of Health | <https://www.ontario.ca/document/covid-19-health-safety-and-operational-guidance-schools-2021-2022/management-covid-19-schools> |
| Guidance for Routine & Catch-Up Immunization Services | Regional guideline | 2022 | Ontario Ministry of Health | <https://www.health.gov.on.ca/en/pro/programs/publichealth/coronavirus/docs/Immunization_Services_during_COVID-19_08-26-2020.pdf> |
| COVID-19 Guidance: Personal Protective Equipment (PPE) for Health Care Workers and Health Care Entities | Regional guideline | 2022 | Ontario Ministry of Health | <https://www.health.gov.on.ca/en/pro/programs/publichealth/coronavirus/docs/ppe_guidance_hcw_hce.pdf> |
| COVID-19 Guidance: Congregate Living for Vulnerable Populations | Regional guideline | 2022 | Ontario Ministry of Health | <https://health.gov.on.ca/en/pro/programs/publichealth/coronavirus/docs/2019_congregate_living_guidance.pdf> |
| COVID-19 Guidance: Community Emergency Evacuations | Regional guideline | 2022 | Ontario Ministry of Health | <https://www.health.gov.on.ca/en/pro/programs/publichealth/coronavirus/docs/2019_community_emergency_evacuations_guidance.pdf> |
| COVID-19 Interim Guidance: Omicron Surge Management of Critical Staffing Shortages in Highest Risk Settings | Regional guideline | 2022 | Ontario Ministry of Health | <https://www.ontariomidwives.ca/sites/default/files/2022%2003%2031%20Omicron%20Critical%20Staffing%20Shortages.pdf> |
| COVID-19 Safety Guidelines for: Camps | Regional guideline | 2022 | Ontario Ministry of Health | <https://www.toronto.ca/wp-content/uploads/2022/03/977d-COVID-19-Safety-Guidelines-for-Camps-V-2.0-2022-03-01.pdf> |
| COVID-19 Guidance: Primary Care Providers in a Community Setting | Regional guideline | 2022 | Ontario Ministry of Health | <https://www.ontariomidwives.ca/sites/default/files/2021%2010%2019_MOH%20primary_care_guidance.pdf> |
| COVID-19: Interim Guidance for Schools and Child Care: Omicron Surge | Regional guideline | 2022 | Ontario Ministry of Health | <https://www.ucdsb.on.ca/common/pages/DisplayFile.aspx?itemId=34961523&msclkid=ce390d4dc14c11eca40406c25718981d> |
| Post COVID-19 Condition: Guidance for Primary Care | Regional guideline | 2021 | Ontario Ministry of Health | <https://www.ontariohealth.ca/sites/ontariohealth/files/2021-12/PostCovidConditionsClinicalGuidance_EN.pdf> |
| Guidance for Employers Managing Workers with Symptoms within 48 Hours of COVID-19 or Influenza Immunization | Regional guideline | 2021 | Ontario Ministry of Health | <https://www.health.gov.on.ca/en/pro/programs/publichealth/coronavirus/docs/guidance_for_screening_vaccinated_individuals.pdf> |
| COVID-19 Guidance: Considerations for Privately Initiated Testing | Regional guideline | 2021 | Ontario Ministry of Health | <https://www.health.gov.on.ca/en/pro/programs/publichealth/coronavirus/docs/Considerations_for_Privately-Initiated_Testing.pdf> |
| COVID-19 Variant of Concern: Case, Contact and Outbreak Management Interim Guidance (version 2.0) | Regional guideline | 2021 | Ontario Ministry of Health | <https://www.ontariomidwives.ca/sites/default/files/COVID-19%20ID%20Resources/2021%2002%2026%20VOC%20case%20and%20contact%20mgt%20v2.pdf> |
| Maternal-Neonatal COVID-19 General Guideline | Regional guideline | 2021 | Ontario Provincial Council for Maternal and Child Health (PCMCH) | <https://guides.hsict.library.utoronto.ca/COVID-19_Guidance_Documents/Pregnancy> |
| Clinical Guidelines: Long COVID-19 | Does not include public health recommendations | 2021 | Oregon Health & Science University (OHSU) | <https://www.ohsu.edu/sites/default/files/2021-04/Long-COVID-19-Clinical-Guidelines-English-April-21-2021.pdf> |
| Guidelines for Care of Critically Ill Adult Patients with COVID-19 in the Americas (Version 3) | Does not include public health recommendations | 2021 | Pan American Health Organization | <https://iris.paho.org/handle/10665.2/53895> |
| Guidelines for Prophylaxis and Management of Patients with Mild and Moderate COVID-19 in Latin America and the Caribbean | Does not include public health recommendations | 2021 | Pan American Health Organization | <https://iris.paho.org/handle/10665.2/55068> |
| Updated Guidance on Use and Prioritization of Monoclonal Antibody Therapy for Treatment of COVID-19 in Adolescents | Does not include public health recommendations | 2022 | Pediatric Infectious Diseases Society | <https://academic.oup.com/jpids/advance-article/doi/10.1093/jpids/piab124/6520293?login=false> |
| Multidisciplinary guidance regarding the use of immunomodulatory therapies for acute coronavirus disease 2019 in pediatric patients | Does not include public health recommendations | 2020 | Pediatric Infectious Diseases Society | <https://academic.oup.com/jpids/article/9/6/716/5893843> |
| Updated general recommendations in cancer management during the COVID-19 pandemic in the Philippines. | Does not include public health recommendations | 2020 | Philippine Society of Medical Oncology | <https://www.ncbi.nlm.nih.gov/pmc/articles/PMC7652546/pdf/can-14-1128.pdf> |
| Recommendations for interventional pulmonology during COVID-19 outbreak: a consensus statement from the Portuguese Pulmonology Society. | Does not include public health recommendations | 2020 | Portuguese Pulmonology Society | <https://www.sciencedirect.com/science/article/pii/S253104372030177X> |
| The expert consensus of proprietary Chinese medicines for the prevention and treatment of novel coronavirus pneumonia | Does not include public health recommendations | 2023 | Professional Committee of Respiratory Disease Drug Research, China Association of Chinese Medicine, World Federation of Chinese Medicine Societies Respiratory Committee, Professional Committee of Chronic Airway Diseases, China Medical Education Association | http://www.hansenzy.com/uploadfile/2022/0506/20220506021317945.pdf |
| Non-contact infrared thermometers (NCIT) | Does not include public health recommendations | 2021 | Public Health Agency Canada (PHAC) | <https://www.canada.ca/en/health-canada/services/drugs-health-products/covid19-industry/medical-devices/non-contact-infrared-thermometers.html> |
| For immunization providers: Interim national vaccine storage, handling and transportation guidelines for ultra-low temperature and frozen temperature COVID-19 vaccines | Does not include public health recommendations | 2022 | Public Health Agency of Canada (PHAC) | <https://www.canada.ca/en/public-health/services/diseases/2019-novel-coronavirus-infection/guidance-documents/vaccine-storage-handling-transportation-ultra-low-temperature-frozen.html> |
| Public health ethics framework: A guide for use in response to the COVID-19 pandemic in Canada | Does not include public health recommendations | 2022 | Public Health Agency of Canada (PHAC) | <https://www.canada.ca/en/public-health/services/diseases/2019-novel-coronavirus-infection/canadas-reponse/ethics-framework-guide-use-response-covid-19-pandemic.html> |
| Requirements for serological antibody tests submitted under the COVID-19 Interim Order: guidance | Does not include public health recommendations | 2022 | Public Health Agency of Canada (PHAC) | <https://www.canada.ca/en/health-canada/services/drugs-health-products/medical-devices/application-information/guidance-documents/covid19-requirements-serological-antibody-tests.html> |
| Pan-Canadian COVID-19 Testing and Screening Guidance: Technical guidance and implementation plan | Does not include public health recommendations | 2021 | Public Health Agency of Canada (PHAC) | <https://www.canada.ca/en/health-canada/services/drugs-health-products/covid19-industry/medical-devices/testing/pan-canadian-guidance.html> |
| Using Ventilation and Filtration to reduce aerosol transmission of COVID-19 in long-term care homes | Does not include public health recommendations | 2021 | Public Health Agency of Canada (PHAC) | <https://www.canada.ca/en/public-health/services/diseases/2019-novel-coronavirus-infection/guidance-documents/guide-ltch-ventilation-covid-19-pandemic.html> |
| COVID-19: Readiness criteria and indicators for easing restrictive public health measures | Does not include public health recommendations | 2021 | Public Health Agency of Canada (PHAC) | <https://www.canada.ca/en/public-health/services/diseases/2019-novel-coronavirus-infection/guidance-documents/readiness-criteria-indicators-easing-restrictive-public-health-measures.html> |
| Polymerase chain reaction (PCR) and cycle threshold (Ct) values in COVID-19 testing | Does not include public health recommendations | 2021 | Public Health Agency of Canada (PHAC) | <https://www.canada.ca/en/public-health/services/diseases/2019-novel-coronavirus-infection/guidance-documents/polymerase-chain-reaction-cycle-threshold-values-testing.html> |
| Reports of myocarditis and pericarditis after COVID-19 vaccination: Communiqué to health practitioners (June 3, 2021) | Does not include public health recommendations | 2021 | Public Health Agency of Canada (PHAC) | <https://www.canada.ca/en/public-health/services/diseases/2019-novel-coronavirus-infection/guidance-documents/reports-myocarditis-pericarditis-after-vaccination-communique-health-practitioners-june-3-2021.html> |
| How businesses and employees can stay safe while operating during COVID-19 | Does not include public health recommendations | 2023 | Public Health Agency of Canada (PHAC) | <https://www.canada.ca/en/public-health/services/diseases/2019-novel-coronavirus-infection/prevention-risks/guidance-workplaces-covid-19.html> |
| Outbreak Guidance: Workplace and Living Settings for Seasonal International Agriculture Workers (IAWs) | Regional guideline | 2022 | Public Health Agency of Canada (PHAC) | <https://www.health.gov.on.ca/en/pro/programs/publichealth/coronavirus/docs/COVID-19_Farm_Outbreak_guidance.pdf> |
| Individual and community-based measures to mitigate the spread of COVID-19 in Canada | Not a guideline | 2022 | Public Health Agency of Canada (PHAC) | <https://www.canada.ca/en/public-health/services/diseases/2019-novel-coronavirus-infection/health-professionals/public-health-measures-mitigate-covid-19.html> |
| Caring for Heritage Collections During the COVID-19 Pandemic | Does not include public health recommendations | 2021 | Public Health Agency of Canada (PHAC) | <https://www.canada.ca/en/conservation-institute/services/conservation-preservation-publications/canadian-conservation-institute-notes/caring-heritage-collections-covid19.html> |
| COVID-19 signs, symptoms and severity of disease: a clinician guide | Does not include public health recommendations | 2022 | Public Health Agency of Canada (PHAC) | <https://www.canada.ca/en/public-health/services/diseases/2019-novel-coronavirus-infection/guidance-documents/signs-symptoms-severity.html> |
| COVID-19: Prevention and risks | Not a guideline | 2022 | Public Health Agency of Canada (PHAC) | <https://www.canada.ca/en/public-health/services/diseases/2019-novel-coronavirus-infection/prevention-risks.html> |
| COVID-19: Travel, testing and borders | Not a guideline | 2022 | Public Health Agency of Canada (PHAC) | <https://travel.gc.ca/travel-covid> |
| Federal safety guidance to protect drivers and limit the spread of COVID-19 in commercial vehicle operations | Does not include public health recommendations | 2022 | Public Health Agency of Canada (PHAC) | <https://tc.canada.ca/en/initiatives/covid-19-measures-updates-guidance-issued-transport-canada/federal-safety-guidance-protect-drivers-limit-spread-covid-19-commercial-vehicle-operations> |
| Federal/provincial/territorial public health response plan for ongoing management of COVID-19 | Does not include public health recommendations | 2022 | Public Health Agency of Canada (PHAC) | <https://www.canada.ca/en/public-health/services/diseases/2019-novel-coronavirus-infection/guidance-documents/federal-provincial-territorial-public-health-response-plan-ongoing-management-covid-19.html> |
| Guidance on the use of influenza vaccine in the presence of COVID-19 | Does not include public health recommendations | 2022 | Public Health Agency of Canada (PHAC) | <https://www.canada.ca/en/public-health/services/immunization/national-advisory-committee-on-immunization-naci/guidance-use-influenza-vaccine-covid-19.html> |
| Interim guidance on the use of rapid antigen detection tests for the identification of SARS-CoV-2 infection | Does not include public health recommendations | 2022 | Public Health Agency of Canada (PHAC) | <https://www.canada.ca/en/public-health/services/diseases/2019-novel-coronavirus-infection/guidance-documents/use-rapid-antigen-detection-tests.html> |
| Reducing COVID-19 risk in community settings: A tool for operators | Not a guideline | 2022 | Public Health Agency of Canada (PHAC) | https://health.canada.ca/en/public-health/services/diseases/2019-novel-coronavirus-infection/guidance-documents/reducing-covid-19-risk-community-settings-tool-operators.html |
| Summary of evidence supporting COVID-19 public health measures | Not a guideline | 2023 | Public Health Agency of Canada (PHAC) | <https://www.canada.ca/en/public-health/services/diseases/2019-novel-coronavirus-infection/guidance-documents/summary-evidence-supporting-covid-19-public-health-measures.html> |
| Planning for the 2021-2022 school year in the context of COVID-19 vaccination | Not a guideline | 2022 | Public Health Agency of Canada (PHAC) | <https://www.canada.ca/en/public-health/services/diseases/2019-novel-coronavirus-infection/guidance-documents/planning-2021-2022-school-year-vaccination.html> |
| Planning guidance for immunization clinics for COVID-19 vaccines | Does not include public health recommendations | 2022 | Public Health Agency of Canada (PHAC) | <https://www.canada.ca/en/public-health/services/diseases/2019-novel-coronavirus-infection/guidance-documents/planning-immunization-clinics-covid-19-vaccines.html> |
| Infection prevention and control for COVID-19: Interim guidance for acute healthcare settings | Does not include public health recommendations | 2022 | Public Health Agency of Canada (PHAC) | <https://www.canada.ca/en/public-health/services/diseases/2019-novel-coronavirus-infection/health-professionals/infection-prevention-control-covid-19-second-interim-guidance.html> |
| Recommendations: co-administration of COVID-19 vaccines in children 5-11 years | Regional guideline | 2022 | Public Health Ontario | <https://www.publichealthontario.ca/-/media/Documents/nCoV/Vaccines/2022/05/oiac-recommendations-co-covid-19-vaccines-children.pdf?rev=9496e2af27184483a26cb3df0dbda201&sc_lang=en> |
| Ethical Considerations Related to Projects Involving Direct Interaction with Participants during the COVID-19 Pandemic | Regional guideline | 2023 | Public Health Ontario (PHO) | <https://www.publichealthontario.ca/-/media/documents/ncov/ethics-review-board-guidelines-covid19.pdf?sc_lang=fr#:~:text=Project%20teams%20should%20consider%20the,when%20sharing%20findings%20with%20participants.> |
| IPAC Recommendations for Use of Personal Protective Equipment for Care of Individuals with Suspect or Confirmed COVID‑19 | Regional guideline | 2022 | Public Health Ontario (PHO) | <https://www.publichealthontario.ca/-/media/documents/ncov/updated-ipac-measures-covid-19.pdf?la=en> |
| Multidisciplinary Recommendations Regarding Post-Vaccine Adenopathy and Radiologic Imaging Radiology Scientific Expert Panel | Does not include public health recommendations | 2021 | Radiological Society of North America (RSNA) | <https://www.ncbi.nlm.nih.gov/pmc/articles/PMC7909071/> |
| A Regional Canadian Expert Consensus on recommendations for restoring exercise and pulmonary function testing in low and moderate-high community COVID-19 prevalence settings | Regional guideline | 2020 | Regional Canadian Expert Consensus | <https://www.cambridge.org/core/journals/infection-control-and-hospital-epidemiology/article/regional-canadian-expert-consensus-on-recommendations-for-restoring-exercise-and-pulmonary-function-testing-in-low-and-moderatehigh-community-covid19-prevalence-settings/3DE66ECC8346C1D7C61DDB8AB0FAA3AD> |
| Guide to the suspected or confirmed COVID-19 patient journey, quick response | Does not include public health recommendations | 2022 | Republic of Tunisia Ministry of Health (INEAS) | <https://www.ineas.tn/sites/default/files/gpc_covid_19_version_11_mai_2021.pdf> |
| Consensus statement and recommendations on the treatment of COVID-19: 2021 update. | Does not include public health recommendations | 2021 | Research Center for Epidemic Prevention - National Yang Ming Chiao Tung University (RCEP-NYCU) | <https://journals.lww.com/jcma/Abstract/9000/Consensus_statement_and_recommendations_on_the.99483.aspx> |
| RHRA Recommendation for Asymptomatic COVID-19 Screen Testing for Retirement Homes | Regional guideline | 2021 | Retirement Homes Regulatory Authority (RHRA) | <https://www.rhra.ca/wp-content/uploads/2021/07/RHRA-Recommendation-for-Asymptomatic-COVID-19-Screen-Testing-for-Retirement-Homes-July-14-2021.pdf> |
| Proposed delay for safe surgery after COVID-19. | Does not include public health recommendations | 2021 | Royal Australian College of Surgeons | <https://www.ncbi.nlm.nih.gov/pmc/articles/PMC7955821/> |
| The Saudi Critical Care Society extracorporeal life support chapter guidance on utilization of veno-venous extracorporeal membrane oxygenation in adults with acute respiratory distress syndrome and special considerations in the era of coronavirus disease 2019. | Does not include public health recommendations | 2021 | Saudi Critical Care Society | <https://smj.org.sa/content/42/6/589> |
| Prevention and management of venous thromboembolism in patients with COVID-19 | Does not include public health recommendations | 2021 | Scottish Intercollegiate Guidelines Network (SIGN) | <https://www.sign.ac.uk/media/1900/sign-163-rg-vte.pdf> |
| COVID-19 position statement: Presentations and management of COVID-19 in older people in acute care | Does not include public health recommendations | 2021 | Scottish Intercollegiate Guidelines Network (SIGN) | <https://www.sign.ac.uk/media/1826/presentations-and-management-of-covid-19-in-older-people-v2-final.pdf> |
| COVID-19 position statement: reducing the risk of postoperative mortality due to COVID-19 in patients undergoing elective surgery | Does not include public health recommendations | 2020 | Scottish Intercollegiate Guidelines Network (SIGN) | <https://www.sign.ac.uk/media/1822/elective-surgery_v2.pdf> |
| COVID-19 position statement: the prevention and management of thromboembolism in hospitalised patients with COVID-19-related disease. | Does not include public health recommendations | 2020 | Scottish Intercollegiate Guidelines Network (SIGN) | <https://www.sign.ac.uk/media/1691/sg_prevention_of_thromboembolism_in_hospitalised_patients.pdf> |
| Prevention of circuit thrombosis in adult inpatients who are COVID-19 positive and undergoing renal replacement therapy (RRT) on critical care wards | Does not include public health recommendations | 2020 | Scottish Intercollegiate Guidelines Network (SIGN) | <https://www.sign.ac.uk/media/1632/sg_prevention_of_thrombosis_in_rrt.pdf> |
| COVID-19 position statement: management of patients attending an endoscopy unit for any endoscopic procedure not requiring general anaesthesia (bronchoscopy, cystoscopy, upper gastrointestinal and lower gastrointestinal endoscopy). | Does not include public health recommendations | 2021 | Scottish Intercollegiate Guidelines Network (SIGN) | <https://www.sign.ac.uk/media/1818/endoscopy-guidance_v11-final.pdf> |
| Urgent and elective proctologic/anorectal interventions in the COVID-19 pandemic: A practical guideline for treatment safety | Does not include public health recommendations | 2021 | Sezai Leventoğlu, Bülent Menteş, Esin Şenol, David Zimmerman, Gianluca Pellino, Eloy Espin | <https://jag.journalagent.com/travma/pdfs/UTD_27_2_180_186.pdf> |
| Interpretation of the local standard "Technical specification for collection of 2019-nCov samples on surface of objects" | Does not include public health recommendations | 2021 | Sheng Xin, Wang Bing, Yang Lihua, Jia Lei | <https://rs.yiigle.com/cmaid/1327541> |
| Expert consensus on the prevention of novel coronavirus pneumonia in the winter and spring of 2022 in Shenzhen | Does not include public health recommendations | 2022 | Shenzhen Medical Quality Control Center for Preventive Treatment of Traditional Chinese Medicine Professional Committee of Infectious Diseases, Shenzhen Society of Traditional Chinese Medicine | <https://oversea.cnki.net/KCMS/detail/detail.aspx?dbcode=CAPJ&dbname=CAPJLAST&filename=JZYB20220314006&uniplatform=OVERSEAS_EN&v=MSKsDCnlU4V2RRa0MUfVLgdfvEnQ-VbKG3v26xgrvHbyY5mRArLTorWWJhfvf1RN> |
| Recommendations for COVID-19 vaccination in people with rheumatic disease: Developed by the Singapore Chapter of Rheumatologists. | Does not include public health recommendations | 2021 | Singapore Chapter of Rheumatologists | <https://www.ncbi.nlm.nih.gov/pmc/articles/PMC8207070/pdf/APL-24-746.pdf> |
| Consensus of the Genetics Branch of the Chilean Society of Pediatrics on the prioritization of people with Down syndrome and rare diseases for vaccination against SARS-CoV-2 | Does not include public health recommendations | 2021 | Sociedad Chilena De Pediatria | <https://www.scielo.cl/scielo.php?pid=S2452-60532021005000411&script=sci_abstract&tlng=en> |
| The heart and COVID-19: what cardiologists need to know | Does not include public health recommendations | 2020 | Sociedade Brasileira de Cardiologia | <http://abccardiol.org/en/article/the-heart-and-covid-19-what-cardiologists-need-to-know/> |
| Update of the recommendations of the Sociedade Portuguesa de Cuidados Intensivos and the Infection and Sepsis Group for the approach to COVID-19 in Intensive Care Medicine | Does not include public health recommendations | 2022 | Sociedade Portuguesa de Cuidados Intensivos and the Infection and Sepsis Group | <https://www.scielo.br/j/rbti/a/xt84HRsVSCyvYVLs4wGrRkz/abstract/?lang=en> |
| Management of Acute Myocardial Infarction During the COVID-19 Pandemic. | Does not include public health recommendations | 2020 | Society for Cardiovascular Angiography and Interventions (SCAI), American College of Cardiology (ACC), American College of Emergency Physicians (ACEP) | https://onlinelibrary.wiley.com/doi/10.1002/ccd.28946 |
| Society for Cardiovascular Magnetic Resonance (SCMR) recommended CMR protocols for scanning patients with active or convalescent phase COVID-19 infection | Does not include public health recommendations | 2020 | Society for Cardiovascular Magnetic Resonance (SCMIR) | <https://link.springer.com/content/pdf/10.1186/s12968-020-00656-6.pdf> |
| Provider considerations for engaging in COVID-19 vaccine counseling with pregnant and lactating patients | Does not include public health recommendations | 2021 | Society for Maternal-Fetal Medicine (SMFM) | <https://www.smfm.org/publications/390-provider-considerations-for-engaging-in-covid-19-vaccine-counseling-with-pregnant-and-lactating-patients> |
| Pediatric Airway Management in COVID-19 patients - Consensus Guidelines from the Society for Pediatric Anesthesia's Pediatric Difficult Intubation Collaborative and the Canadian Pediatric Anesthesia Society. | Does not include public health recommendations | 2020 | Society for Pediatric Anesthesia's Pediatric Difficult Intubation Collaborative, Canadian Pediatric Anesthesia Society | https://www.ncbi.nlm.nih.gov/pmc/articles/PMC7173403/ |
| Difficult Airway Management in Adult COVID-19 Patients: Statement by the Society of Airway Management | Does not include public health recommendations | 2021 | Society of Airway Management | <https://journals.lww.com/anesthesia-analgesia/Fulltext/2021/10000/Difficult_Airway_Management_in_Adult_Coronavirus.10.aspx> |
| Management considerations for pregnant patients with COVID-19 | Does not include public health recommendations | 2021 | Society of Maternal & Fetal Medicine | <https://s3.amazonaws.com/cdn.smfm.org/media/2734/SMFM_COVID_Management_of_COVID_pos_preg_patients_2-2-21_(final).pdf> |
| Perioperative Coronavirus Vaccination-Timing and Implications: A Guidance Document | Does not include public health recommendations | 2021 | Society of Thoracic Surgeons Workforce on Critical Care | <https://pubmed.ncbi.nlm.nih.gov/34370980/> |
| Diagnostic-therapeutic recommendations of the group of experts from FACME ad-hoc on the management of related cerebral venous thrombosis with vaccination against COVID-19 | Does not include public health recommendations | 2021 | Spanish Federation of Medical and Scientific Associations [Federación de Asociaciones Científico Médicas Espanolas (FACME)] | <https://www.elsevier.es/es-revista-neurologia-295-articulo-recomendaciones-diagnostico-terapeuticas-del-grupo-trabajo-S0213485321000839> |
| Management of hyponatraemia and hypernatraemia during the Covid-19 pandemic: a consensus statement of the Spanish Society for Endocrinology (Acqua Neuroendocrinology Group). | Does not include public health recommendations | 2021 | Spanish Society for Endocrinology (Acqua Neuroendocrinology Group) | <https://link.springer.com/article/10.1007/s11154-021-09627-3> |
| Key issues in emergency department management of COVID-19: proposals for improving care for patients in Latin America | Does not include public health recommendations | 2020 | Spanish Society of Emergency Medicine (SEMES), Latin American Federation of Emergency Medicine (FLAME) | http://emergencias.portalsemes.org/descargar/puntos-clave-sobre-la-covid19-en-los-servicios-de-urgencias-propuestas-de-mejora-para-su-atencin-en-latinoamrica/english/ |
| Recommendations for the management of critically ill patients with COVID-19 in Intensive Care Units | Does not include public health recommendations | 2021 | Spanish Society of Intensive and Critical Care Medicine and Coronary Units [Sociedad Española de Medicina Intensiva, Crítica y Unidades Coronarias (SEMI-CYUC)] | <https://www.sciencedirect.com/science/article/pii/S217357272100179X> |
| Recommendations for ophthalmologic practice during the easing of COVID-19 control measures | Does not include public health recommendations | 2021 | Spanish Society of Ophthalmology | <https://onlinelibrary.wiley.com/doi/epdf/10.1111/aos.14752> |
| Perioperative and critical care concerns in coronavirus pandemic. | Does not include public health recommendations | 2020 | Sukhminder Jit Singh Bajwa, Rashi Sarna, Chashamjot Bawa, Lalit Mehdiratta | <https://www.ncbi.nlm.nih.gov/pmc/articles/PMC7189905/> |
| Surviving Sepsis Campaign Guidelines on the Management of Adults With Coronavirus Disease 2019 (COVID-19) in the ICU: First Update. | Does not include public health recommendations | 2021 | Surviving Sepsis Campaign (SSC) | <https://journals.lww.com/ccmjournal/Fulltext/2021/03000/Surviving_Sepsis_Campaign_Guidelines_on_the.21.aspx?casa_token=5O7X2d-yP8MAAAAA:1W3jcN5fUZreINSBFACfawzN4EIs_drJHWvIeurFbAXNpoqu1mVYyJWz3faqv9R49ld6YJI1ZS4fVgAlItjW28us> |
| Triage for intensive care treatment under scarcity | Does not include public health recommendations | 2020 | Swiss Academy of Medical Sciences | <https://smw.ch/index.php/smw/article/view/2937/4834> |
| Swiss Recommendations for the Follow-Up and Treatment of Pulmonary Long COVID | Does not include public health recommendations | 2021 | Swiss COVID Lung Study Group, Swiss Society of Pulmonology (SSP) | <https://www.karger.com/Article/FullText/517255> |
| Rehabilitation programs for patients with COronaVIrus Disease 2019: consensus statements of Taiwan Academy of Cardiovascular and Pulmonary Rehabilitation | Does not include public health recommendations | 2020 | Taiwan Academy of Cardiovascular and Pulmonary Rehabilitation | https://www.sciencedirect.com/science/article/pii/S0929664620303892?via%3Dihub |
| Recommendations for inpatient treatment of patients with COVID-19 | Does not include public health recommendations | 2022 | The Association of the Scientific Medical Societies of Germany (AWMF) | https://register.awmf.org/de/leitlinien/detail/113-001LG |
| AWMF S1-Leitlinie Long/ Post-COVID | Does not include public health recommendations | 2022 | The Association of the Scientific Medical Societies of Germany (AWMF), German Society for Pneumology and Respiratory Medicine | https://www.awmf.org/uploads/tx_szleitlinien/020-027l_S1_Post_COVID_Long_COVID_2022-08.pdf |
| Multi-organ point-of-care ultrasound for COVID-19 (PoCUS4COVID): international expert consensus | Does not include public health recommendations | 2020 | The European Society of Radiology | https://www.ncbi.nlm.nih.gov/pmc/articles/PMC7759024/#CR47 |
| Critical Care for COVID-19 Affected Patients: Updated Position Statement of the Indian Society of Critical Care Medicine | Does not include public health recommendations | 2020 | The Indian Society of Critical Care Medicine (ISCCM) | https://www.ncbi.nlm.nih.gov/pmc/articles/PMC7724933/ |
| Towards a standardised method of patient prioritisation that accounts for clinical harm | Does not include public health recommendations | 2021 | The Royal College of Surgeons of England (RCS) | https://www.ncbi.nlm.nih.gov/pmc/articles/PMC8004326/ |
| Consensus statement: Safe Airway Society principles of airway management and tracheal intubation specific to the COVID-19 adult patient group | Does not include public health recommendations | 2020 | The Safe Airway Society | https://www.ncbi.nlm.nih.gov/pmc/articles/PMC7267410/ |
| The Saudi Critical Care Society practice guidelines on the management of COVID-19 in the ICU: Therapy Section | Does not include public health recommendations | 2021 | The Saudi Critical Care Society | https://www.sciencedirect.com/science/article/pii/S1876034121003269?via%3Dihub |
| Expert advice on management of urban nucleic acid testing base | Does not include public health recommendations | 2021 | Tianjin Medical Doctor Association | https://rs.yiigle.com/CN112150202106/1326332.htm |
| Integrating Chinese andwestern medicine for COVID-19: A living evidence-based guideline (version 1) | Does not include public health recommendations | 2021 | Trust-worthy Traditional Chinese Medicine Recommendations (TCMRecs) working group | https://onlinelibrary.wiley.com/doi/full/10.1111/jebm.12444?casa_token=8LMElW5mPY0AAAAA%3AcYwydpZWDesRz1riiT2uq0DRmlxbVLWGDG5wVCwyO_umG9AJFOWfRcwbAdkUOOFtkq14j0L134_tZTwG |
| Framework for reopening schools | Not a guideline | 2020 | United Nations Educational, Scientific and Cultural Organization (UNESCO), United Nations International Children's Emergency Fund (UNICEF), World Bank, World Food Programme, United Nations High Commissioner for Refugees (UNHCR) | https://www.unicef.org/documents/framework-reopening-schools |
| Temperature-sensitive health products in the expanded programme on immunization cold chain: a WHO-UNICEF joint statement encouraging greater health commodity supply chain integration for temperature-sensitive pharmaceuticals where appropriate, 19 November 2020 Screen reader support enabled. | Does not include public health recommendations | 2020 | United Nations International Children's Emergency Fund (UNICEF), World Health Organization (WHO) | https://apps.who.int/iris/handle/10665/336748 |
| Addressing Post-COVID Symptoms: A Guide for Primary Care Physicians | Does not include public health recommendations | 2021 | University of Michigan’s Campus Health Response Committee, Michigan Medicine Hospital System | https://www.jabfm.org/content/jabfp/34/6/1229.full.pdf |
| A Delphi-Based Consensus Statement: Recommendation for Physiotherapy Management and Rehabilitation of People Living with Long COVID in Bangladesh | Does not include public health recommendations | 2021 | Unnamed multi-disciplinary, international working group | https://papers.ssrn.com/sol3/papers.cfm?abstract_id=3969173 |
| Remdesivir for severe covid-19: a clinical practice guideline | Does not include public health recommendations | 2020 | Unnamed multi-disciplinary, international working group | https://www.bmj.com/content/370/bmj.m2924#:~:text=Recommendations%20The%20guideline%20panel%20makes,randomised%20controlled%20trials%20examining%20remdesivir. |
| Guidelines for the prevention and management of children and adolescents with COVID‑19 | Does not include public health recommendations | 2022 | Unnamed multi-disciplinary, international working group | https://doi.org/10.1007/s00431-022-04615-4 |
| Inpatient obstetric management of COVID-19. | Does not include public health recommendations | 2020 | Unnamed multi-disciplinary, international working group (ACOG, SMFM...) | https://www.ncbi.nlm.nih.gov/pmc/articles/PMC7373047/ |
| Guidance for the Management of Patients with Vascular Disease or Cardiovascular Risk Factors and COVID-19: Position Paper from VAS-European Independent Foundation in Angiology/Vascular Medicine | Does not include public health recommendations | 2020 | VAS-European Independent Foundation in Angiology/Vascular Medicine | <https://pubmed.ncbi.nlm.nih.gov/32920811/> |
| Perinatal COVID-19: review of current evidence and practical approach towards prevention and management. | Not a guideline | 2020 | Venkateshwarlu Vardhelli, Aakash Pandita, Anish Pillai, Susanta Kumar Badatya | <https://link.springer.com/article/10.1007/s00431-020-03866-3> |
| Point of view on the vaccination against COVID-19 in patients with autoimmune inflammatory rheumatic diseases | Does not include public health recommendations | 2021 | Victoria Furer, Christien Rondaan, Nancy Agmon-Levin, Sander van Assen, Marc Bijl, Meliha Crnkic Kapetanovic, Annette de Thurah, Ulf Mueller-Ladner, Daphna Paran, Karen Schreiber, Klaus Warnatz, Nico M. Wulffraat, Ori Elkayam | <https://rmdopen.bmj.com/content/7/1/e001594.info> |
| Safeguarding pregnant asylum-seekers and refugees during the era of COVID-19 | Not a guideline | 2021 | Weill Cornell's Department of Anesthesiology | <https://www.ncbi.nlm.nih.gov/pmc/articles/PMC7914379/> |
| COVID‑19 and idiopathic nephrotic syndrome in children: systematic review of the literature and recommendations from a highly affected area | Does not include public health recommendations | 2021 | Working group of pediatric nephrologists (Italy, name not specified) | <https://link.springer.com/article/10.1007%2Fs00467-021-05330-2> |
| Drugs to prevent COVID-19: living guideline | Does not include public health recommendations | 2023 | World Health Organization (WHO) | <https://www.who.int/publications/i/item/WHO-2019-nCoV-prophylaxes-2023.1> |
| Evaluation of COVID-19 vaccine effectiveness: interim guidance, 17 March 2021 | Does not include public health recommendations | 2022 | World Health Organization (WHO) | <https://apps.who.int/iris/bitstream/handle/10665/340301/WHO-2019-nCoV-vaccine-effectiveness-measurement-2021.1-rus.pdf> |
| Therapeutics and COVID-19: living guideline | Does not include public health recommendations | 2022 | World Health Organization (WHO) | <https://apps.who.int/iris/rest/bitstreams/1467050/retrieve> |
| Mental health and psychosocial support aspects of the COVID-19 response. | Does not include public health recommendations | 2022 | World Health Organization (WHO) | <https://apps.who.int/iris/bitstream/handle/10665/331927/Mental-health-COVID-19-eng.pdf?sequence=5&isAllowed=y> |
| Maintaining infection prevention and control measures for COVID-19 in health care facilities: Policy brief, 7 June 2022 | Does not include public health recommendations | 2022 | World Health Organization (WHO) | <https://apps.who.int/iris/rest/bitstreams/1429794/retrieve> |
| COVID-19 and mandatory vaccination: ethical considerations | Does not include public health recommendations | 2022 | World Health Organization (WHO) | <https://apps.who.int/iris/handle/10665/354585?search-result=true&query=COVID-19+and+mandatory+vaccination%3A+Ethical+considerations&scope=&rpp=10&sort_by=score&order=desc> |
| Indicators to monitor health-care capacity and utilization for decision-making on COVID-19 | Does not include public health recommendations | 2022 | World Health Organization (WHO) | <https://apps.who.int/iris/handle/10665/333754> |
| Interim guidance note for hospitals : managing hospital services, maintaining essential routine health care and generating surge capacity | Does not include public health recommendations | 2022 | World Health Organization (WHO) | <https://apps.who.int/iris/handle/10665/332381> |
| Risk assessment and management of health-care workers in the context of COVID-19 | Does not include public health recommendations | 2022 | World Health Organization (WHO) | <https://apps.who.int/iris/handle/10665/334366> |
| Injection safety in the context of coronavirus disease (COVID-19) vaccination | Does not include public health recommendations | 2022 | World Health Organization (WHO) | <https://www.who.int/publications/i/item/WHO-2019-nCoV-Policy_brief-Vaccination-Injection_safety-Addendum-2022.1> |
| Repurposing facilities for isolation and management of mild COVID-19 cases | Does not include public health recommendations | 2022 | World Health Organization (WHO) | <http://iris.wpro.who.int/handle/10665.1/14528> |
| Digital documentation of COVID-19 certificates: test result: technical specifications and implementation guidance, 31 March 2022 | Does not include public health recommendations | 2022 | World Health Organization (WHO) | <https://www.who.int/publications/i/item/WHO-2019-nCoV-Digital_certificates_diagnostic_test_results-2022.1> |
| In the wake of the pandemic. Preparing for Long COVID. | Does not include public health recommendations | 2022 | World Health Organization (WHO) | <https://apps.who.int/iris/bitstream/handle/10665/339629/Policy-brief-39-1997-8073-eng.pdf> |
| Strengthening COVID-19 vaccine demand and uptake in refugees and migrants | Does not include public health recommendations | 2022 | World Health Organization (WHO) | <https://www.who.int/publications/i/item/WHO-2019-nCoV-immunization-demand_planning-refugees_and_migrants-2022.1> |
| Use of SARS-CoV-2 antigen-detection rapid diagnostic tests for COVID-19 self-testing "Update coming soon" | Does not include public health recommendations | 2022 | World Health Organization (WHO) | <https://apps.who.int/iris/handle/10665/352350> |
| Operational guidance on establishing an ultra-cold chain system in support of the Pfizer-BioNTech COVID-19 Vaccine rollout | Does not include public health recommendations | 2022 | World Health Organization (WHO) | <https://www.who.int/publications/i/item/WHO-2019-nCoV-UCC_systems-Pfizer-BioNTech_vaccine-2022.1> |
| Interim recommendations for heterologous COVID-19 vaccine schedules Interim guidance | Does not include public health recommendations | 2021 | World Health Organization (WHO) | <https://www.who.int/publications/i/item/WHO-2019-nCoV-vaccines-SAGE-recommendation-heterologous-schedules> |
| Assessing the impact of COVID-19 on older people in the African region | Does not include public health recommendations | 2021 | World Health Organization (WHO) | <https://apps.who.int/iris/handle/10665/351134> |
| Key planning recommendations for mass gatherings in the context of COVID-19 | Does not include public health recommendations | 2021 | World Health Organization (WHO) | <https://apps.who.int/iris/rest/bitstreams/1387941/retrieve> |
| Interim recommendations for an extended primary series with an additional vaccine dose for COVID-19 vaccination in immunocompromised persons | Does not include public health recommendations | 2021 | World Health Organization (WHO) | <https://apps.who.int/iris/handle/10665/347079> |
| Co administration of seasonal inactivated influenza and COVID-19 vaccines | Does not include public health recommendations | 2021 | World Health Organization (WHO) | <https://www.who.int/publications/i/item/WHO-2019-nCoV-vaccines-SAGE_recommendation-coadministration-influenza-vaccines> |
| Antigen-detection in the diagnosis of SARS-CoV-2 infection | Does not include public health recommendations | 2021 | World Health Organization (WHO) | <https://www.who.int/publications/i/item/antigen-detection-in-the-diagnosis-of-sars-cov-2infection-using-rapid-immunoassays> |
| Advancing tobacco control during the COVID-19 pandemic-MPOWER implementation Tobacco Free Initiative | Does not include public health recommendations | 2021 | World Health Organization (WHO) | <https://apps.who.int/iris/handle/10665/352319> |
| COVID-19 vaccine post-introduction evaluation (cPIE) guide: interim guidance, 25 August 2021 | Does not include public health recommendations | 2021 | World Health Organization (WHO) | <https://apps.who.int/iris/handle/10665/344721> |
| Guidance for clinical case management of thrombosis with thrombocytopenia syndrome (TTS) following vaccination to prevent coronavirus disease (COVID-19) | Does not include public health recommendations | 2021 | World Health Organization (WHO) | <https://apps.who.int/iris/handle/10665/342999> |
| Guidance on utilization of COVID-19 vaccines before the date of expiry, 19 July 2021 | Does not include public health recommendations | 2021 | World Health Organization (WHO) | <https://apps.who.int/iris/handle/10665/343035> |
| WHO information note: COVID-19: considerations for tuberculosis (TB) care, 5 May 2021 | Does not include public health recommendations | 2021 | World Health Organization (WHO) | <https://apps.who.int/iris/handle/10665/341126> |
| Meeting of the Strategic Advisory Group of Experts on Immunization, 22–24 March 2021: conclusions and recommendations | Does not include public health recommendations | 2021 | World Health Organization (WHO) | <https://apps.who.int/iris/handle/10665/341624> |
| Evidence review – Public health measures in the aviation sector in the context of COVID-19: quarantine and isolation (‎21 May 2021)‎ – Examen des données factuelles – Mesures de santé publique dans le secteur du transport aérien dans le contexte de la COVID-19: quarantaine et isolement (‎21 mai 2021)‎ | Does not include public health recommendations | 2021 | World Health Organization (WHO) | <https://apps.who.int/iris/handle/10665/341437> |
| Implementing telemedicine services during COVID-19 : guiding principles and considerations for a stepwise approach | Does not include public health recommendations | 2021 | World Health Organization (WHO) | <https://apps.who.int/iris/handle/10665/336862> |
| Estimating COVID-19 vaccine effectiveness against severe acute respiratory infections (SARI) hospitalisations associated with laboratory-confirmed SARS-CoV-2: an evaluation using the test-negative design: guidance document | Does not include public health recommendations | 2021 | World Health Organization (WHO) | <https://apps.who.int/iris/handle/10665/341111> |
| Operational guidance: COVID-19 immunization service delivery modalities | Does not include public health recommendations | 2021 | World Health Organization (WHO) | <https://apps.who.int/iris/bitstream/handle/10665/341118/WHO-EURO-2021-2404-42159-58089-eng.pdf> |
| Data for action: achieving high uptake of COVID-19 vaccines: gathering and using data on the behavioural and social drivers of vaccination: a guidebook for immunization programmes and implementing partners: interim guidance, 1 April 2021 | Does not include public health recommendations | 2021 | World Health Organization (WHO) | <https://apps.who.int/iris/handle/10665/340645> |
| Roadmap to improve and ensure good indoor ventilation in the context of COVID-19 | Does not include public health recommendations | 2021 | World Health Organization (WHO) | <https://apps.who.int/iris/handle/10665/339857> |
| Health workers in focus: policies and practices for successful public response to COVID-19 vaccination: strategic considerations for member states in the WHO European region | Does not include public health recommendations | 2021 | World Health Organization (WHO) | <https://apps.who.int/iris/handle/10665/339854> |
| Maintaining a safe and adequate blood supply and collecting convalescent plasma in the context of the COVID-19 pandemic: interim guidance, 17 February 2021 | Does not include public health recommendations | 2021 | World Health Organization (WHO) | <https://apps.who.int/iris/handle/10665/339793> |
| COVID-19 strategic preparedness and response plan: 1 February 2021 to 31 January 2022 | Does not include public health recommendations | 2021 | World Health Organization (WHO) | <https://apps.who.int/iris/handle/10665/340072> |
| COVID-19: occupational health and safety for health workers: interim guidance, 2 February 2021 | Does not include public health recommendations | 2021 | World Health Organization (WHO) | <https://apps.who.int/iris/handle/10665/339151> |
| Operational guidance: legal and regulatory framework facilitating vaccine deployment | Does not include public health recommendations | 2021 | World Health Organization (WHO) | <https://apps.who.int/iris/handle/10665/339391?show=full> |
| Risk communication and community engagement for COVID-19 contact tracing: interim guidance | Does not include public health recommendations | 2021 | World Health Organization (WHO) | <https://apps.who.int/iris/handle/10665/339100> |
| Acceptance and demand for COVID-19 vaccines: Interim guidance, 31 January 2021 | Does not include public health recommendations | 2021 | World Health Organization (WHO) | <https://apps.who.int/iris/handle/10665/339449> |
| Considerations for forming a regional COVID-19 review committee (‎‎RRC)‎‎: technical brief, 29 January 2021 | Does not include public health recommendations | 2021 | World Health Organization (WHO) | <https://apps.who.int/iris/handle/10665/339153> |
| Operational guidance: acceptance and uptake of COVID-19 vaccines | Does not include public health recommendations | 2021 | World Health Organization (WHO) | <https://apps.who.int/iris/handle/10665/338855> |
| Surveillance case definitions for human infection with novel coronavirus (‎nCoV)‎, interim guidance, 15 January 2020 | Does not include public health recommendations | 2021 | World Health Organization (WHO) | <https://apps.who.int/iris/handle/10665/332412> |
| SARS-CoV-2 genomic sequencing for public health goals | Does not include public health recommendations | 2021 | World Health Organization (WHO) | <https://apps.who.int/iris/handle/10665/338483> |
| Rational use of personal protective equipment for COVID-19 and considerations during severe shortages: interim guidance, 23 December 2020 | Does not include public health recommendations | 2020 | World Health Organization (WHO) | <https://apps.who.int/iris/handle/10665/338033> |
| COVID-19 global risk communication and community engagement strategy | Does not include public health recommendations | 2020 | World Health Organization (WHO) | <https://apps.who.int/iris/handle/10665/338057> |
| SARS-CoV-2 antigen-detecting rapid diagnostic tests: an implementation guide | Does not include public health recommendations | 2020 | World Health Organization (WHO) | <https://apps.who.int/iris/handle/10665/337948> |
| Regional guiding framework for risk communication and community engagement for the COVID-19 response in the Eastern Mediterranean Region/Middle East and North Africa: December 2020 | Does not include public health recommendations | 2020 | World Health Organization (WHO) | <https://apps.who.int/iris/handle/10665/342030> |
| Technical specifications of personal protective equipment for COVID-19: interim guidance, 13 November 2020 | Does not include public health recommendations | 2020 | World Health Organization (WHO) | <https://apps.who.int/iris/handle/10665/336622> |
| Pandemic fatigue: reinvigorating the public to prevent COVID-19: policy framework for supporting pandemic prevention and management | Does not include public health recommendations | 2020 | World Health Organization (WHO) | <https://apps.who.int/iris/handle/10665/337574> |
| Promoting the health of migrant workers in the WHO European Region during COVID-19. Interim guidance, 6 November 2020 | Does not include public health recommendations | 2020 | World Health Organization (WHO) | <https://apps.who.int/iris/handle/10665/336549> |
| Prevention, identification and management of health worker infection in the context of COVID-19: interim guidance, 30 October 2020 | Does not include public health recommendations | 2020 | World Health Organization (WHO) | <https://apps.who.int/iris/handle/10665/336265> |
| Regional strategy to improve access to medicines and vaccines in the Eastern Mediterranean, 2020–2030, including lessons from the COVID-19 pandemic | Regional guideline | 2020 | World Health Organization (WHO) | <https://apps.who.int/iris/handle/10665/335952> |
| Infection prevention and control for the safe management of a dead body in the context of COVID-19: interim guidance, 4 September 2020 | Does not include public health recommendations | 2020 | World Health Organization (WHO) | <https://apps.who.int/iris/handle/10665/334156> |
| Summary report on the virtual meeting of the regional Green Light Committee for the Eastern Mediterranean, 18 June 2020 | Does not include public health recommendations | 2020 | World Health Organization (WHO) | <https://apps.who.int/iris/handle/10665/334336> |
| Strengthening the health systems response to COVID-19: technical guidance #5: adapting primary health care services to more effectively address COVID-19, 17 June 2020 | Does not include public health recommendations | 2020 | World Health Organization (WHO) | <https://apps.who.int/iris/handle/10665/332783> |
| Use of Chest Imaging in the Diagnosis and Management of COVID-19 | Does not include public health recommendations | 2020 | World Health Organization (WHO) | <https://www.ncbi.nlm.nih.gov/pmc/articles/PMC7393953/> |
| Technical specifications for pressure swing adsorption (‎PSA)‎ oxygen plants | Does not include public health recommendations | 2020 | World Health Organization (WHO) | <https://apps.who.int/iris/handle/10665/332313> |
| Effectiveness of different forms of oxygen therapy for COVID-19 management | Does not include public health recommendations | 2020 | World Health Organization (WHO) | <https://apps.who.int/iris/handle/10665/332305> |
| Management of severe / critical cases of COVID-19 with non-invasive or mechanical ventilation: based on information as at 1st June 2020 | Does not include public health recommendations | 2020 | World Health Organization (WHO) | <https://apps.who.int/iris/handle/10665/332340> |
| Operational guidance: health workforce and security | Does not include public health recommendations | 2020 | World Health Organization (WHO) | <https://apps.who.int/iris/handle/10665/341117> |
| Summary report on a WHO online consultation in response to the COVID-19 pandemic planning for rapid dissemination and implementation of the WHO consolidated guideline on self-care interventions to strengthen sexual and reproductive health in the Eastern Mediterranean Region, virtual meeting, 30 April 2020 | Does not include public health recommendations | 2020 | World Health Organization (WHO) | <https://apps.who.int/iris/handle/10665/334335> |
| Strengthening the health system response to COVID-19: technical guidance #1: maintaining the delivery of essential health care services while mobilizing the health workforce for the COVID-19 response, 18 April 2020 | Does not include public health recommendations | 2020 | World Health Organization (WHO) | <https://apps.who.int/iris/handle/10665/332559> |
| Technical specifications for invasive and non-invasive ventilators for COVID-19 | Does not include public health recommendations | 2020 | World Health Organization (WHO) | <https://apps.who.int/iris/handle/10665/331792> |
| Strengthening the health systems response to COVID-19: technical guidance #3: supply of essential medicines and health technologies, 6 April 2020 | Does not include public health recommendations | 2020 | World Health Organization (WHO) | <https://apps.who.int/iris/handle/10665/332569> |
| Strengthening the health systems response to COVID-19: technical guidance #2: creating surge capacity for acute and intensive care, 6 April 2020 | Does not include public health recommendations | 2020 | World Health Organization (WHO) | <https://apps.who.int/iris/handle/10665/332562> |
| Oxygen sources and distribution for COVID-19 treatment centres: interim guidance, 4 April 2020 | Does not include public health recommendations | 2020 | World Health Organization (WHO) | <https://apps.who.int/iris/handle/10665/331746> |
| Strengthening the health system response to COVID-19 in the WHO transmission scenarios: action points: action points for the WHO European Region (‎‎1 April 2020)‎‎ | Does not include public health recommendations | 2020 | World Health Organization (WHO) | <https://apps.who.int/iris/handle/10665/333075> |
| Strengthening preparedness for COVID-19 in cities and urban settings: interim guidance for local authorities | Does not include public health recommendations | 2020 | World Health Organization (WHO) | <https://apps.who.int/iris/handle/10665/331896?show=full> |
| Guidance for laboratories shipping specimens to WHO reference laboratories that provide confirmatory testing for COVID-19 virus: interim guidance, 31 March 2020 | Does not include public health recommendations | 2020 | World Health Organization (WHO) | <https://apps.who.int/iris/handle/10665/331639> |
| WHO recommendations to reduce risk of transmission of emerging pathogens from animals to humans in live animal markets or animal product markets, 26 March 2020 | Does not include public health recommendations | 2020 | World Health Organization (WHO) | <https://apps.who.int/iris/handle/10665/332217> |
| Risk assessment and management of exposure of health care workers in the context of COVID-19: interim guidance, 19 March 2020 | Does not include public health recommendations | 2020 | World Health Organization (WHO) | <https://apps.who.int/iris/handle/10665/331340> |
| WHO R&D Blueprint: novel Coronavirus: outline of trial designs for experimental therapeutics, January 27, 2020, Geneva, Switzerland | Does not include public health recommendations | 2020 | World Health Organization (WHO) | <https://apps.who.int/iris/handle/10665/330694> |
| Addressing noncommunicable diseases in the COVID-19 response | Does not include public health recommendations | 2022 | World Health Organization (WHO) | <https://iris.wpro.who.int/handle/10665.1/14511> |
| Advice on the use of point-of-care immunodiagnostic tests for COVID-19 scientific brief, 8 April 2020 | Not a guideline | 2020 | World Health Organization (WHO) | <https://apps.who.int/iris/handle/10665/331713> |
| Aide memoire: use of medical and non-medical/fabric masks for community outreach activities during the COVID-19 pandemic, based on current WHO guidance, 31 May 2021 | Does not include public health recommendations | 2021 | World Health Organization (WHO) | <https://apps.who.int/iris/handle/10665/341570> |
| Algorithm for COVID-19 triage and referral patient triage and referral for resource-limited settings during community transmission | Does not include public health recommendations | 2020 | World Health Organization (WHO) | <https://iris.wpro.who.int/handle/10665.1/14502> |
| An Implementation Guide for the Management of COVID-19 on Board Cargo Ships and Fishing Vessels | Does not include public health recommendations | 2022 | World Health Organization (WHO) | <https://apps.who.int/iris/handle/10665/350941> |
| Analysing and using routine data to monitor the effects of COVID-19 on essential health services practical guide for national and subnational decision-makers | Does not include public health recommendations | 2021 | World Health Organization (WHO) | <https://apps.who.int/iris/handle/10665/338689> |
| Bacille Calmette-Guérin (BCG) vaccination and COVID-19 | Does not include public health recommendations | 2020 | World Health Organization (WHO) | <https://apps.who.int/iris/handle/10665/331745> |
| Calibrating long-term non-pharmaceutical interventions for COVID-19: Principles and facilitation tools | Does not include public health recommendations | 2022 | World Health Organization (WHO) | <https://apps.who.int/iris/handle/10665/332099> |
| Clinical management of severe acute respiratory infection (‎‎‎SARI)‎‎‎ when COVID-19 disease is suspected interim guidance, 13 March 2020 | Does not include public health recommendations | 2020 | World Health Organization (WHO) | <https://apps.who.int/iris/handle/10665/331446> |
| Continuing essential sexual reproductive, maternal, neonatal, child and adolescent health services during COVID-19 pandemic practical considerations | Does not include public health recommendations | 2020 | World Health Organization (WHO) | <https://apps.who.int/iris/handle/10665/332162> |
| COVID-19 and food safety guidance for food businesses interim guidance, 07 April 2020 | Does not include public health recommendations | 2020 | World Health Organization (WHO) | <https://apps.who.int/iris/handle/10665/331705> |
| COVID-19 support mission to Turkmenistan, 6–16 July 2020 | Does not include public health recommendations | 2020 | World Health Organization (WHO) | <https://apps.who.int/iris/handle/10665/334246> |
| COVID-19 technical mission of experts to Tajikistan 1–11 May 2020 | Does not include public health recommendations | 2020 | World Health Organization (WHO) | <https://apps.who.int/iris/handle/10665/334214> |
| COVID-19 Vaccines safety surveillance manual. Module on safety surveillance of COVID-19 vaccines in pregnant and breastfeeding women | Does not include public health recommendations | 2021 | World Health Organization (WHO) | <https://apps.who.int/iris/handle/10665/342538> |
| Diagnostic testing for SARS-CoV-2 interim guidance, 11 September 2020 | Does not include public health recommendations | 2020 | World Health Organization (WHO) | <https://apps.who.int/iris/handle/10665/334254> |
| Exploration of COVID-19 health-care worker cases implications for action | Does not include public health recommendations | 2020 | World Health Organization (WHO) | <http://iris.wpro.who.int/handle/10665.1/14573> |
| Frequently asked questions: COVID-19 vaccines and breastfeeding based on WHO interim recommendations | Does not include public health recommendations | 2021 | World Health Organization (WHO) | <https://apps.who.int/iris/handle/10665/345208> |
| Immunization as an essential health service: guiding principles for immunization activities during the COVID-19 pandemic and other times of severe disruption, 1 November 2020 | Does not include public health recommendations | 2020 | World Health Organization (WHO) | <https://apps.who.int/iris/handle/10665/336542> |
| Infection prevention and control during health care when coronavirus disease (COVID-19) is suspected or confirmed: interim guidance, 12 July 2021 | Does not include public health recommendations | 2021 | World Health Organization (WHO) | <https://apps.who.int/iris/handle/10665/342620> |
| Information note on HIV and COVID-19 | Does not include public health recommendations | 2022 | World Health Organization (WHO) | <https://apps.who.int/iris/handle/10665/331919> |
| Laboratory biosafety guidance related to coronavirus disease (‎‎‎‎COVID-19)‎‎‎‎ interim guidance, 28 January 2021 | Does not include public health recommendations | 2021 | World Health Organization (WHO) | <https://apps.who.int/iris/handle/10665/339056> |
| Mitigating the impact of COVID-19 on control of vaccine-preventable diseases a health risk management approach focused on catch-up vaccination | Does not include public health recommendations | 2020 | World Health Organization (WHO) | <https://apps.who.int/iris/handle/10665/334248> |
| Role of primary care in the COVID-19 response. Interim guidance. Revised and republished as of 9 April 2021 | Does not include public health recommendations | 2021 | World Health Organization (WHO) | <https://apps.who.int/iris/handle/10665/331921> |
| SARS-CoV-2 in animals used for fur farming GLEWS+ risk assessment, 20 January 2021 | Does not include public health recommendations | 2021 | World Health Organization (WHO) | <https://apps.who.int/iris/handle/10665/339626> |
| Schools and other educational institutions transmission investigation protocol for coronavirus disease 2019 (COVID-19) | Not a guideline | 2020 | World Health Organization (WHO) | <https://apps.who.int/iris/handle/10665/336253> |
| Strategic considerations in preparing for deployment of COVID-19 vaccine and vaccination in the WHO European Region, 9 October 2020 | Does not include public health recommendations | 2020 | World Health Organization (WHO) | <https://apps.who.int/iris/handle/10665/335940> |
| Strengthening the health systems response to COVID-19 technical guidance 4 community pharmacy, 1 May 2020 | Does not include public health recommendations | 2021 | World Health Organization (WHO) | <https://apps.who.int/iris/handle/10665/332572> |
| Transmission of SARS-CoV-2 implications for infection prevention precautions | Not a guideline | 2020 | World Health Organization (WHO) | <https://apps.who.int/iris/handle/10665/333114> |
| Western Pacific Regional guide for the immunization programme and vaccine-preventable disease surveillance during the COVID-19 pandemic | Not a guideline | 2020 | World Health Organization (WHO) | <http://iris.wpro.who.int/handle/10665.1/14650> |
| WHO high-level mission to North Macedonia on coronavirus disease 2019 (‎COVID-19)‎ 23–25 June 2020 | Does not include public health recommendations | 2020 | World Health Organization (WHO) | <https://apps.who.int/iris/handle/10665/336267> |
| Why people living and working in detention facilities should be included in national COVID-19 vaccination plans: advocacy brief | Not a guideline | 2021 | World Health Organization (WHO) | <https://apps.who.int/iris/handle/10665/341497> |
| Protecting people with disability during the COVID-19 pandemic | Not a guideline | 2020 | World Health Organization (WHO) | <https://apps.who.int/iris/handle/10665/332820> |
| Implementation of WHO guidance on maintaining influenza surveillance and monitoring of SARS-CoV-2 through national surveillance systems during the COVID-19 pandemic in the SEA Region Member States | Not a guideline | 2021 | World Health Organization (WHO) | <https://apps.who.int/iris/handle/10665/350448> |
| Clinical Management of COVID-19 | Does not include public health recommendations | 2023 | World Health Organization (WHO) | <https://apps.who.int/iris/handle/10665/365580> |
| Operational considerations for respiratory virus surveillance in Europe | Not a guideline | 2022 | World Health Organization (WHO), European Center for Disease Prevention and Control (ECDC) | https://apps.who.int/iris/handle/10665/360349 |
| Operational guide for engaging communities in contact tracing | Does not include public health recommendations | 2021 | World Health Organization (WHO), International Federation of Red Cross and Red Crescent Societies (IFRC), United Nations International Children's Emergency Fund (UNICEF), Global Outbreak Alert and Response Network (GOARN) | https://apps.who.int/iris/handle/10665/341553 |
| COVID-19 vaccination: supply and logistics guidance: interim guidance, 12 February 2021 | Does not include public health recommendations | 2021 | World Health Organization (WHO), United Nations International Children's Emergency Fund (UNICEF) | https://apps.who.int/iris/handle/10665/339561 |
| Disability considerations for COVID-19 vaccination: WHO and UNICEF policy brief, 19 April 2021 | Does not include public health recommendations | 2021 | World Health Organization (WHO), United Nations International Children's Emergency Fund (UNICEF) | https://apps.who.int/iris/handle/10665/340858 |
| Monitoring COVID-19 vaccination Considerations for the collection and use of vaccination data | Does not include public health recommendations | 2021 | World Health Organization (WHO), United Nations International Children's Emergency Fund (UNICEF) | <https://apps.who.int/iris/handle/10665/339993> |
| Water, sanitation, hygiene, and waste management for SARS-CoV-2, the virus that causes COVID-19: interim guidance, 29 July 2020 | Does not include public health recommendations | 2020 | World Health Organization (WHO), United Nations International Children's Emergency Fund (UNICEF) | https://apps.who.int/iris/handle/10665/333560 |
| The management of surgical patients in the emergency setting during COVID-19 pandemic: the WSES position paper | Does not include public health recommendations | 2021 | World Society of Emergency Surgery educational board (WSES) | <https://www.ncbi.nlm.nih.gov/pmc/articles/PMC7983964/> |
| A guidance on diagnosis and management of hyperglycemia at COVID care facilities in India. | Does not include public health recommendations | 2021 | Yashdeep Gupta, Alpesh Goyal, Suraj Kubihal, Kiran Kumar Golla, Nikhil Tandon | <https://www.sciencedirect.com/science/article/pii/S1871402121000175?casa_token=lXPJJFBlf2YAAAAA:YFhDYybldH2iV5SD2cDpIKm-Spg6fSLCqHdNkm2JgZcRjQQRKa2uyZxEzIGqDolF8KNLM2rCSLw> |
| Management of critically ill patients with COVID-19 in ICU: statement from front-line intensive care experts in Wuhan, China | Does not include public health recommendations | 2020 | You Shang, Chun Pan, Xianghong Yang, Ming Zhong, Xiuling Shang, Zhixiong Wu, Zhui Yu, Wei Zhang, Qiang Zhong, Xia Zheng, Ling Sang, Li Jiang, Jiancheng Zhang, Wei Xiong, Jiao Liu, Dechang Chen | <https://annalsofintensivecare.springeropen.com/articles/10.1186/s13613-020-00689-1> |
| Guidelines on the treatment with integrated traditional Chinese medicine and western medicine for severe coronavirus disease 2019 | Does not include public health recommendations | 2021 | Zhi-Yu Li, Zhi-Jun Xie, Hai-Chang Li, Jian-Jian Wang, Xiang-Hui Wen, Shou-Yuan Wu, Jiao Chen, Juan-Juan Zhang, Lin Li, Qiang-Qiang Guo, Qiu-Ping Liu, Hui Lan, Yue-Peng Jiang, Dian-Ming Li, Xiao-Feng Xu, Si-Yue Song, Ming Zhang, Shan Fang, Wei-Dong Lai, Yi-Ni Gao, Feng-Qi Zhang, Wen-Qing Luo, Yu Lou, Wu Chen, Xia-Feng Zhang, Ke-Er Wang, Ming-Qian Zhou, Yuan-Fang He, An-Ran Xi, Yan Gao, Yi Zhang, Yao-Long Chen, Cheng-Ping Wen | <https://www.sciencedirect.com/science/article/pii/S1043661821005399?via%3Dihub> |
| Treatment of patients with nonsevere and severe coronavirus disease 2019: an evidence-based guideline | Does not include public health recommendations | 2020 | Zhikang Ye, Bram Rochwerg, Ying Wang, Neill K. Adhikari, Srinivas Murthy, François Lamontagne, Robert A. Fowler, Haibo Qiu, Li Wei, Ling Sang, Mark Loeb, Ning Shen, Minhua Huang, Zhaonan Jiang, Yaseen M. Arabi, Luis Enrique Colunga-Lozano, Li Jiang, Younsuck Koh, Dong Liu, Fang Liu, Jason Phua, Aizong Shen, Tianyi Huo, Bin Du, Suodi Zhai, Gordon H. Guyatt | <https://www.cmaj.ca/content/192/20/E536> |

### Supplement 3: Studies referenced by guidelines as evidence of values and preferences

1. Centers for Disease Control and Prevention and University of Iowa/RAND survey, unpublished.
2. NIS-CCM estimates (5-17 years) available on COVIDVaxView. cdc.gov. Published November 3, 2021. Accessed March 20, 2024. https://www.cdc.gov/vaccines/imz-managers/coverage/covidvaxview/index.html
3. KFF COVID-19 Vaccine Monitor. KFF.org. Published November 17, 2023. Accessed March 20, 2024. <https://www.kff.org/coronavirus-covid-19/dashboard/kff-covid-19-vaccine-monitor-dashboard/#parents>.
4. Sparks G, Lopes L, Montero A, Hamel L, Brodie M. KFF COVID-19 Vaccine Monitor: April 2022. KFF.org. Published May 4, 2022. Accessed March 20, 2024. <https://www.kff.org/coronavirus-covid-19/poll-finding/kff-covid-19-vaccine-monitor-april-2022>.
5. Trung T, Hoang A-D, Nguyen TT, Dinh V-H, Nguyen Y-C, Pham H-H. Dataset of Vietnamese student’s learning habits during COVID-19. *Data in Brief.* 2020;30:105682. doi:10.1016/j.dib.2020.105682
6. Margolius M, Doyle L, Pufall A, Jones E, Hynes M. The State of Young People during COVID-19: Findings from a Nationally Representative Survey of High School Youth. America’s Promise Alliance. Published June 2020. <https://files.eric.ed.gov/fulltext/ED606305.pdf>
7. Spinelli M, Lionetti F, Pastore M, Fasolo M. Parents' Stress and Children's Psychological Problems in Families Facing the COVID-19 Outbreak in Italy. *Front Psychol.* 2020;11:1713.
8. Hamel L, Lopes L, Kearney A, Sparks G, Stokes M, Brodie M. KFF COVID-19 Vaccine Monitor: June 2021. KFF.org. Published June 30, 2021. Accessed March 20, 2024. <https://www.kff.org/coronavirus-covid-19/poll-finding/kff-covid-19-vaccine-monitor-june-2021/>
9. Americans On COVID-19 Vaccine Requirements: Yes For Health Care Workers, No For Restaurant Customers, Quinnipiac University National Poll Finds; Majority Support Masking Up Indoors And At Schools. poll.qu.edu. Published August 5, 2021. Accessed March 20, 2024. <https://poll.qu.edu/poll-release?releaseid=3815>
10. Hamel L, Kirzinger A, Lopes L, Sparks G, Kearney A, Stokes M, Brodie M. KFF COVID-19 Vaccine Monitor: May 2021. KFF.org. Published May 28, 2021. Accessed March 20, 2024. <https://www.kff.org/coronavirus-covid-19/poll-finding/kff-covid-19-vaccine-monitor-may-2021/>
11. AP-NORC. Safety concerns remain main driver of vaccine hesitancy. apnorc.org. Published February 10, 2021. Accessed March 20, 2024. <https://apnorc.org/projects/safety-concerns-remain-main-driver-of-vaccine-hesitancy/>
12. Boyle J, Brassell T, Dayton J. Many Americans remain hesitant about getting a COVID-19 immunization as soon as an FDA-approved vaccine is available. icf.com. Published July 9, 2020. Accessed March 20, 2024. <https://www.icf.com/insights/health/covid-19-survey-americans-hesitant-vaccine>
13. Malik AA, McFadden SM, Elharake J, Omer SB. Determinants of COVID-19 vaccine acceptance in the US. *EClinicalMedicine*. 2020;26:100495. doi:10.1016/j.eclinm.2020.100495
14. AP-NORC. Many remain doubtful about getting COVID-19 vaccine. apnorc.org. Published December 9, 2020. Accessed March 20, 2024. <https://apnorc.org/projects/many-remain-doubtful-about-getting-covid-19-vaccine/>
15. Nguyen KH, Srivastav A, Razzaghi H, et al. COVID-19 vaccination intent, perceptions, and reasons for not vaccinating among groups prioritized for early vaccination - United States, September and December 2020. *Am J Transplant*. 2021;21(4):1650-1656. doi:10.1111/ajt.16560
16. Funk C, Tyson A. Intent to Get a COVID-19 Vaccine Rises to 60% as Confidence in Research and Development Process Increases. pewresearch.org. Published December 3, 2020. Accessed March 20, 2024. <https://www.pewresearch.org/science/2020/12/03/intent-to-get-a-covid-19-vaccine-rises-to-60-as-confidence-in-research-and-development-process-increases/>
17. Hamel L, Kirzinger A, Muñana C, Brodie M. KFF COVID-19 Vaccine Monitor: December 2020. KFF.org. Published December 15, 2020. Accessed March 20, 2024. <https://www.kff.org/coronavirus-covid-19/report/kff-covid-19-vaccine-monitor-december-2020/>
18. Szilagyi PG, Thomas K, Shah MD, et al. National Trends in the US Public’s Likelihood of Getting a COVID-19 Vaccine—April 1 to December 8, 2020. *JAMA.* 2021;325(4):396–398. doi:10.1001/jama.2020.26419
19. Oliver S. Evidence to Recommendation Framework: Moderna COVID-19 vaccine, Spikevax. Presentation to ACIP; February 4, 2022. Accessed March 20, 2024. [www.cdc.gov/vaccines/acip/meetings/downloads/slides-2022-02-04/07-COVID-Oliver-508.pdf](http://www.cdc.gov/vaccines/acip/meetings/downloads/slides-2022-02-04/07-COVID-Oliver-508.pdf)
20. CDC Covid Data tracker. cdc.gov. Accessed March 20, 2024. <https://covid.cdc.gov/covid-data-tracker/#vaccine-confidence>.
21. Jackson C, Newall M, Duran J, Rollason C, Golden J. Most Americans not worrying about COVID going into 2022 Holidays. ipsos.com. December 6, 2022. Accessed March 20, 2024. <https://www.ipsos.com/en-us/news-polls/axios-ipsos-coronavirus-index>.
22. Kirzinger A, Kearney A, Hamel L, Brodie M. KFF COVID-19 Vaccine Monitor: Early Omicron Update. KFF.org. Published December 21, 2021. Accessed March 20, 2024. <https://www.kff.org/coronavirus-covid-19/poll-finding/kff-covid-19-vaccine-monitor-early-omicron-update/>
23. Mitropoulos A. More Americans getting vaccinated following full FDA approval of Pfizer COVID vaccine. abcnews.go.com. Published August 31, 2021. Accessed March 20, 2024. <https://abcnews.go.com/Health/americans-vaccinated-full-fda-approval-pfizer-covid-vaccine/story?id=79750505>
24. Talev M. Axios-Ipsos poll: Parents split on vaccinating kids. axios.com. Published April 6, 2021. Accessed March 20, 2024. <https://www.axios.com/2021/04/06/adults-say-yes-to-the-vaccine-for-themselves-not-their-kids>
25. Ipsos. Axios/Ipsos Poll – Wave 44 Topline and Methodology. Published April 19, 2021. Accessed March 20, 2024. <https://www.ipsos.com/sites/default/files/ct/news/documents/2021-04/topline-axios-coronavirus-index-W44.pdf>
26. Calarco JM, Anderson EM. “I’m Not Gonna Put That On My Kids”: Gendered Opposition to New Public Health Initiatives. *SocArXiv*. 2021;doi:10.31235/osf.io/tv8zw.
27. Benisek A, Ratini M. COVID-19 Vaccines. Accessed March 20, 2024. <https://www.webmd.com/vaccines/covid-19-vaccine/news/20210302/webmd-survey-many-parents-back-kid-teacher-covid-shots>.
28. National Parents Union. Vaccine distrust high among parents; 40% won’t commit to having their children vaccinated. January 2021. Accessed March 20, 2024. <https://nationalparentsunion.org/2021/01/20/vaccine-distrust-high-among-parents-40-wont-commit-to-having-their-children-vaccinated/>.
29. Simonson MD, Baum M, Lazer D, et al. The COVID States Project #45: Vaccine hesitancy and resistance among parents. *OSF Preprints*. 2021;doi:10.31219/osf.io/e95bc.
30. Parents Together. First Look: Early Research on Parental Attitudes about the COVID-19 Vaccination & Children Identifies Gaps and Suggests Steps to Decrease Hesitancy. Published March 31, 2021. Accessed March 20, 2024. <https://parents-together.org/wp-content/uploads/2021/03/PT-Brief_-Parental-Attitudes-about-COVID-19-Vaccine.pdf>
31. Szilagyi PG, Shah MD, Delgado JR, et al. Parents' Intentions and Perceptions About COVID-19 Vaccination for Their Children: Results From a National Survey. *Pediatrics*. 2021;148(4):e2021052335. doi:10.1542/peds.2021-052335
32. Ruggiero KM, Wong J, Sweeney CF, et al. Parents' Intentions to Vaccinate Their Children Against COVID-19. *J Pediatr Health Care.* 2021;35(5):509-517. doi:10.1016/j.pedhc.2021.04.005
33. Brenan M. In U.S., 55% Would Get COVID-19 Vaccine for Young Child. news.gallup.com. Published September 28, 2021. Accessed March 20, 2024. <https://news.gallup.com/poll/354998/covid-vaccine-young-child.aspx>
34. Lopes L, Hamel L, Sparks G, Stokes M, Brodie M. KFF COVID-19 Vaccine Monitor: Vaccination Trends Among Children And COVID-19 In Schools. KFF.org. Published September 30, 2021. Accessed March 20, 2024. <https://www.kff.org/coronavirus-covid-19/poll-finding/kff-covid-19-vaccine-monitor-trends-among-children-school/>
35. Tsai R, Hervey J, Hoffman K, et al. COVID-19 Vaccine Hesitancy and Acceptance Among Individuals With Cancer, Autoimmune Diseases, or Other Serious Comorbid Conditions: Cross-sectional, Internet-Based Survey. *JMIR Public Health Surveill*. 2022;8(1):e29872. Published 2022 Jan 5. doi:10.2196/29872
36. Garcia P, Montez-Rath ME, Moore H, et al. SARS-CoV-2 Vaccine Acceptability in Patients on Hemodialysis: A Nationwide Survey. *J Am Soc Nephrol.* 2021;32(7):1575-1581. doi:10.1681/ASN.2021010104
37. World Health Organization. Apart Together survey: preliminary overview of refugees and migrants self-reported impact of COVID-19. who.int. Published December 18, 2020. Accessed March 20, 2024. <https://www.who.int/publications/i/item/9789240017924>
38. Loiacono MM, Mahmud SM, Chit A, et al. Patient and practice level factors associated with seasonal influenza vaccine uptake among at-risk adults in England, 2011 to 2016: An age-stratified retrospective cohort study. *Vaccine X.* 2020;4:100054. Published 2020 Jan 13. doi:10.1016/j.jvacx.2020.100054
39. Deal A, Hayward SE, Huda M, et al. Strategies and action points to ensure equitable uptake of COVID-19 vaccinations: A national qualitative interview study to explore the views of undocumented migrants, asylum seekers, and refugees. *J Migr Health.* 2021;4:100050. doi:10.1016/j.jmh.2021.100050
40. Han K, Francis MR, Zhang R, et al. Confidence, Acceptance and Willingness to Pay for the COVID-19 Vaccine among Migrants in Shanghai, China: A Cross-Sectional Study. *Vaccines (Basel)*. 2021;9(5):443. Published 2021 May 2. doi:10.3390/vaccines9050443
41. Human Rights Watch. Lebanon: refugees, migrants left behind in vaccine rollout: ensure greater information access. hrw.org. Published April 6, 2021. Accessed March 20, 2024. <https://www.hrw.org/news/2021/04/06/lebanon-refugees-migrants-left-behind-vaccine-rollout>
42. Salibi N, Abdulrahim S, El Haddad M, et al. COVID-19 vaccine acceptance in older Syrian refugees: Preliminary findings from an ongoing study. *Prev Med Rep*. 2021;24:101606. doi:10.1016/j.pmedr.2021.101606
43. Khaled SM, Petcu C, Bader L, et al. Prevalence and Potential Determinants of COVID-19 Vaccine Hesitancy and Resistance in Qatar: Results from a Nationally Representative Survey of Qatari Nationals and Migrants between December 2020 and January 2021. *Vaccines (Basel)*. 2021;9(5):471. Published 2021 May 7. doi:10.3390/vaccines9050471
44. Galvin G. Novavax’s Traditional Vaccine for COVID-19 Could Hit the U.S. Soon. Most Unvaccinated Adults Wouldn’t Be Swayed. pro.morningconsult.com. Published July 5, 2022. Accessed March 20, 2024. <https://pro.morningconsult.com/trend-setters/novavax-protein-based-covid-vaccine-survey>
45. Lopes L, Hamel L, Sparks G, Montero Am Presiado M, Brodie M.KFF COVID-19 Vaccine Monitor: July 2022. KFF.org. Published July 26, 2022. Accessed March 20, 2024. <https://www.kff.org/coronavirus-covid-19/poll-finding/kff-covid-19-vaccine-monitor-july-2022/>
46. Lazarus JV, Ratzan SC, Palayew A, et al. A global survey of potential acceptance of a COVID-19 vaccine. *Nat Med.* 2021;27, 225–228. <https://doi.org/10.1038/s41591-020-1124-9>
47. YouGov. COVID-19: Willingness to be vaccinated. yougov.co.uk. Published January 12, 2021. Accessed March 20, 2024. [https://yougov.co.uk/international/articles/33674-covid-19-willingness-be vaccinated?redirect_from=%2Ftopics%2Finternational%2Farticlesreports%2F2021%2F01%2F12%2Fcovid-19-willingness-be-vaccinated](https://yougov.co.uk/international/articles/33674-covid-19-willingness-be%20vaccinated?redirect_from=%2Ftopics%2Finternational%2Farticlesreports%2F2021%2F01%2F12%2Fcovid-19-willingness-be-vaccinated)
48. Ipsos. Global attitudes : COVID-19 vaccines. ipsos.com. Published February 9, 2021. Accessed March 20, 2024. <https://www.ipsos.com/en-ca/global-attitudes-covid-19-vaccine-january-2021>
49. Skjefte M, Ngirbabul M, Akeju O, et al. COVID-19 vaccine acceptance among pregnant women and mothers of young children: results of a survey in 16 countries. *Eur J Epidemiol.* 2021;36(2):197-211. doi:10.1007/s10654-021-00728-6
